# Comparison of Mental Health Symptoms Prior to and During COVID-19: Evidence from a Systematic Review and Meta-analysis of 134 Cohorts

**DOI:** 10.1101/2021.05.10.21256920

**Authors:** Ying Sun, Yin Wu, Suiqiong Fan, Tiffany Dal Santo, Letong Li, Xiaowen Jiang, Kexin Li, Yutong Wang, Amina Tasleem, Ankur Krishnan, Chen He, Olivia Bonardi, Jill T. Boruff, Danielle B. Rice, Sarah Markham, Brooke Levis, Marleine Azar, Ian Thombs-Vite, Dipika Neupane, Branka Agic, Christine Fahim, Michael S. Martin, Sanjeev Sockalingam, Gustavo Turecki, Andrea Benedetti, Brett D. Thombs

## Abstract

**Objectives:** The rapid pace, high volume, and limited quality of mental health evidence that has been generated during COVID-19 poses a barrier to understanding mental health outcomes. We sought to summarize results from studies that compared mental health outcomes during COVID-19 to outcomes assessed prior to COVID-19 in the same cohort in the general population and in other groups for which data have been reported.

**Design:** Living systematic review.

**Data Sources:** MEDLINE (Ovid), PsycINFO (Ovid), CINAHL (EBSCO), EMBASE (Ovid), Web of Science Core Collection: Citation Indexes, China National Knowledge Infrastructure, Wanfang, medRxiv (preprints), and Open Science Framework Preprints (preprint server aggregator).

**Eligibility criteria for selecting studies:** For this report, we included studies that compared general mental health, anxiety symptoms, or depression symptoms, assessed January 1, 2020 or later, to the same outcomes collected between January 1, 2018 and December 31, 2019. Any population was eligible. We required ≥ 90% of participants pre-COVID-19 and during COVID-19 to be the same or the use of statistical methods to address missing data. For population groups with continuous outcomes for at least two studies in an outcome domain, we conducted restricted maximum-likelihood random-effects meta-analyses. Worse COVID-19 mental health outcomes are reported as positive. Risk of bias of included studies was assessed using an adapted version of the Joanna Briggs Institute Checklist for Prevalence Studies.

**Results:** As of April 11, 2022, we had reviewed 94,411 unique titles and abstracts and identified 137 unique eligible studies with data from 134 cohorts. Almost all studies were from high-income (105, 77%) or upper-middle income (28, 20%) countries. Among adult general population studies, we did not find changes in general mental health (standardized mean difference of change [SMD_change_ = 0.11, 95% CI -0.00 to 0.22) or anxiety symptoms (SMD_change_ = 0.05, 95% CI -0.04 to 0.13), but depression symptoms worsened minimally (SMD_change_ = 0.12, 95% CI 0.01 to 0.24). Among women or females, mental health symptoms worsened by minimal to small amounts in general mental health (SMD_change_ = 0.22, 95% CI 0.08 to 0.35), anxiety symptoms (SMD_change_ = 0.20, 95% CI 0.12 to 0.29), and depression symptoms (SMD_change_ = 0.22, 95% CI 0.05 to 0.40). Of 27 other analyses across outcome domains, among subgroups other than women or females, 5 analyses suggested minimal or small amounts of symptom worsening, and 2 suggested minimal or small symptom improvements. No other subgroup experienced statistically significant changes across outcome domains. In the 3 studies with data from March to April 2020 and later in 2020, symptoms either were unchanged from pre-COVID-19 at both time points or increased initially then returned to pre-COVID-19 levels. Heterogeneity measured by the I^2^ statistic was high (e.g., > 80%) for most analyses, and there was concerning risk of bias in most studies.

**Conclusions:** High risk of bias in many studies and substantial heterogeneity suggest that point estimates should be interpreted cautiously. Nonetheless, there was general consistency across analyses in that most symptom change estimates were close to zero and not statistically significant, and changes that were identified were of minimal to small magnitudes. There were, however, small negative changes for women or females in all domains. It is possible that gaps in data have not allowed identification of changes in some vulnerable groups. Continued updating is needed as evidence accrues.

**Funding:** Canadian Institutes of Health Research (CMS-171703; MS1-173070; GA4-177758; WI2-179944); McGill Interdisciplinary Initiative in Infection and Immunity Emergency COVID-19 Research Fund (R2-42).

**Registration:** PROSPERO (CRD42020179703); registered on April 17, 2020.

---

Worldwide, more than 6.5 million deaths from the SARS-CoV-2 coronavirus disease (COVID-19) pandemic have been reported to the World Health Organization.^1^ The pandemic has disrupted the lives of people across the world due to its rapid spread, morbidity and mortality, disruption of the social fabric, toll on health care systems, and economic impact.^2, 3^ Since the beginning of the pandemic, there have been substantial concerns about effects on mental health, including possible long-term post-pandemic mental health implications, particularly among vulnerable populations.^4–6^

The sheer volume and low quality of mental health evidence that has been generated and disseminated in COVID-19, however, has posed a barrier to effective evidence synthesis, decision-making, and understanding of possible long-term mental health implications.^7–9^ Thousands of cross-sectional studies have published proportions of participants with scores above thresholds on easy-to-administer mental health scales and interpreted results as “prevalence” of mental health problems, with seemingly high levels attributable to COVID-19, despite not comparing results to pre-COVID-19 levels.^8^ These scales, however, are not intended or valid for estimating prevalence. Rather, thresholds on these scales are typically set to cast a wide net for screening, and proportions of people above thresholds dramatically overestimate prevalence compared to validated diagnostic methods.^10–14^ Making matters worse, hundreds of different measure and threshold combinations have been used during the pandemic to report mental health disorder “prevalence”. Many media stories have uncritically reported results from these studies and concluded that we are experiencing a COVID-19 “mental health pandemic” or “tsunami” of mental health consequences.^15^

Evidence from longitudinal cohorts that compare mental health symptoms pre-COVID-19 to symptoms during the pandemic is needed to assess the degree of mental health changes, the nature of any changes, and who may have been affected. No studies from prior infectious disease outbreaks have compared mental health during or after the outbreak to previously collected mental health data.^16^ Many systematic reviews on mental health symptoms in COVID-19 have been published; but, with one exception, all have reported proportions of participants above different questionnaires and thresholds in cross-sectional studies. The one exception^17^ reviewed 65 longitudinal studies published up to January 2021 and reported that there was a small overall increase in mental health symptoms in early to mid-2020 compared to pre-COVID-19 (standardized mean difference [SMD] = 0.11, 95% confidence interval [CI] 0.04 to 0.17). That review, however, searched only a limited number of English-language databases, and many otherwise eligible studies were not included because they were listed in other databases or published in other languages, including Chinese. Furthermore, more studies on mental health changes in COVID-19 have been published since the January 2021 search date of that review than prior to that date.^7^

Many countries have passed their peak pandemic period, but there is still concern that the extended burden of the pandemic and public health measures that people have endured are having important, ongoing, negative effects on mental health. Given the sheer volume of information on mental health, much based on poor-quality evidence and with misleading messaging, a synthesis and summary of studies of mental health changes in COVID-19 is critically needed to help decision makers address immediate needs, consider negative long-term mental health implications of COVID-19, and prepare for future pandemics or other society-wide disasters.

We have conducted a series of living systematic reviews^18^ on mental health in COVID-19, including a review of longitudinal studies that compare mental health in COVID-19 to mental health prior to the pandemic in the same cohort.^7, 8^ The objective of the present report was to evaluate changes in mental health symptoms in COVID-19 by comparing outcomes assessed during COVID-19 to outcomes from the same cohort of participants prior to COVID-19 in the adult general population and other population groups (e.g., sex or gender, including sex or gender minorities; children and adolescents; young adults; older adults; university students; parents; and people at risk due to pre-existing medical conditions, pre-existing mental health conditions, and work as medical staff).

## METHODS

Our series of systematic reviews on mental health in COVID-19, including the review of studies of symptoms prior to and during COVID-19, was registered in the PROSPERO prospective register of systematic reviews (CRD 42020179703). A protocol was uploaded to the Open Science Framework (https://osf.io/96csg/) prior to initiation.^19^ Results from studies included in our reviews are posted online (https://www.depressd.ca/covid-19-mental-health).^7^ The present report is a subset of our overall review of longitudinal studies and includes evidence from studies that assessed general mental health, anxiety symptoms, or depression symptoms during COVID-19 and prior to the pandemic. Results are reported in accordance with the PRISMA statement.^20^

## Eligible Studies

Studies on any population, regardless of COVID-19 infection status, were included in the present report if they compared eligible outcomes assessed between January 1, 2018 and December 31, 2019, when China first reported COVID-19 to the World Health Organization,^21^ to the same outcomes collected January 1, 2020 or later. We required studies to report data from comparison samples with at least 90% of the same participants pre- and during COVID-19 or to use statistical methods to account for missing participant data. We did not include repeated cross-sectional surveys. Studies with < 100 participants were excluded for feasibility reasons and due to their limited relative value in evaluating mental health changes.

Eligible outcomes in our overall systematic review of longitudinal studies included (1) continuous scores on a validated mental health symptom questionnaire; (2) the proportion of participants above a threshold on a validated mental health symptom questionnaire; or (3) the proportion of participants meeting diagnostic criteria for a mental disorder using a validated diagnostic interview. In the overall systematic review, mental health outcomes were defined broadly to include, for example, symptoms of anxiety, symptoms of depression, general mental health, stress, loneliness, anger, grief, burnout, other emotional disturbances, or emotional well-being. In the present report, we included only general mental health, anxiety symptoms, and depression symptoms because few studies reported on other outcome domains. General mental health included measures of mental health quality of life, general symptoms or well-being, and combined symptom domains (e.g., a single measure of symptoms of anxiety and depression). Results from outcome domains not included in the present report are available online (https://www.depressd.ca/covid-19-mental-health).

## Identification and Selection of Eligible Studies

The same search strategies were used for all research questions in our systematic reviews. We searched MEDLINE (Ovid), PsycINFO (Ovid), CINAHL (EBSCO), EMBASE (Ovid), Web of Science Core Collection: Citation Indexes, China National Knowledge Infrastructure, Wanfang, medRxiv (preprints), and Open Science Framework Preprints (preprint server aggregator), using a search strategy designed and built by an experienced health sciences librarian. The China National Knowledge Infrastructure and Wanfang databases were searched using Chinese search terms chosen based on our English-language search strategy. Because of the urgent need for synthesized evidence in the early pandemic, we did not delay our project launch to formally peer review the search strategy; however, COVID-19 terms were developed in collaboration with other librarians working on the topic and updated as COVID-19-specific subject headings became available. See Supplementary Material 1 for all search strategies. Our initial search was conducted from December 31, 2019 to April 13, 2020, then automated searches were set for daily updates. On December 28, 2020, we converted to weekly updates to improve processing efficiency.

Search results were uploaded into the systematic review software DistillerSR (Evidence Partners, Ottawa, Canada), where duplicate references were identified and removed. Two independent reviewers evaluated titles and abstracts in random order. If either reviewer deemed a study potentially eligible, a full-text review was completed, also by two independent reviewers. Discrepancies at the full-text level were resolved through consensus, with a third investigator consulted as necessary. To ensure accurate identification of eligible studies, a coding guide with inclusion and exclusion criteria was developed and pre-tested, and all team members were trained over several sessions. See Supplementary Material 2.

## Data Extraction and Synthesis

For each included study, one reviewer extracted data using a pre-specified standardized form, and a second reviewer validated the extracted data using the DistillerSR Quality Control function. This function allows the second reviewer to review and edit all extracted data in the DistillerSR platform and then flags any edits as conflicts between reviewers to resolve. Reviewers extracted (1) publication characteristics (e.g., first author, publication year, journal); (2) population characteristics and demographics, including study eligibility criteria, recruitment method, number of participants, timing of assessments, age, and population group (adult general population, older adults, young adults, children and adolescents, parents, university students, people with pre-existing medical conditions, medical staff, and groups defined by sex or gender with studies in the present report, though we extracted any subgroups for which we found data); (3) mental health assessment measures and outcomes; and (4) adequacy of study methods and reporting. We used World Bank classifications to classify income and region of countries where studies were conducted.^22^ We used an adapted version of the Joanna Briggs Institute Checklist for Prevalence Studies, which includes items that assess risk of bias and adequacy of study methods and reporting. Items assessed the appropriateness of the sampling frame for the target population, appropriateness of recruiting methods, adequacy of sample size, description of setting and participants, participation or response rate, methods for outcome assessment, standardization of assessments across participants, appropriateness of statistical analyses, and follow-up rate.^23^ See Supplementary Material 3.

For each continuous outcome, we extracted an SMD effect size with 95% confidence intervals (CIs) for the change from pre-COVID-19 to during COVID-19. If not provided, we calculated it using Hedges’ g^24^ as g = mean_change_/standard deviation_within_ x the Hedges’ g adjustment factor, as described by Borenstein et al.^25^ In this report, we present SMDs as positive when mental health worsened from pre-COVID-19 to COVID-19 and negative when it improved. For pre-COVID-19 and COVID-19 proportions, if 95% CIs were not reported, we calculated a 95% CI using Agresti and Coull’s approximate method for binomial proportions.^26^ For changes in proportions, if 95% CIs were not reported, we generated them using Newcombe’s method for differences between binomial proportions based on paired data.^27^ Doing this requires knowing the number of participants above a threshold at both assessment points, which is not always available. If it was not available, we assumed that 50% of cases above a threshold pre-COVID-19 continued to be above the threshold during COVID-19. We confirmed that results did not differ substantively if we assumed values within a plausible range of 30% to 70%.

We prioritized continuous data due to pitfalls in interpreting proportions of participants crossing a dichotomous threshold. See Box 1 on interpreting outcomes from mental health symptom measures. For each population group with continuous outcomes for at least two studies in an outcome domain, SMDs were pooled across studies via restricted maximum-likelihood random-effects meta-analysis. Heterogeneity was assessed with the I^2^ statistic. For studies where more than one continuous outcome in a domain was assessed (e.g., two depression symptom measures), we pooled relevant SMDs prior to fitting the meta-analysis, so that each unique sample contributed only one observation. For studies where results were reported separately for more than one unique sample group (e.g., cancer survivors and healthy controls), SMDs were presented and included separately. For one study^28^ that calculated change based on both a difference with the last pre-COVID-19 cohort assessment and via a fixed effects regression that included all pre-COVID-19 assessments, we included unadjusted estimates from the fixed effects regression model that estimated within-person change. Meta-analyses were performed in R (R version 3.6.3, Rstudio Version 1.2.5042) using the rma.uni function in the metafor package.^29^ Forest plots were generated using the forest.rma function in metafor. We referred to commonly used metrics to characterize changes as minimal (SMD < 0.20), small (SMD = 0.20), medium (SMD = 0.50), or large (SMD = 0.80).^30^

### Box 1.

Interpreting SMD Effect Sizes and Changes in Proportion Above a Threshold on Mental Health Measures

Symptom changes assessed with mental health patient-reported outcome measures in COVID-19 have been reported as changes in continuous scores and the proportion of study participants above a threshold. Continuously measured symptom changes are presented in terms of SMDs, which describe change in terms of within-group standard deviations, rather than raw change scores, which are measure-specific and not easily compared across measures. To illustrate, Box 1 – Figure 1 illustrates the amount of change, assuming a normal distribution, for SMD = 0.25. The hypothetical blue distribution represents pre-COVID-19 scores, and the grey distribution represents post-COVID-19 scores with a mean symptom increase of SMD = 0.25.

When studies report an increase or decrease in the proportion of participants above a measure threshold, dichotomous thresholds used for this purpose are sometimes labelled as thresholds for “clinically significant” symptoms or as reflecting the presence of a condition (e.g., depression).^10^ These designations, are not, however, based on evidence that a threshold represents a meaningful divide between impairment and non-impairment and do not reflect the presence of a mental disorder. Most commonly, they reflect a point on a measure that balances sensitivity and specificity when used for screening, which does not inform when score levels might become clinically meaningful.^10–14^

Thresholds on different symptom measures are often located at different places in the symptom distribution. This can lead to divergent estimates of proportions crossing a threshold, depending on the measure used, rather than because of actual differences in symptom changes. As shown in Box 1 – Figure 1, the same change in symptoms in a hypothetical study sample would result in a 7% increase in participants at or above the threshold on one measure (black line, one standard deviation above pre-COVID-19 distribution mean) but an increase of only 2% on another (red line, two standard deviations above pre-COVID-19 distribution mean).

We have prioritized interpretation of continuous score changes. We have also reported proportions above thresholds, as they can be informative, such as when they are reported for two time points in the same study or as an indicator if some level of change may have occurred. We have, however, avoided interpretation of the magnitudes of proportions above thresholds.

## Patient and Public Involvement

Dr. Sarah Markham, who is an experienced patient advisor and member of BMJ’s International Patient Panel, was included as a member of the research team from the inception of the project. She provided input on the project design, underwent training on procedures used in the study, and was involved in selection of eligible studies. She reviewed and provided comments on the content of this article.

## Amendments to Protocol

Our systematic review was rapidly designed and initiated in April 2020, and several amendments or clarifications were made. First, we changed from daily to weekly search updates on December 28, 2020 for more efficient reference processing. Second, on January 27, 2021, we made a minor change to the MEDLINE search strategy to incorporate the new Physical Distancing Medical Subject Heading created by the National Library of Medicine in light of the COVID-19 pandemic. Third, we made several amendments to Chinese-language search strategies to facilitate processing (see Supplementary Material 1). Fourth, we added a criterion to stipulate that eligible pre-COVID-19 assessments had to be completed between January 1, 2018 and December 31, 2019. We added this criterion because we had not anticipated that studies would report comparisons of outcomes during COVID-19 to outcomes assessed many years prior, which in some cases occurred during a different developmental life stage from assessments carried out during the pandemic.

## RESULTS

### Search Results and Selection of Eligible Studies

As of April 11, 2022, we had identified 94,411 unique titles and abstracts from our database searches. Of these, we excluded 92,457 after title and abstract review and 1,523 after full-text review, leaving 431 studies with longitudinal data collection. Of those, 276 studies only assessed outcomes longitudinally during the pandemic period and did not include pre-COVID-19 data, 11 studies only assessed outcomes (e.g., stress, loneliness) not included in the present report, 1 study used the same outcome measure but for different time periods pre-COVID-19 (worst month in last year) and COVID-19 (last month), and 6 studies reported data that were from the same dataset as another study, leaving 137 unique studies that reported data from 134 cohorts for inclusion (Figure 1).

**Figure 1.**
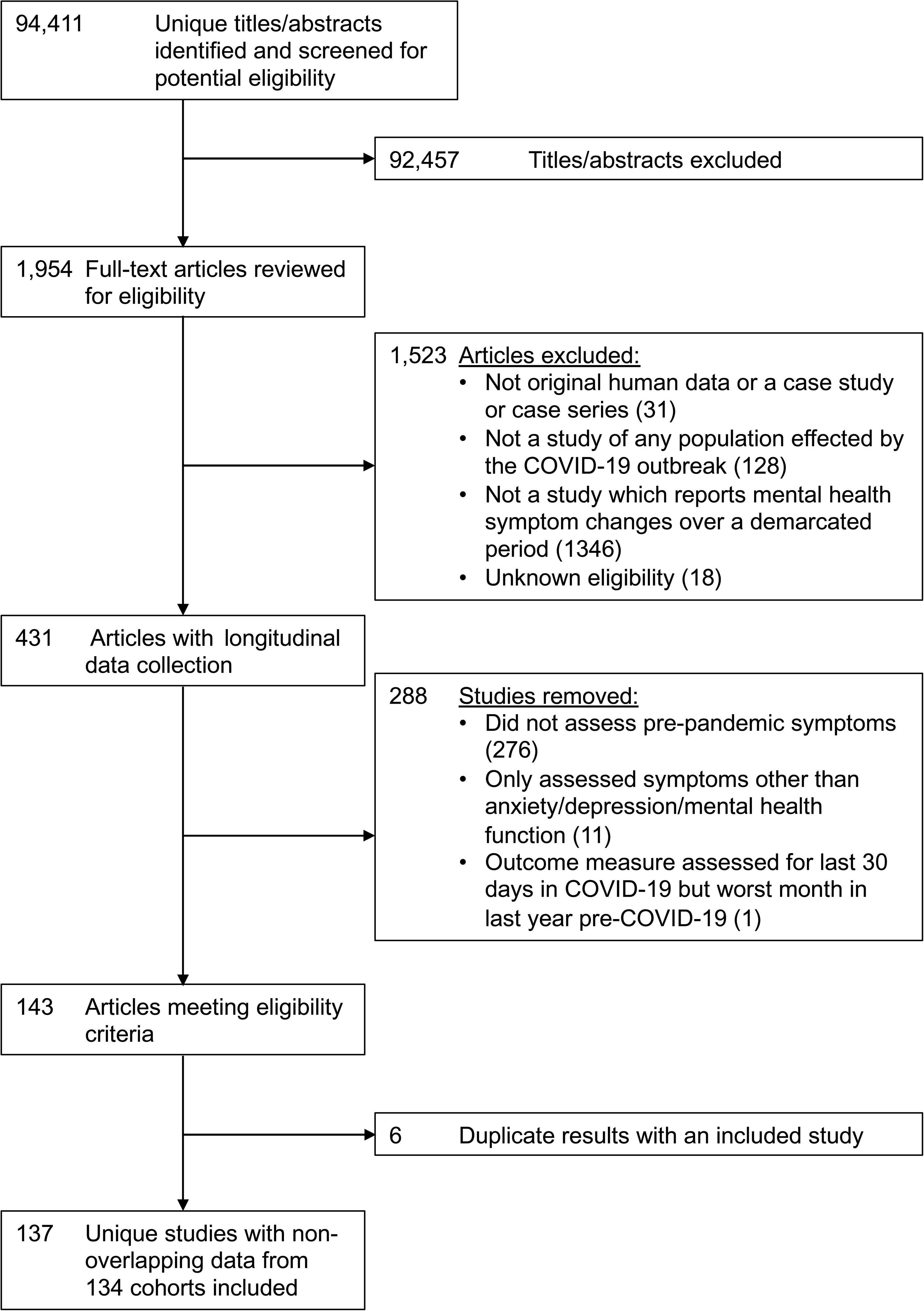
PRISMA flow diagram

## Characteristics of Included Studies

Supplementary Table 1 shows characteristics of included studies.^S1-S137^ All cohorts reported COVID-19 outcome data collected in 2020, including 4 studies that reported a single data collection period that bridged 2020 and 2021.^S74,S96,S114,S128^ All studies reported data from March 2020 or later except for 7 studies from China,^S5,S52,S54,S79,S99,S121,S133^ 1 study from Japan,^S82^ and 1 study from Taiwan^S130^ that reported data from January or February 2020. Large national probability-based cohorts from the United Kingdom^S11,S12^ and the Netherlands^S16,S17^ and a cohort of people with a pre-existing medical condition (systemic sclerosis)^S118^ reported data collected at multiple time points during 2020. The systemic sclerosis study also reported data collected at 3 time points in 2021,^S118^ but no other studies reported 2021 outcomes for all participants. Of the 137 included studies, 105 (77%) were from high-income (New Zealand = _2S1,S119; Italy = 4S2,S30,S88,S126; United States = 24S4,S6,S18,S29,S40,S48,S50,S59-S61,S71,S77,S92,S102,S103,S108,S109,S111,S112,S114, S120, S122, S131,S135; Finland = 1S9; Spain = 5S10,S27,S76,S81,S116; United Kingdom = 13S11,S12,S21,S28,S39,S42,S47,S63,S64,S83,S84,S98,S132; Japan = 9S13,S24,S41,S65,S73,S82,S115,S127,S128; Denmark = 2S15,S22; the Netherlands = 9S16,S17,S32,S33,S53,S72,S75,S110,S113; Australia = 5S19,S37,S87,S125,S134; Ireland = 1S20; Chile = 1S23; Sweden_ = 2^S25,S96^; Singapore = 3^S26,S31,S36^; Hong Kong, China = 2^S3,S35^; Switzerland = 2^S38,S46^; Canada = _4S43,S51,S104,S136; Portugal = 2S44,S89; Lithuania = 2S49,S80; Germany = 5S67,S90,S91,S93,S124; Israel =_ 1^S94^; Taiwan = 1^S130^; France = 1^S137^; multiple countries = 4^S8,S66,S118,S123^), 28 (20%) from upper-_middle-income (China = 23S5,S34,S45,S52,S54-S58,S68-S70,S78,S79,S85,S86,S95,S97,S99-S101,S121,S133; Turkey =_ 2^S7,S117^; Brazil = 2^S74,S105^; Mexico = 1^S107^), 1 (1%) from mixed high-income and upper-middle-income (Italy and Paraguay)^S129^, 3 (2%) from lower-middle-income (Iran = 1^S14^; India = 1^S62^; Bangladesh = 1^S106^), and none from low-income countries. By region, there were 52 studies _from Europe and Central Asia,S2,S7,S9-S12,S15-S17,S20-S22,S25,S27,S28,S30,S32,S33,S38,S39,S42,S44,S46,S47,S49,S53,S63,S64,S66,S67,S72,S75,S76,S80,S81,S83,S84,S88-S91,S93,S96,S98,S110,S113,S116,S117,S124,S126,S132,S137 46 from East Asia and the Pacific,S1,S3,S5,S13,S19,S24,S26,S31,S34-S37,S41,S45,S52,S54-S58,S65,S68-S70,S73,S78,S79,S82,S85-S87,S95,S97,S99-S101,S115,S119,S121,S123,S125,S127,S128,S130,S133,S134 28 from North America,S4,S6,S18,S29,S40,S43,S48,S50,S51,S59-S61,S71,S77,S92,S102-S104,S108,S109,S111,S112,S114, S120, S122, S131,S135,S136 4 from Latin America and the_ Caribbean,^S23,S74,S105,S107^ 2 from Middle East and North Africa,^S14,S94^ 2 from South Asia,^S62,S106^ 2 from mixed Europe and North American samples,^S8,S118^ 1 from a mixed Europe and Latin America and the Caribbean sample,^S129^ and none from Sub-Saharan Africa.

There were 18 studies^S1-S18^ that reported on 16 different adult general population cohorts, including large national probability-based samples from the United Kingdom (N = 10,918 to 15,376),^S11,S12^ Denmark (N = 4,234),^S15^ and the Netherlands (N = 3,983 to 4,064)^S16,S17^ and 13 non-probabilistic convenience samples with 102 to 3,124 participants from New Zealand,^S1^ Italy,^S2^ China,^S3,S5^ the United States,^S4,S6,S18^ Turkey,^S7^ Finland,^S9^ Spain,^S10^ Japan,^S13^ Iran,^S14^ and from multiple countries via an online crowdsourcing platform.^S8^

There were 18 studies with data on older adults,^S19-S36^ including one (N = 1,679)^S33^ that reported subgroup data from the large Dutch national probability sample,^S16,S17^ and other samples of at least 1,000 participants from Australia (N = 1,671),^S19^ Ireland (N = 3,490),^S20^ the United Kingdom (N = 3,281),^S21^ Sweden (N = 1,071),^S25^ the Netherlands (N = 1,068),^S32^ and China (N = 2,745).^S34^ Eleven other studies from Denmark,^S22^ Chile,^S23^ Japan,^S24^ Singapore,^S26,S31,S36^ Spain,^S27^ Scotland,^S28^ the United States,^S29^ Italy,^S30^ and Hong Kong, China,^S35^ included between 104 and 721 participants.

There were 7 studies of young adults^S37-S43^ from Australia,^S37^ Switzerland,^S38^ the United Kingdom,^S39,S42^ the United States,^S40^ Japan,^S41^ and Canada,^S43^ which assessed between 1,039 and 3,694 participants. There were also 28 studies of university students,^S44-S71^ including 10 from China,^S45,S52,S54-S58,S68-S70^ 6 from the United States,^S48,S50,S59-S61,S71^ three from the United Kingdom,^S47,S63,S64^ and one each from Portugal,^S44^ Switzerland,^S46^ Lithuania,^S49^ Canada,^S51^ the Netherlands,^S53^ India,^S62^ Japan,^S65^ combined Germany and Lithuania,^S66^ and Germany.^S67^ Of these, 9 included at least 1,000 participants (1,004 to 8,079).^S45,S50,S55,S57-S59,S65,S68,S70^

There were 30 studies of children and adolescents,^S72-S101^ including 27 that focused mostly or entirely on adolescents (ages 10 to 19^31^),^S72-S77,S80-S94,S96-S101^ 3 mixed studies of children (ages up to 9 years^31^) and adolescents,^S78,S79,S95^ and none that focused only on children. There were studies with at least 1,000 participants from Japan,^S73,S82^ the United Kingdom,^S84^ China,^S86,S95,S97,S100,S101^ Italy,^S88^ Portugal,^S89^ and Israel^S94^ plus smaller studies from the Netherlands,^S72,S75^ Brazil,^S74^ Spain,^S76,S81^ the United States,^S77,S92^ China,^S78,S79,S85,S99^ Lithuania,^S80^ the United Kingdom,^S83,S98^ Australia,^S87^ Germany,^S90, S91,S93^ and Sweden.^S96^ Studies with data on adolescents from the Netherlands^S72^ and Spain^S76^ also reported data from parents, as did 7 additional studies from the United States,^S102,S103,S108^ Canada,^S104^ Brazil,^S105^ Bangladesh,^S106^ and Mexico,^S107^ one of which included over 1,000 participants (N = 1,136).^S105^

There were 22 studies of people with pre-existing medical conditions,^S29,S35,S109-S128^ including a study of 2,829 older adults with type 2 diabetes from the United States,^S111^ a study of 2,176 patients with colorectal cancer from the Netherlands,^S113^ and a study of 1,504 participants with rheumatic diseases from the United States.^S120^ Nineteen other studies from the United _States,S29,S109,S112,S114,S122 the Netherlands,S110 Japan,S115,S127,S128 Spain,S116 Turkey,S117 New_ Zealand,^S119^ China,^S121^ Germany,^S124^ Australia,^S125^ Italy,^S126^ Hong Kong, China,^S35^ and multiple countries,^S118,S123^ included between 104 and 852 participants. There were also 4 studies of people with pre-existing mental health conditions, including a study of 12,653 people from the UK with a pre-COVID-19 depressive or anxiety disorder diagnosis^S132^ and 3 studies of 110 to 144 outpatients from Italy or Paraguay,^S129^ Taiwan,^S130^ and the United States.^S132^

There were two studies of medical workers,^S103,S133^ including a study of 180 physicians who were also parents from the United States^S103^ and a study of 385 physicians in training from China.^S133^

There were three studies of people who identified as sexual or gender minorities, including 681 gay and bisexual men from Australia,^S134^ 2,288 people with a range of gender identities from the United States,^S135^ and 780 trans and non-binary individuals from Canada.^S136^

## Risk of Bias and Adequacy of Study Methods and Reporting

Ratings of risk of bias and adequacy of methods and reporting are shown in Supplementary Table 2. Overall, only the national probability-based cohort from the Netherlands^S16,S17,S33^ was rated “Yes” on all items. Overall, 37 of 137 studies (27%) used sampling frames that were close representations of the target population; 32 of 137 (23%) used census or random sampling methods; 13 of 137 (9%) had response rates of at least 75% or established that the sample was representative, and 43 of 137 (31%) successfully followed up with at least 75% of participants or included methods to address dropout considerations. For adequate sample size, participant and setting description, use of valid assessment methods (which was an inclusion requirement for our systematic review), standard outcome collection methods, and appropriately analysed results, proportions with “Yes” ratings were between 73% and 100%.

## Changes in Mental Health Symptoms

Changes in mental health symptoms for individual studies by population category are shown in Supplementary Table 3 for general mental health, Supplementary Table 4 for anxiety symptoms, and Supplementary Table 5 for depression symptoms. Table 1 shows meta-analyses results for the general population and other populations for continuously measured general mental health, anxiety symptoms, and depression symptoms.

**Table 1.** Meta-analyses of Continuous General Mental Health, Anxiety Symptoms, and Depression Symptoms by Population Group^a^

|  | General Mental Health |  |  | Anxiety Symptoms |  |  | Depression Symptoms |  |  |
| --- | --- | --- | --- | --- | --- | --- | --- | --- | --- |
|  | Hedges' g |  |  | Hedges' g |  |  | Hedges' g |  |  |
|  | N Cohorts | Standardized Mean | I<sup>2</sup> | N Cohorts | Standardized Mean | I<sup>2</sup> | N Cohorts | Standardized Mean | I<sup>2</sup> |
|  | (Participants) | Difference (95% CI) |  | (Participants) | Difference (95% CI) |  | (Participants) | Difference (95% CI) |  |
| General Population | 11 (30,185) | 0.11 (-0.00 to 0.22) | 97% | 4 (2,632) | 0.05 (-0.04 to 0.13) | 37% | 4 (3,470) | 0.12 (0.01 to 0.24) | 81% |
| Women or Females | 6 (10,329) | 0.22 (0.08 to 0.35) | 91% | 5<sup>b</sup> (3,500) | 0.20 (0.12 to 0.29) | 41% | 7<sup>b</sup> (3,851) | 0.22 (0.05 to 0.40) | 89% |
| Men or Males | 6 (11,546) | 0.11 (-0.12 to 0.35) | 98% | 4 (1,271) | 0.07 (-0.01 to 0.14) | 0% | 7 (3,905) | 0.01 (-0.14 to 0.16) | 82% |
| Older Adults | 11 (9,960) | -0.01 (-0.12 to 0.11) | 93% | 6<sup>b</sup> (7,193) | 0.14 (-0.00 to 0.28) | 93% | 7<sup>b</sup> (7,419) | 0.22 (0.06 to 0.38) | 95% |
| Young Adults | 2 (4,221) | 0.16 (-0.07 to 0.39) | 96% | 2 (4,602) | 0.05 (-0.16 to 0.27) | 95% | 4 (8,043) | 0.02 (-0.15 to 0.18) | 96% |
| University Students | 6<sup>c,d</sup> (6,957) | 0.00 (-0.17 to 0.17) | 95% | 16<sup>c</sup> (12,642) | -0.07 (-0.21 to 0.06) | 96% | 19<sup>c</sup> (26,164) | 0.14 (0.01 to 0.26) | 98% |
| Children and Adolescents | 16 (11,505) | 0.19 (-0.05 to 0.42) | 99% | 8 (12,064) | 0.02 (-0.12 to 0.16) | 96% | 10 (11,679) | 0.06 (-0.08 to 0.20) | 96% |
| Parents | 3 (932) | 0.39 (0.21 to 0.56) | 57% | 1 (147) | 0.25 (0.02 to 0.49) | ----- | 5 (1,639) | 0.15 (-0.05 to 0.35) | 87% |
| People with Pre-existing Medical Conditions | 12 (6,511) | 0.10 (-0.01 to 0.20) | 86% | 11 (5,775) | 0.08 (-0.04 to 0.21)<sup>e,f</sup> | 89% | 16 (21,594) | 0.04 (-0.04 to 0.12)<sup>g,h</sup> | 90% |
| People with Pre-existing Mental Health Conditions | 2 (457) | -0.22 (-0.35 to -0.09) | 0% | 3 (12,362) | 0.12 (-0.11 to 0.35) | 80% | 3 (12,352) | -0.05 (-0.08 to -0.03) | 0% |
| Medical Staff | ----- | ----- | ----- | ----- | ----- | ----- | 1 (180) | 0.11 (-0.09 to 0.32) | ----- |
| Sexual or Gender Minorities | ----- | ----- | ----- | 3 (3,743) | 0.23 (-0.09 to 0.54) | 98% | 3 (3,741) | 0.19 (0.10 to 0.28) | 67% |

<sup>a</sup>Positive Hedges' g effect sizes indicate worse mental health in COVID-19 compared to pre-COVID-19. <sup>b</sup>Cancer survivors and matched controls from Rentscher study counted as one cohort although shown separately in forest plot. <sup>c</sup>Students from Lithuania and Germany from Truskauskaite-Kuneviciene study counted as one cohort although shown separately in forest plot. <sup>d</sup>Sensitivity analysis conducted with results from Savage et al. from October 2020 instead of April 2020; SMD = 0.00 (-0.17 to 0.17), I<sup>2</sup> = 94.8%. <sup>e</sup>Sensitivity analysis conducted with results from Henry et al. from September to October 2020 instead of April 2020; SMD = 0.05 (-0.04 to 0.14), I<sup>2</sup> = 76%. <sup>f</sup>Sensitivity analysis conducted with results from Henry et al. from March 2021 instead of April 2020; SMD = 0.04 (-0.04 to 0.13), I<sup>2</sup> = 76%. <sup>g</sup>Sensitivity analysis conducted with results from Henry et al. from September to October 2020 instead of April 2020; SMD = 0.03 (-0.05 to 0.11), I<sup>2</sup> = 91%. <sup>h</sup>Sensitivity analysis conducted with results from Henry et al. from March 2021 instead of April 2020; SMD = 0.03 (-0.05 to 0.11), I<sup>2</sup> = 91%.

### General Mental Health

Forest plots are shown in Figures 2a to 2j. Estimated reduction in general mental health in the general population was minimal and not statistically significant (Figure 2a; 11 cohorts, N = 30,185; SMD_change_ = 0.11, 95% CI -0.00 to 0.22; I^2^ = 97%). Among subgroups, there was a small, statistically significant worsening for women or females (Figure 2b; 6 cohorts, N = 10,329; SMD_change_ = 0.22, 95% CI 0.08 to 0.35; I^2^ = 91%) and a small to medium, statistically significant worsening for parents (Figure 2h; 3 cohorts, N = 932; SMD_change_ = 0.39, 95% CI 0.21 to 0.56; I^2^ = 57%). Symptoms improved by a small amount among people with pre-existing mental health conditions (Figure 2j; 2 cohorts, N = 457; SMD_change_ = -0.22, 95% CI -0.35 to -0.09; I^2^ = 0%). No other subgroup change estimates were statistically significantly different from zero. The percentage of variance due to heterogeneity (I^2^) across analyses was high (57% to 99%), except for among people with pre-existing mental health conditions (0%).

**Figure 2a.**
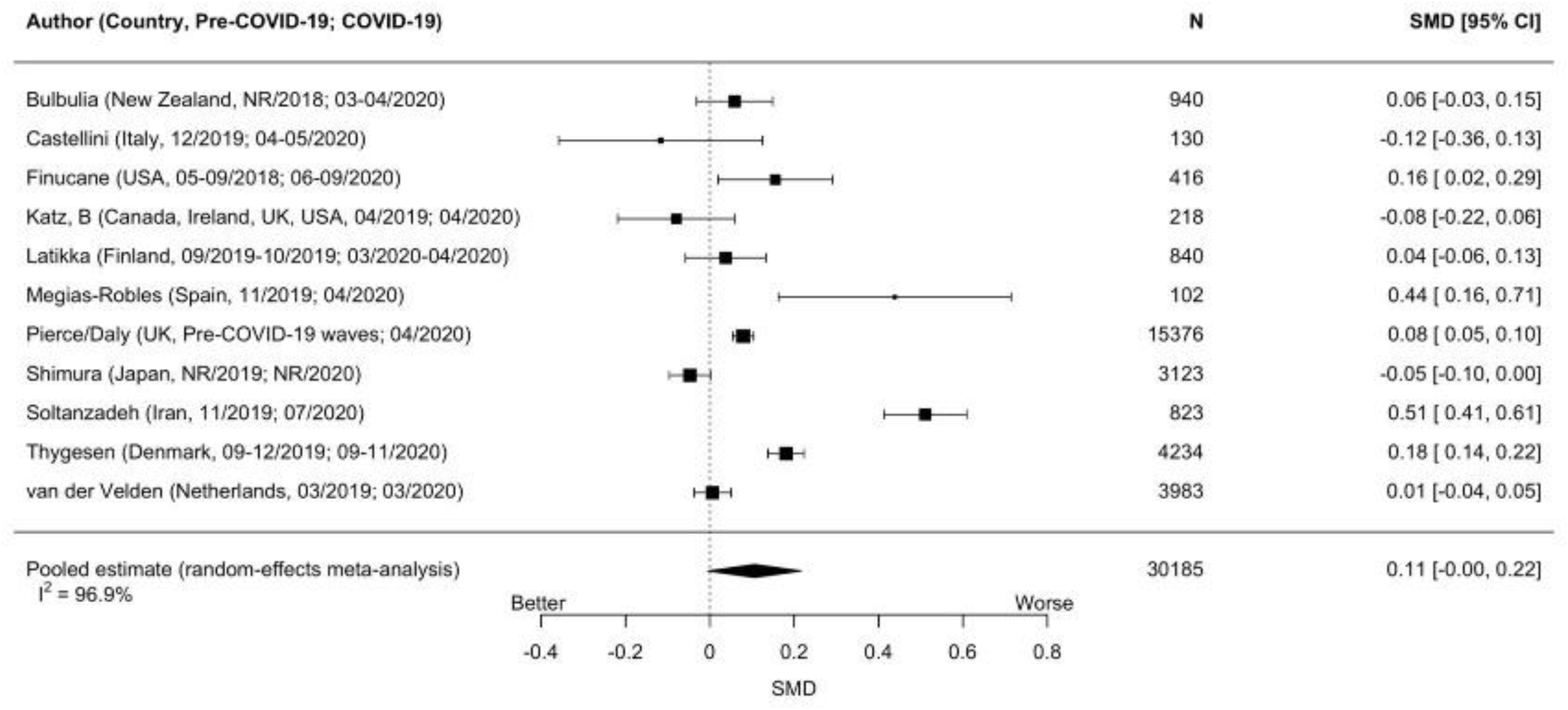
Forest plots of standardized mean difference changes in general mental health for studies of the general population

**Figure 2b.**
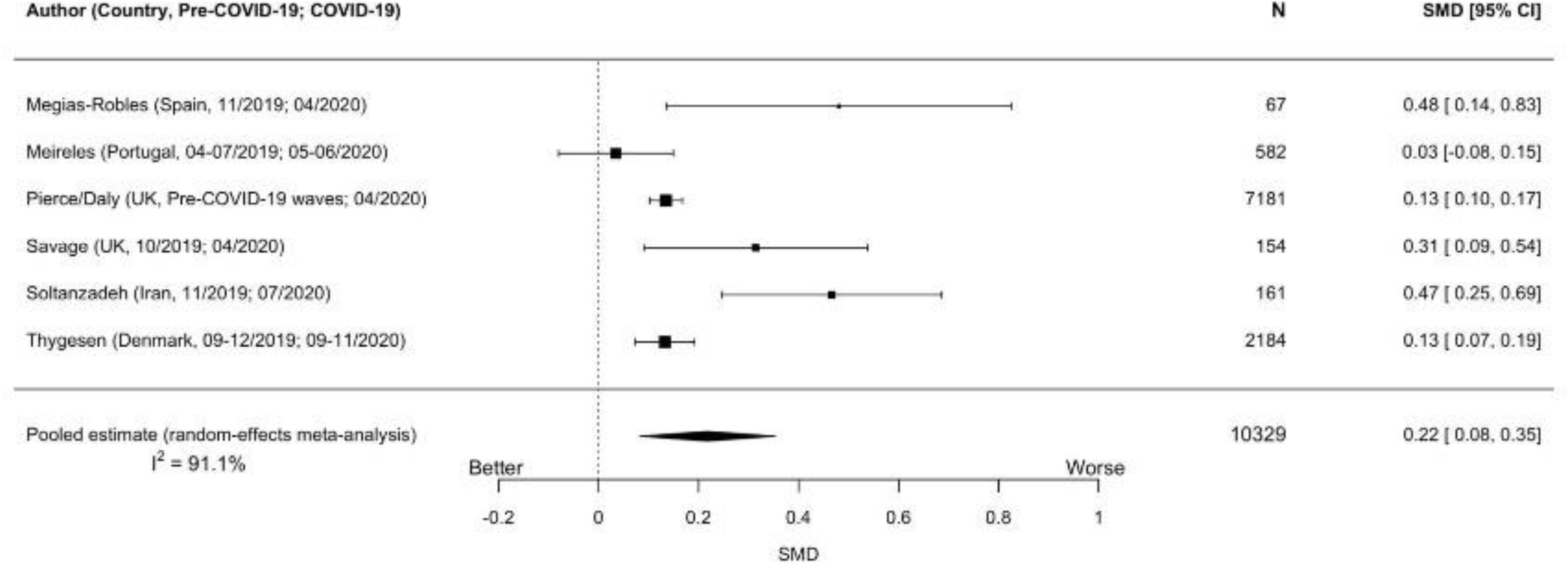
Forest plots of standardized mean difference changes in general mental health for studies of women or females

**Figure 2c.**
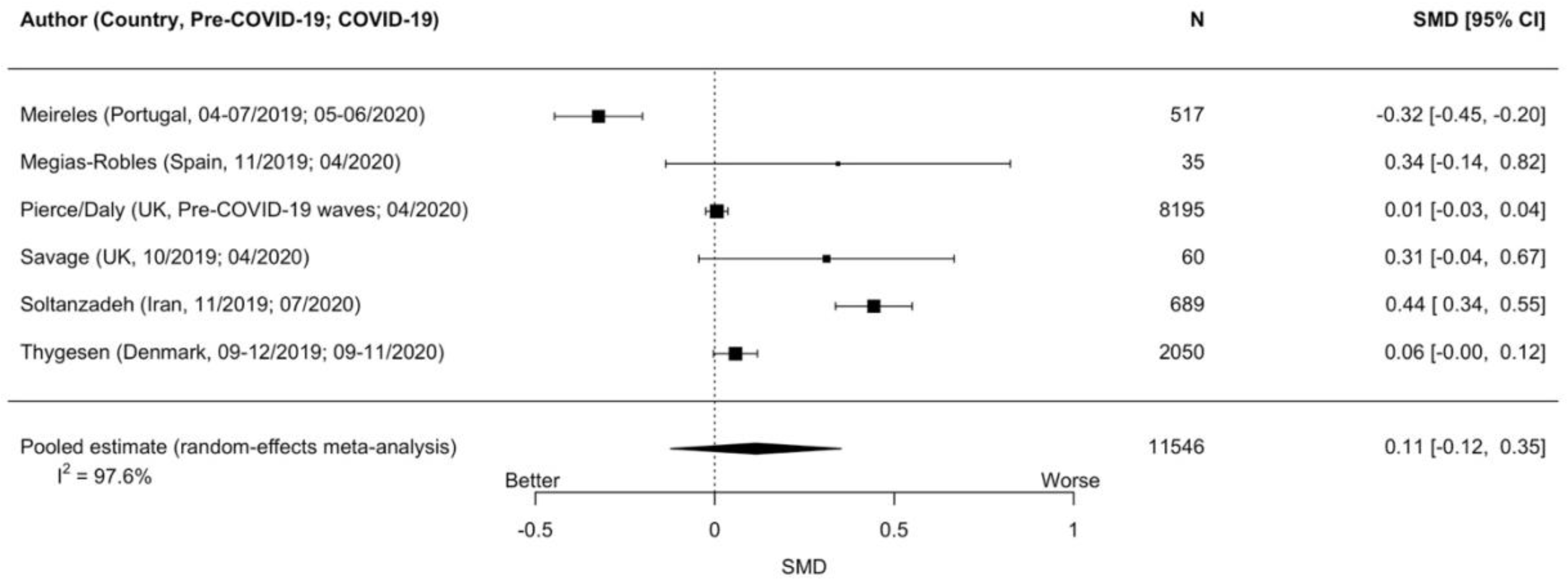
Forest plots of standardized mean difference changes in general mental health for studies of men or males

**Figure 2d.**
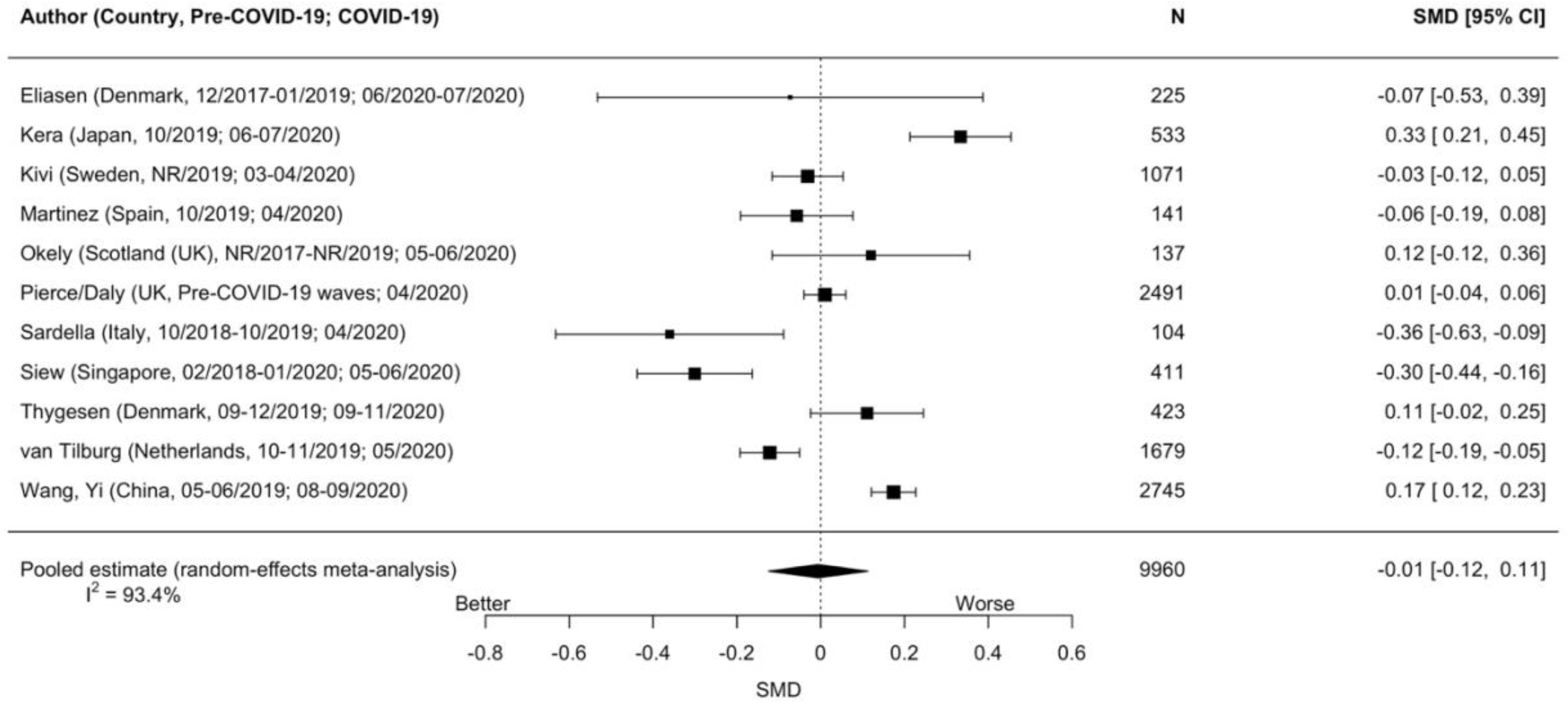
Forest plots of standardized mean difference changes in general mental health for studies of older adults

**Figure 2e.**
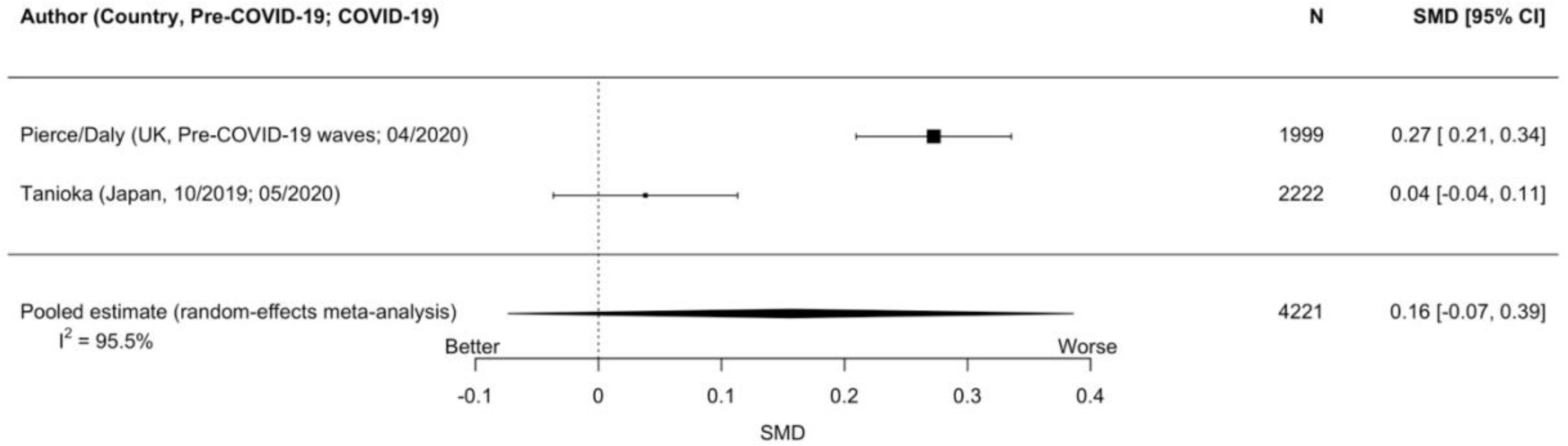
Forest plots of standardized mean difference changes in general mental health for studies of young adults

**Figure 2f.**
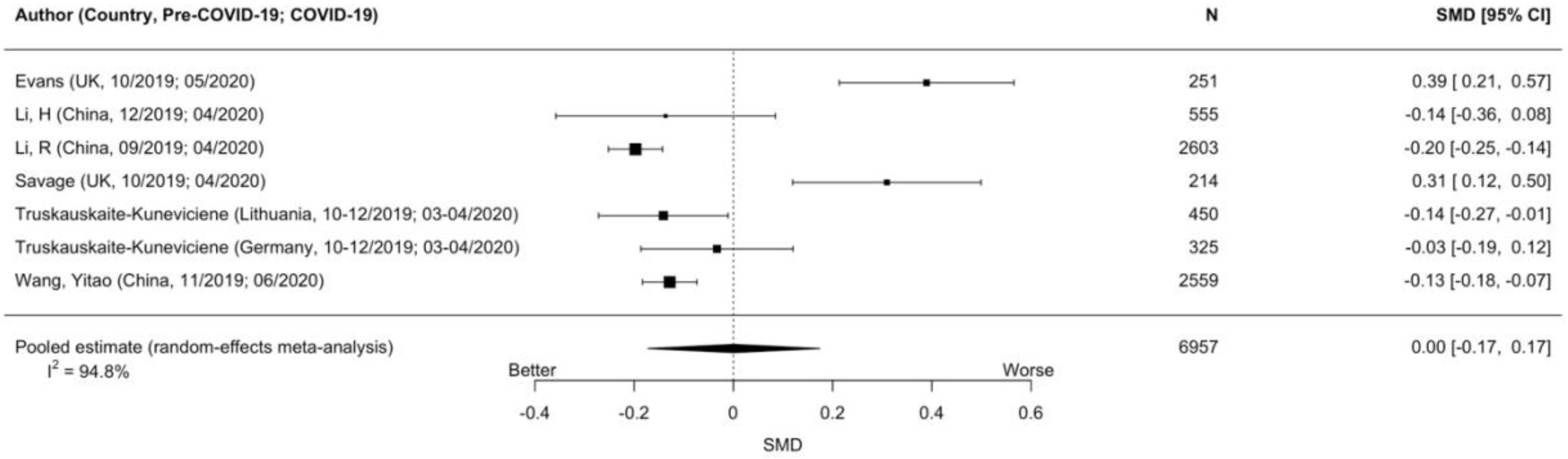
Forest plots of standardized mean difference changes in general mental health for studies of university students

**Figure 2g.**
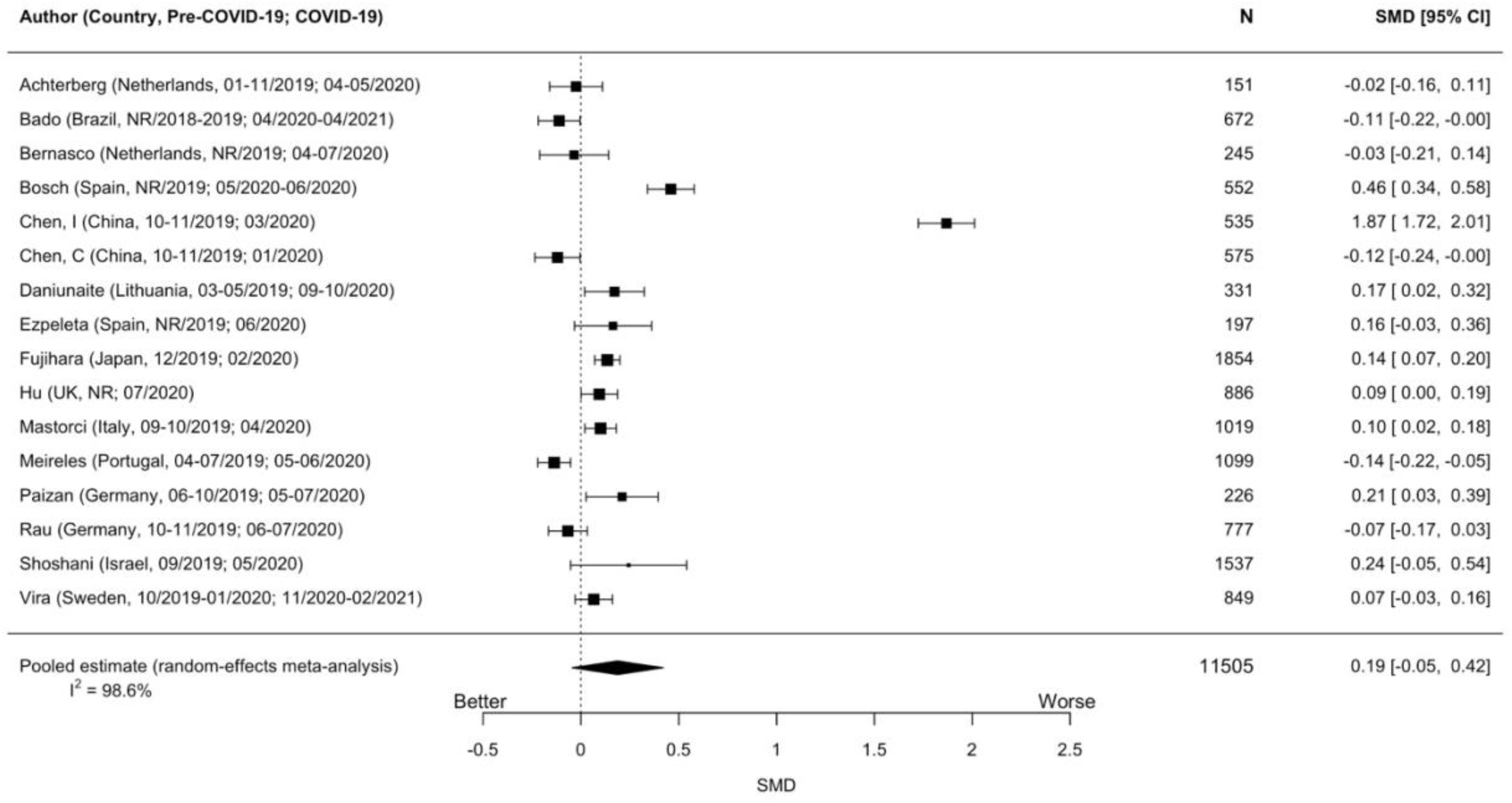
Forest plots of standardized mean difference changes in general mental health for studies of children and adolescents

**Figure 2h.**
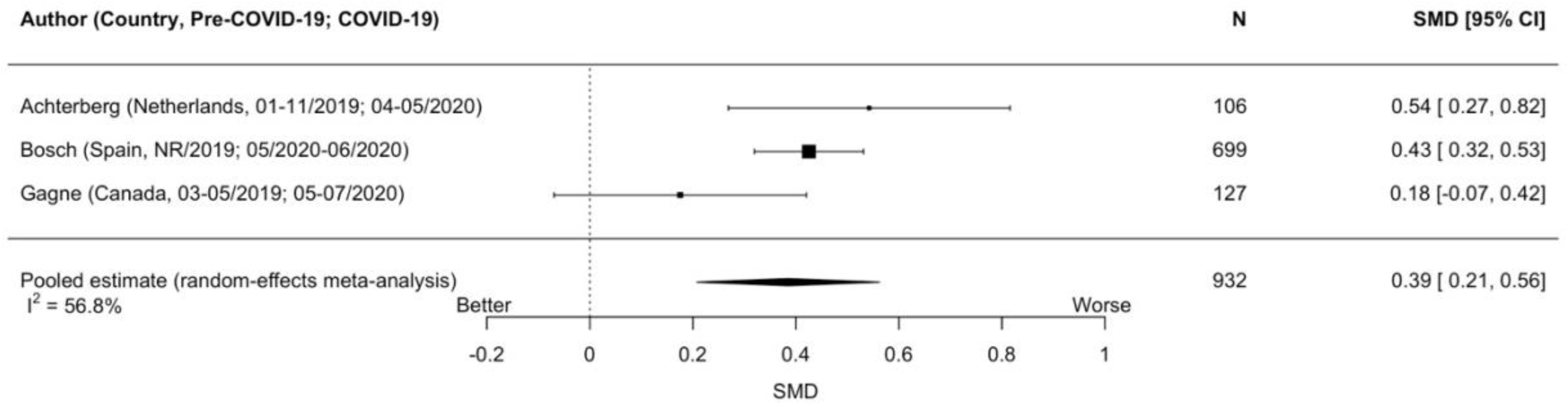
Forest plots of standardized mean difference changes in general mental health for studies of parents

**Figure 2i.**
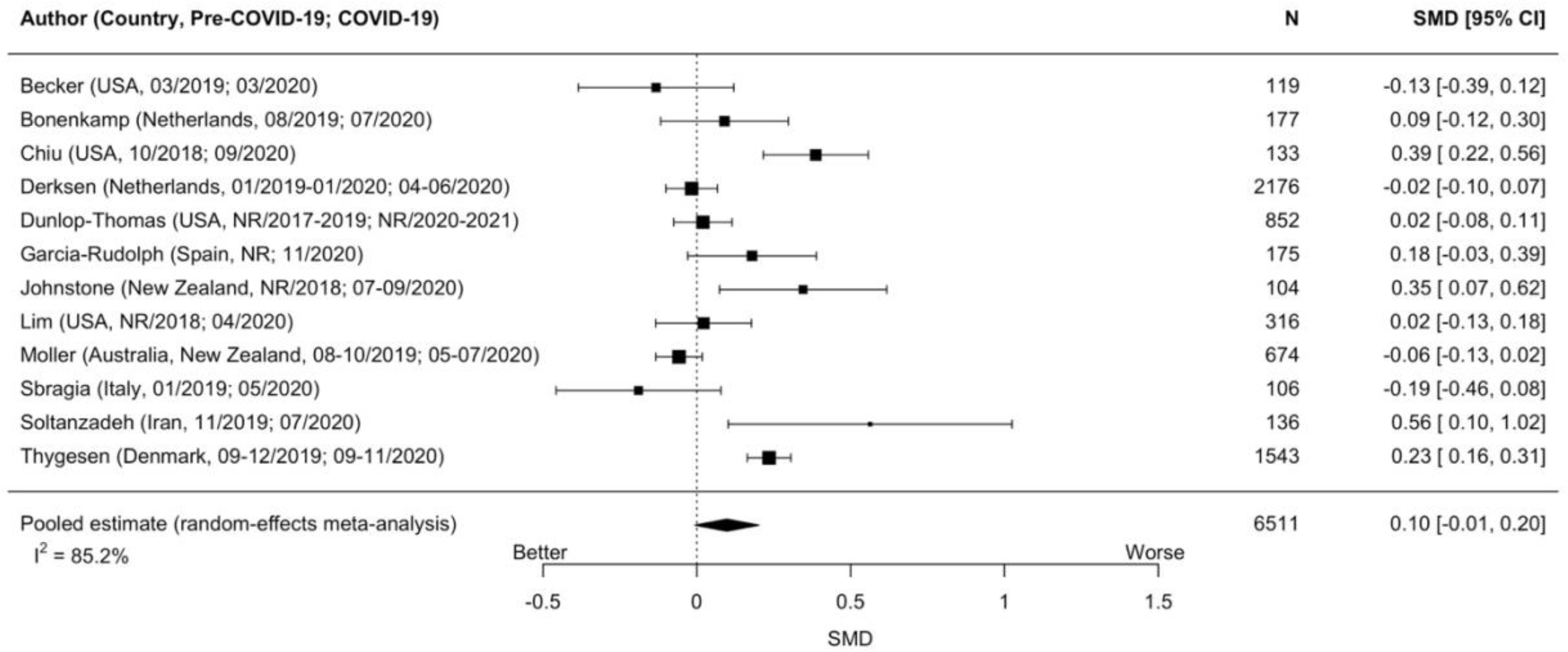
Forest plots of standardized mean difference changes in general mental health for studies of people with pre-existing medical conditions

**Figure 2j.**
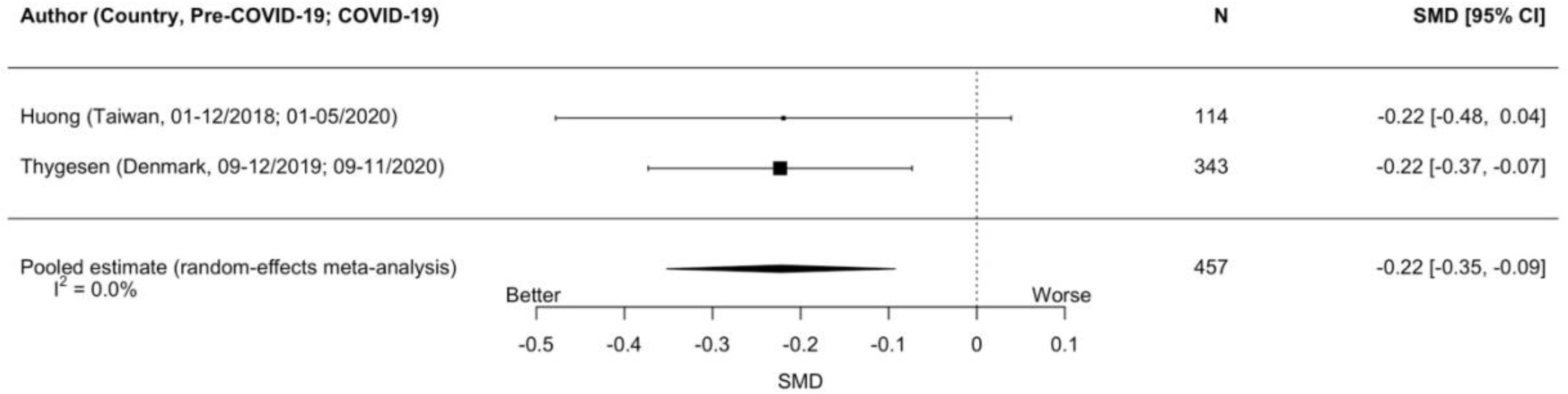
Forest plots of standardized mean difference changes in general mental health for studies of people with pre-existing mental health conditions

Results did not change from the main analysis for university students in a sensitivity analysis, in which outcomes for one study from April 2020^S63^ were replaced by a later measurement from October 2020^S64^ (see Supplementary Figure 1). Two large nationally sampled cohorts with continuous results from early 2020 reported dichotomous data from early and late 2020 but not continuous data for late 2020. Based on dichotomous data, the UK cohort saw an increase of 8.7% (95% CI 6.9% to 10.4%) of people with a GHQ-12 score of 4 or higher from pre-COVID-19 to April 2020, but this dissipated by September 2020 (change from pre-COVID-19 = 0.0%, 95% CI -2.0% to 1.9%).^S12^ Results were similar in that cohort for subgroups of women or females, and men or males, older adults, and young adults.^S12^ The general population cohort from the Netherlands, on the other hand, did not identify substantive changes from pre-COVID-19 in general mental health in either early or late 2020.^S16,S17,S33^

### Anxiety Symptoms

Forest plots for are shown in Figures 3a to 3j. Pooling of general population cohorts resulted in a non-statistically significant estimate of change in anxiety symptoms from pre-COVID-19 that was close to zero (Figure 3a; 4 cohorts, N = 2,632; SMD_change_ = 0.05, 95% CI - 0.04 to 0.13; I^2^ = 37%). Anxiety symptoms worsened statistically significantly by small amounts among women or females (Figure 3b; 5 cohorts, N = 3,500; SMD_change_ = 0.20, 95% CI 0.12 to 0.29; I^2^ = 41%) and parents (1 cohort, N = 147; SMD_change_ = 0.25, 95% CI 0.02 to 0.49). Estimates were non-statistically significant and close to zero for all other subgroups. I^2^ ranged from 0% to 41% for the general population, women or females, and men or males but was higher for all other subgroups (80% to 98%). For people with pre-existing medical conditions, results did not change in sensitivity analyses when data from September to October 2020 (Supplementary Figure 2) or March 2021 (Supplementary Figure 3) were substituted for results from early 2020 in one study with multiple assessments.^S118^

**Figure 3a.**
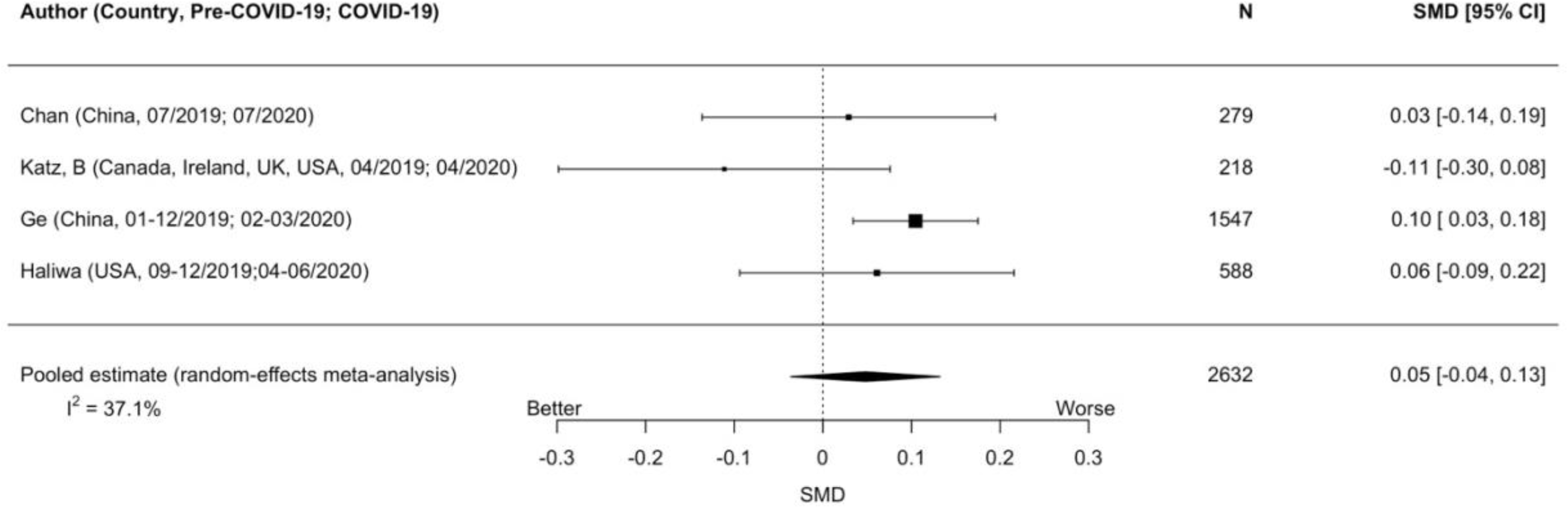
Forest plots of standardized mean difference changes in anxiety symptoms for studies of the general population

**Figure 3b.**
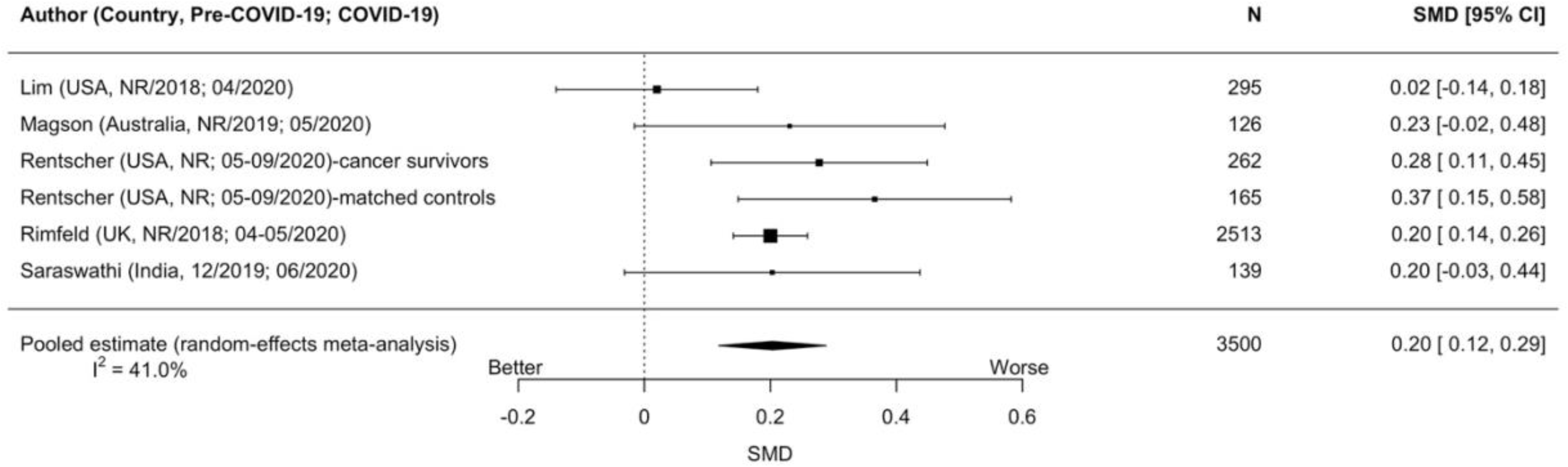
Forest plots of standardized mean difference changes in anxiety symptoms for studies of women or females

**Figure 3c.**
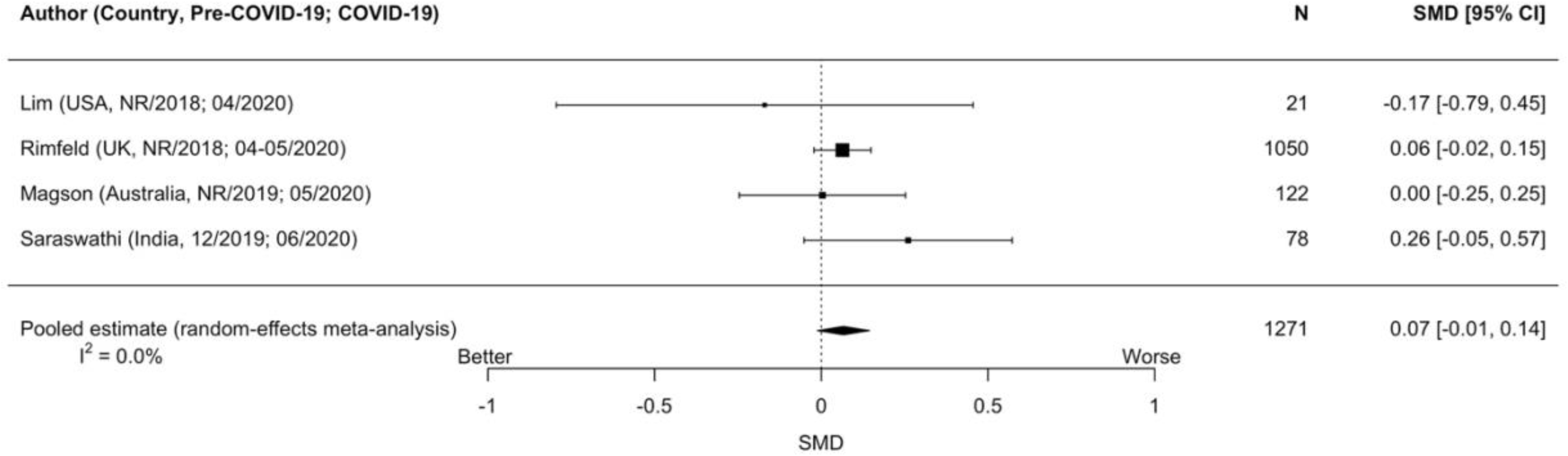
Forest plots of standardized mean difference changes in anxiety symptoms for studies of men or males

**Figure 3d.**
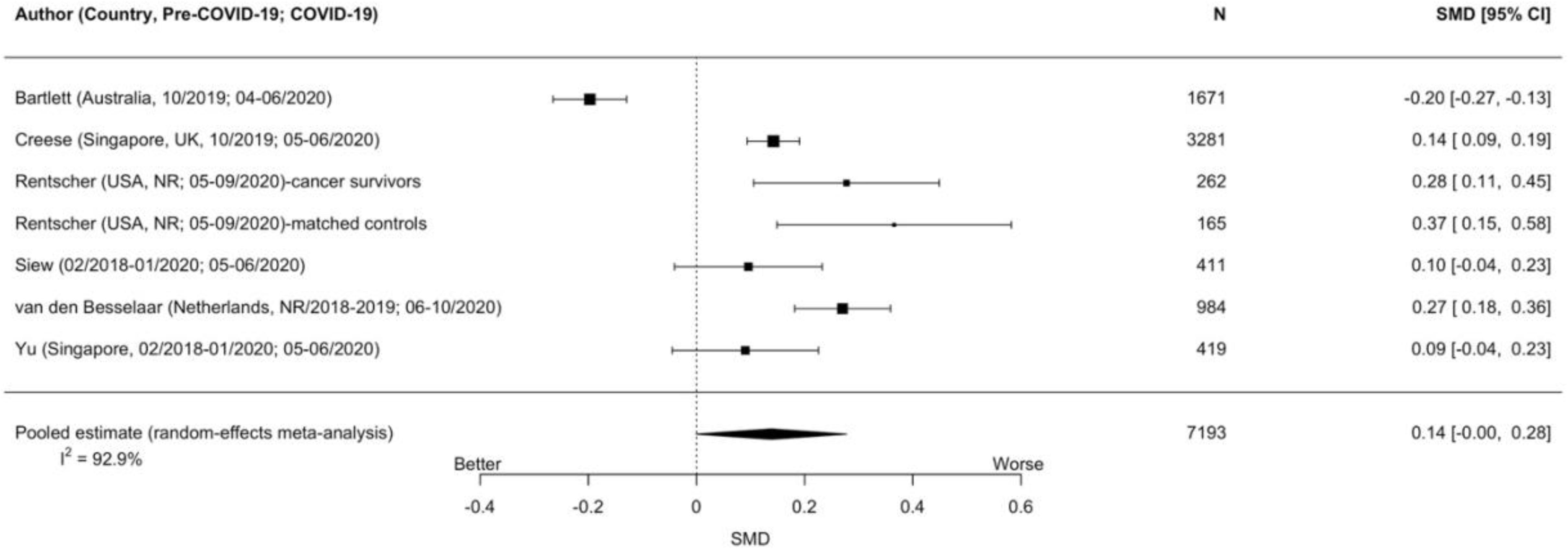
Forest plots of standardized mean difference changes in anxiety symptoms for studies of older adults

**Figure 3e.**
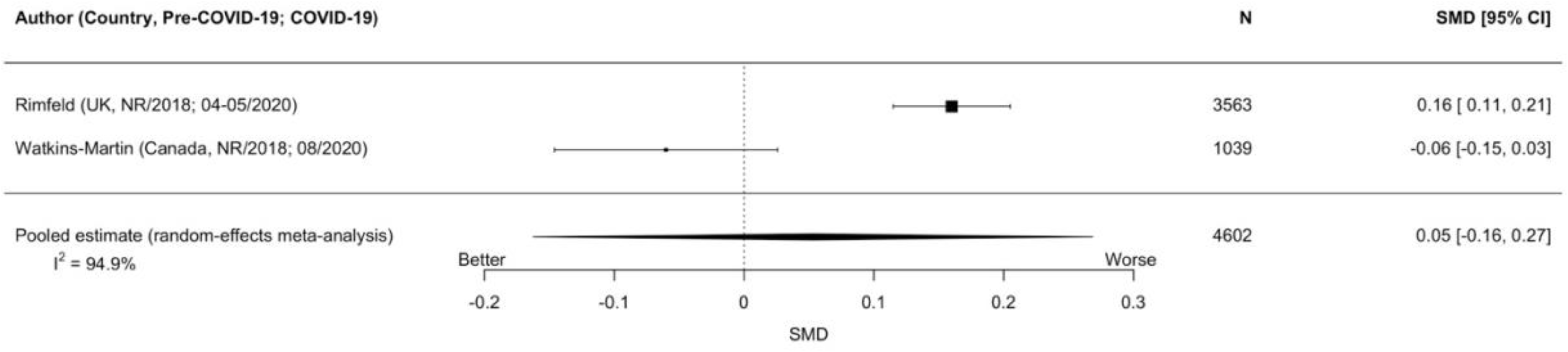
Forest plots of standardized mean difference changes in anxiety symptoms for studies of young adults

**Figure 3f.**
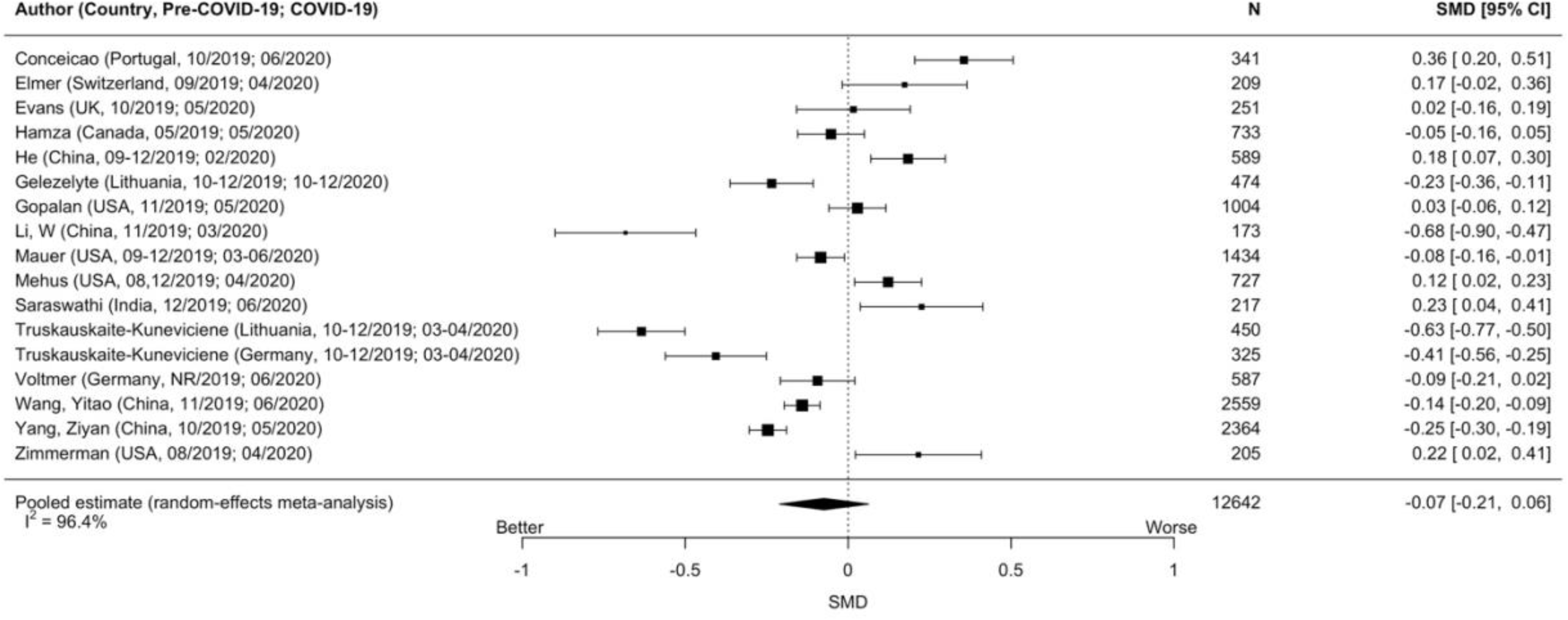
Forest plots of standardized mean difference changes in anxiety symptoms for studies of university students

**Figure 3g.**
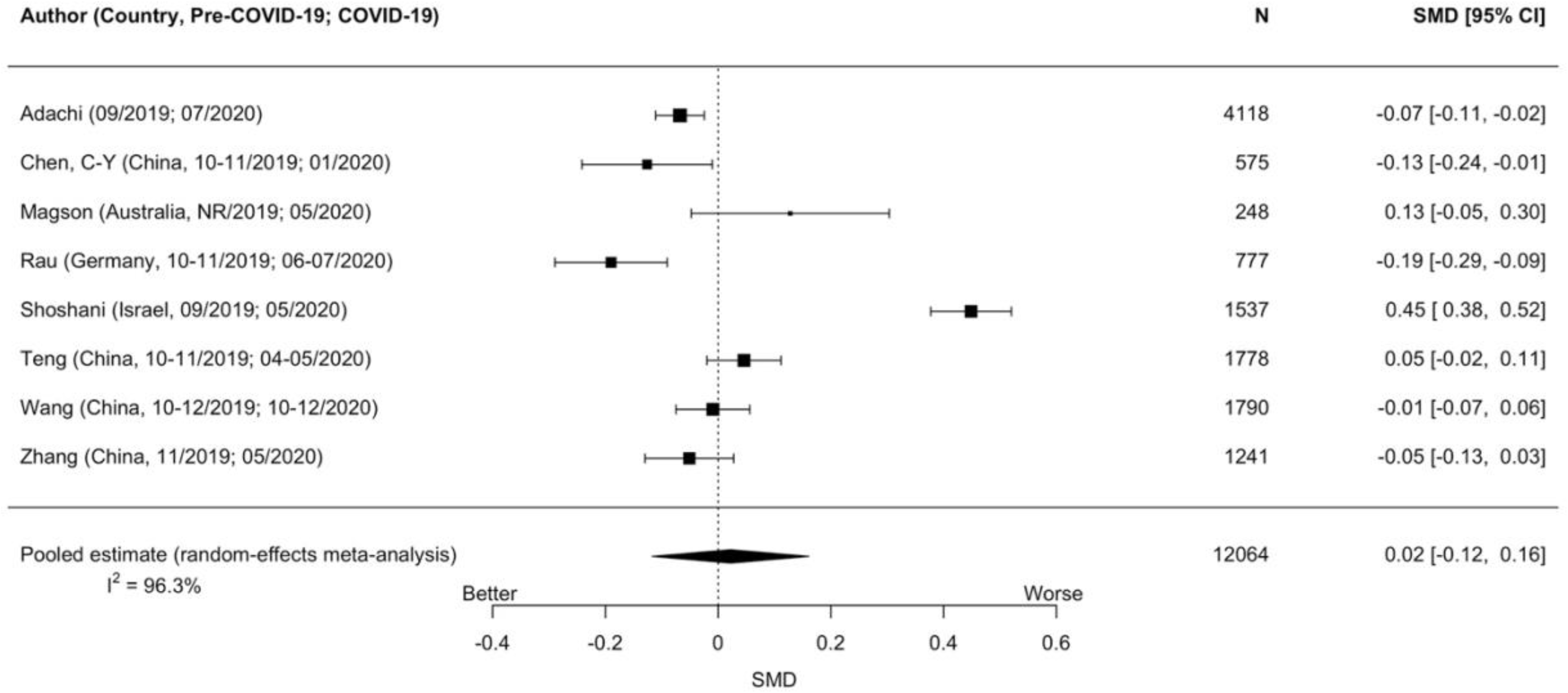
Forest plots of standardized mean difference changes in anxiety symptoms for studies of children and adolescents

**Figure 3h.**
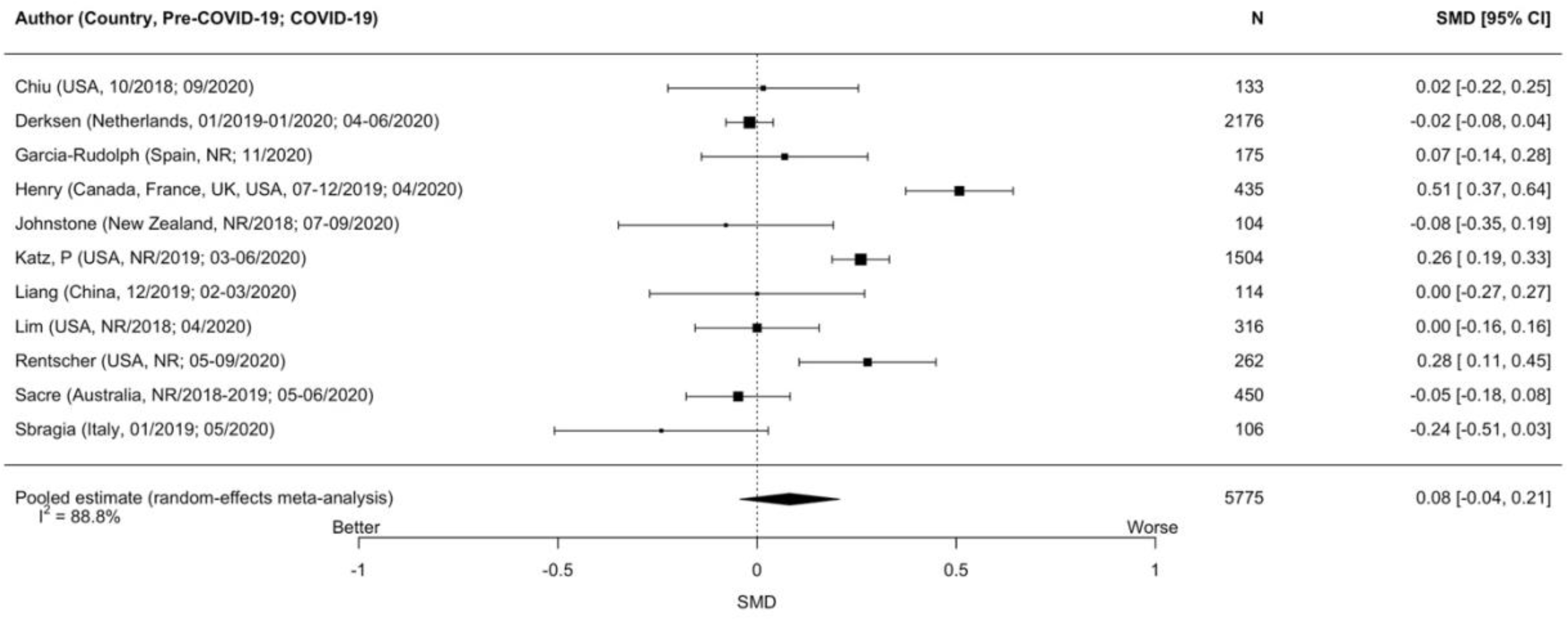
Forest plots of standardized mean difference changes in anxiety symptoms for studies of people with pre-existing medical conditions

**Figure 3i.**
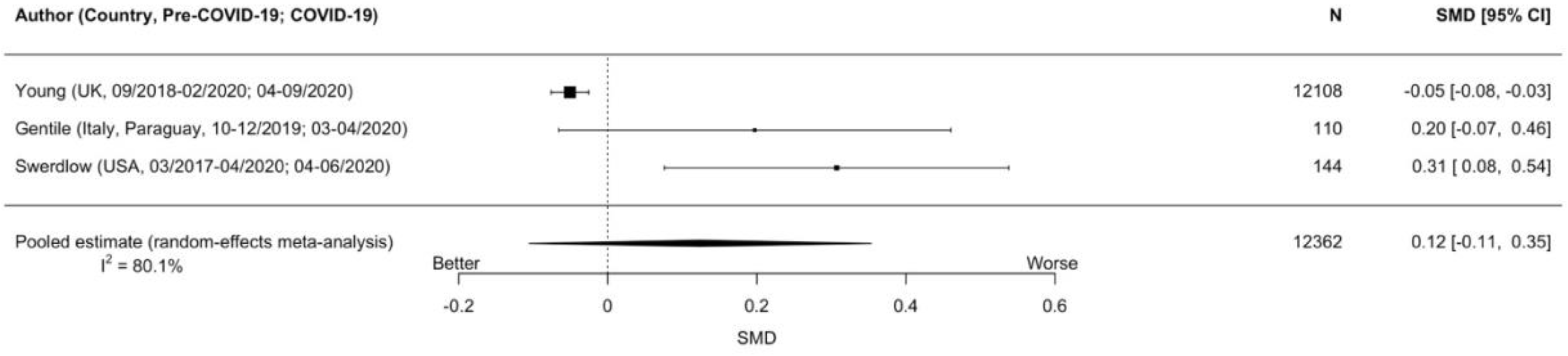
Forest plots of standardized mean difference changes in anxiety symptoms for studies of people with pre-existing mental health conditions

**Figure 3j.**
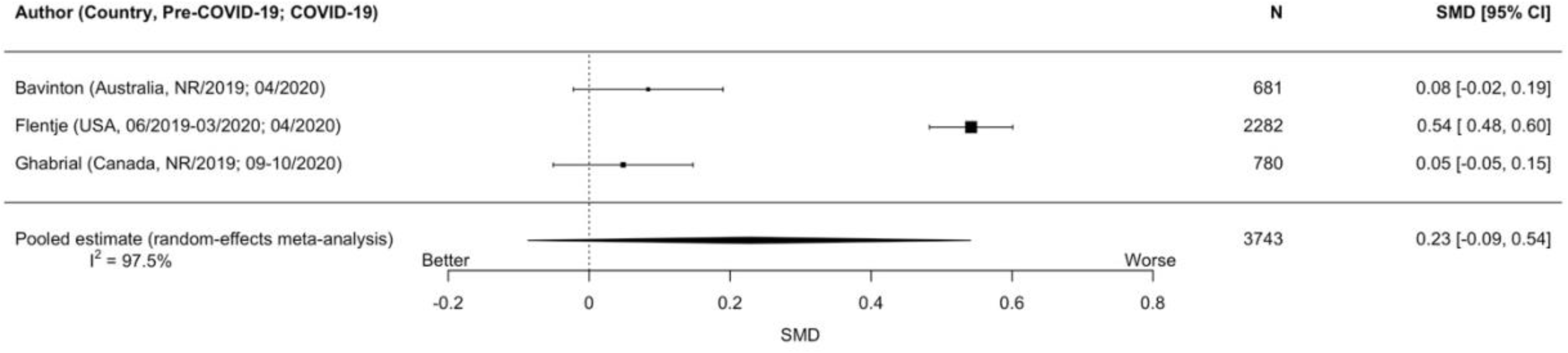
Forest plots of standardized mean difference changes in anxiety symptoms for studies of people who identify as sexual or gender minorities

### Depression Symptoms

Forest plots are shown in Figures 4a to 4k. In general population cohorts, symptoms of depression increased statistically significantly by a minimal amount (Figure 4a; 4 cohorts, N = 3,470; SMD_change_ = 0.12, 95% CI 0.01 to 0.24; I^2^ = 81%). They also increased significantly by minimal to small amounts among women or females (Figure 4b; 7 cohorts, N = 3,851; SMD_change_ = 0.22, 95% CI 0.05 to 0.40, I^2^ = 89%), older adults (Figure 4d; 7 cohorts, N = 7,419; SMD_change_ = 0.22, 95% CI 0.06 to 0.38, I^2^ = 95%), university students (Figure 4e; 19 cohorts, N = 26,164; SMD_change_ = 0.14, 95% CI 0.01 to 0.26, I^2^ = 98%), and people who identified as sexual or gender minorities (Figure 4k, 3 cohorts, N = 3,741; SMD_change_ = 0.19, 95% CI 0.10 to 0.28; I^2^ = 67%).

**Figure 4a.**
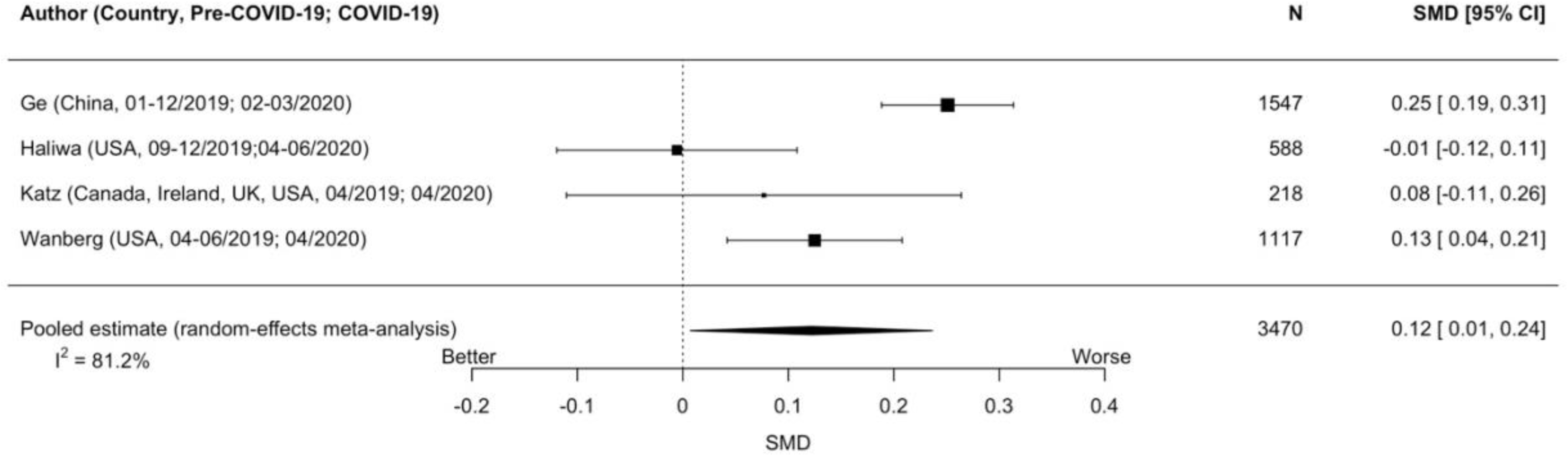
Forest plots of standardized mean difference changes in depression symptoms for studies of the general population

**Figure 4b.**
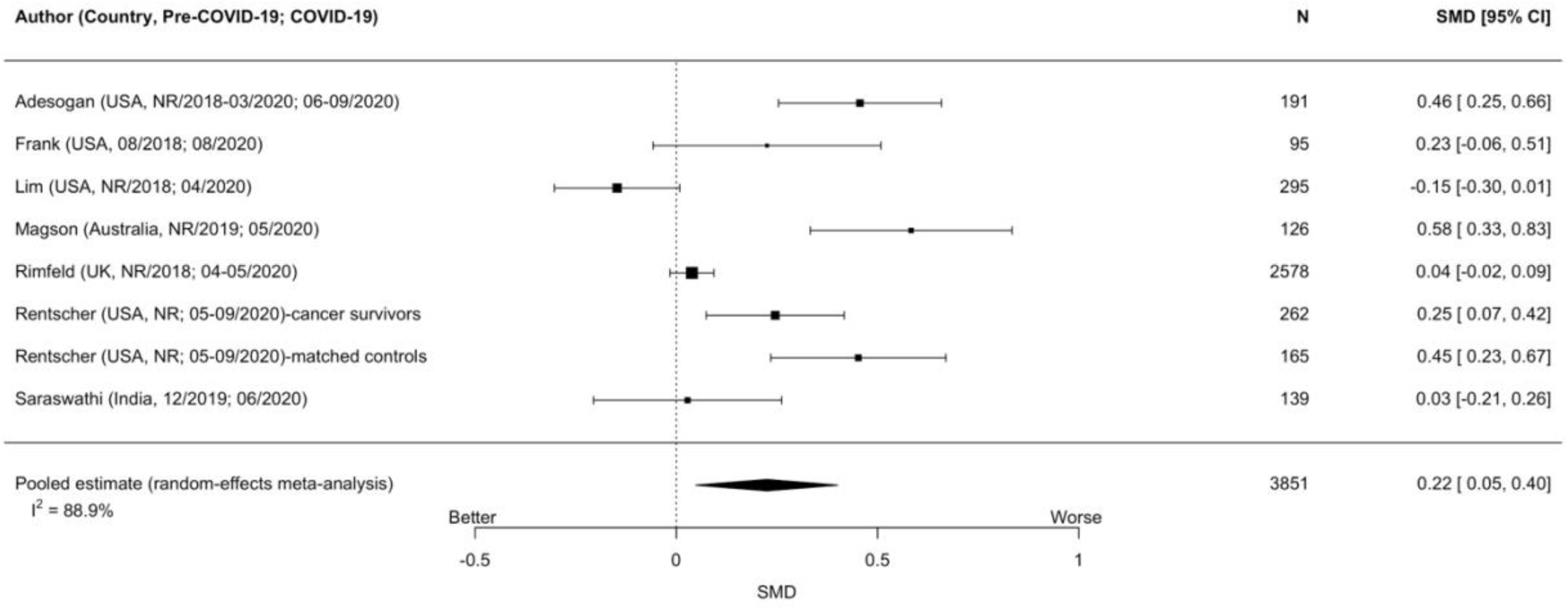
Forest plots of standardized mean difference changes in depression symptoms for studies of women or females

**Figure 4c.**
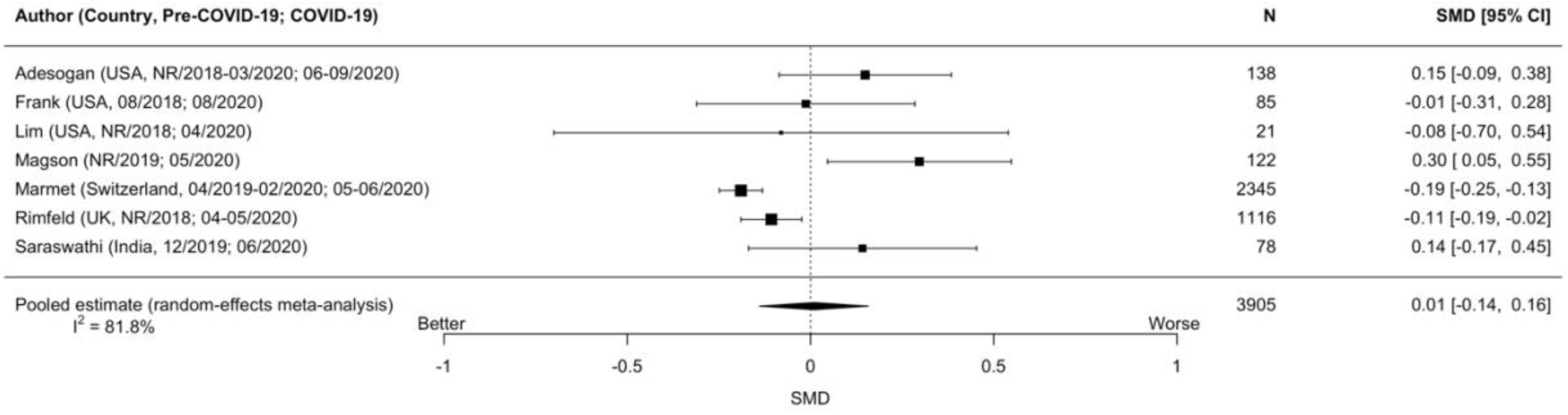
Forest plots of standardized mean difference changes in depression symptoms for studies of men or males

**Figure 4d.**
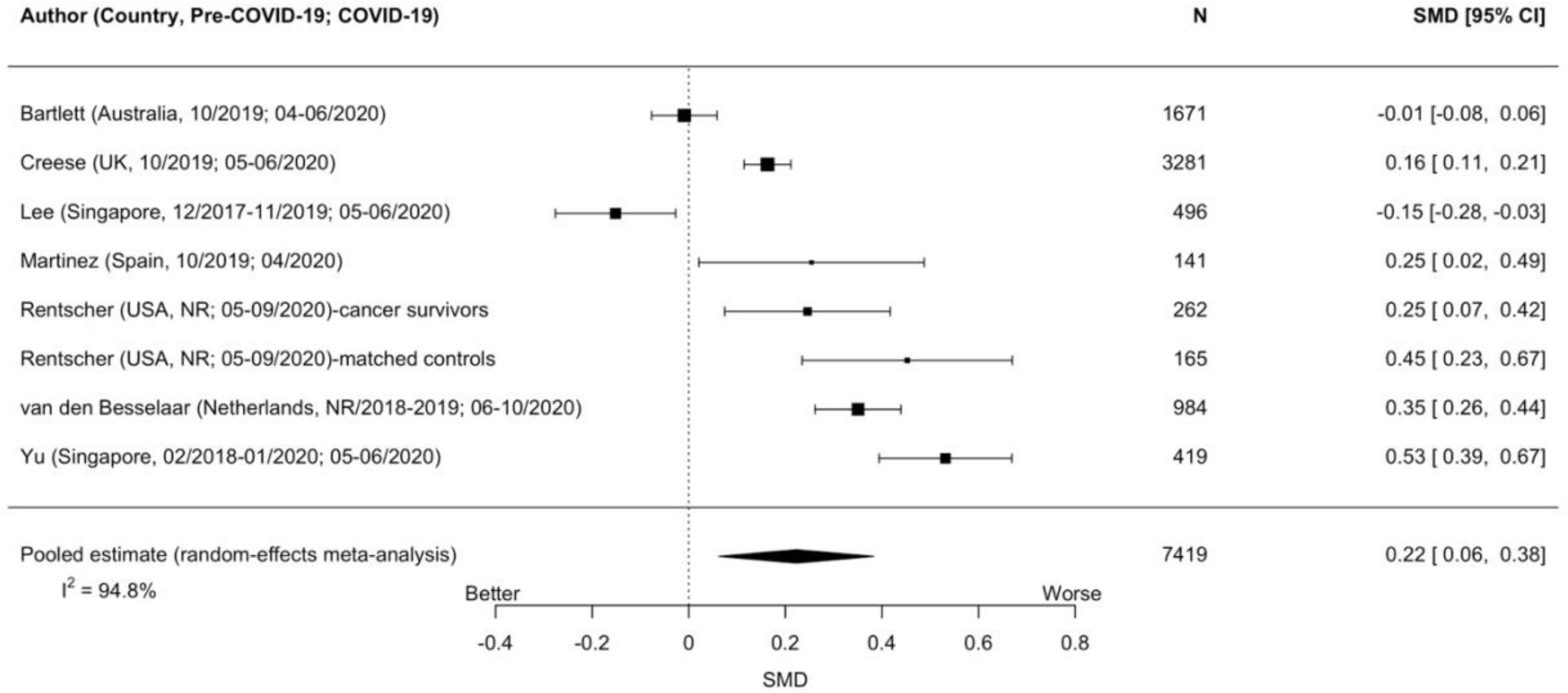
Forest plots of standardized mean difference changes in depression symptoms for studies of older adults

**Figure 4e.**
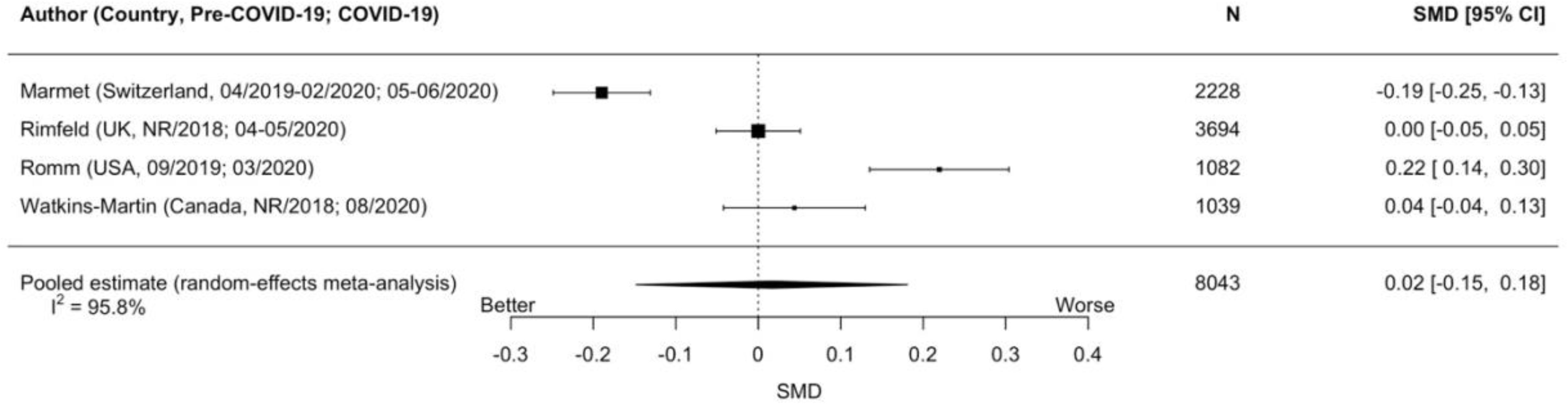
Forest plots of standardized mean difference changes in depression symptoms for studies of young adults

**Figure 4f.**
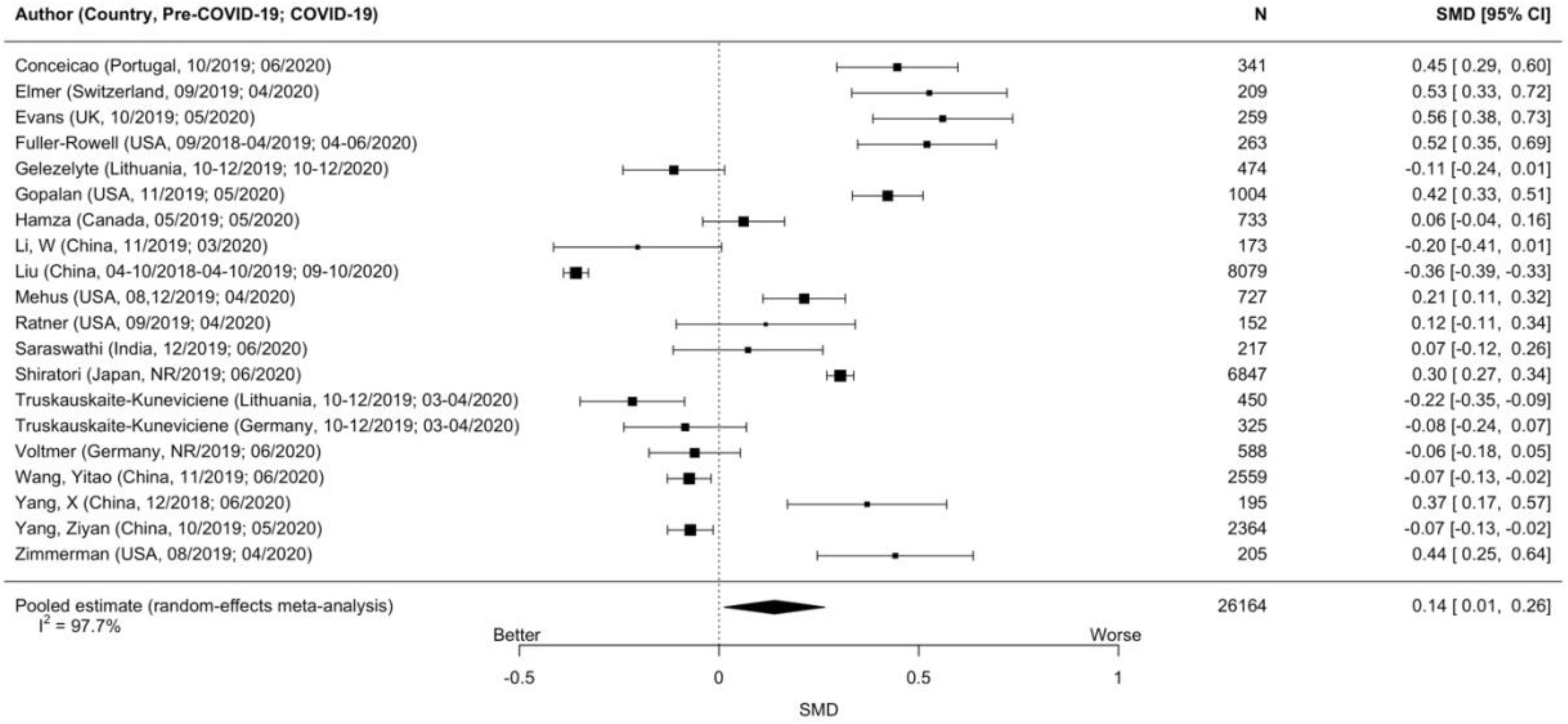
Forest plots of standardized mean difference changes in depression symptoms for studies of university students

**Figure 4g.**
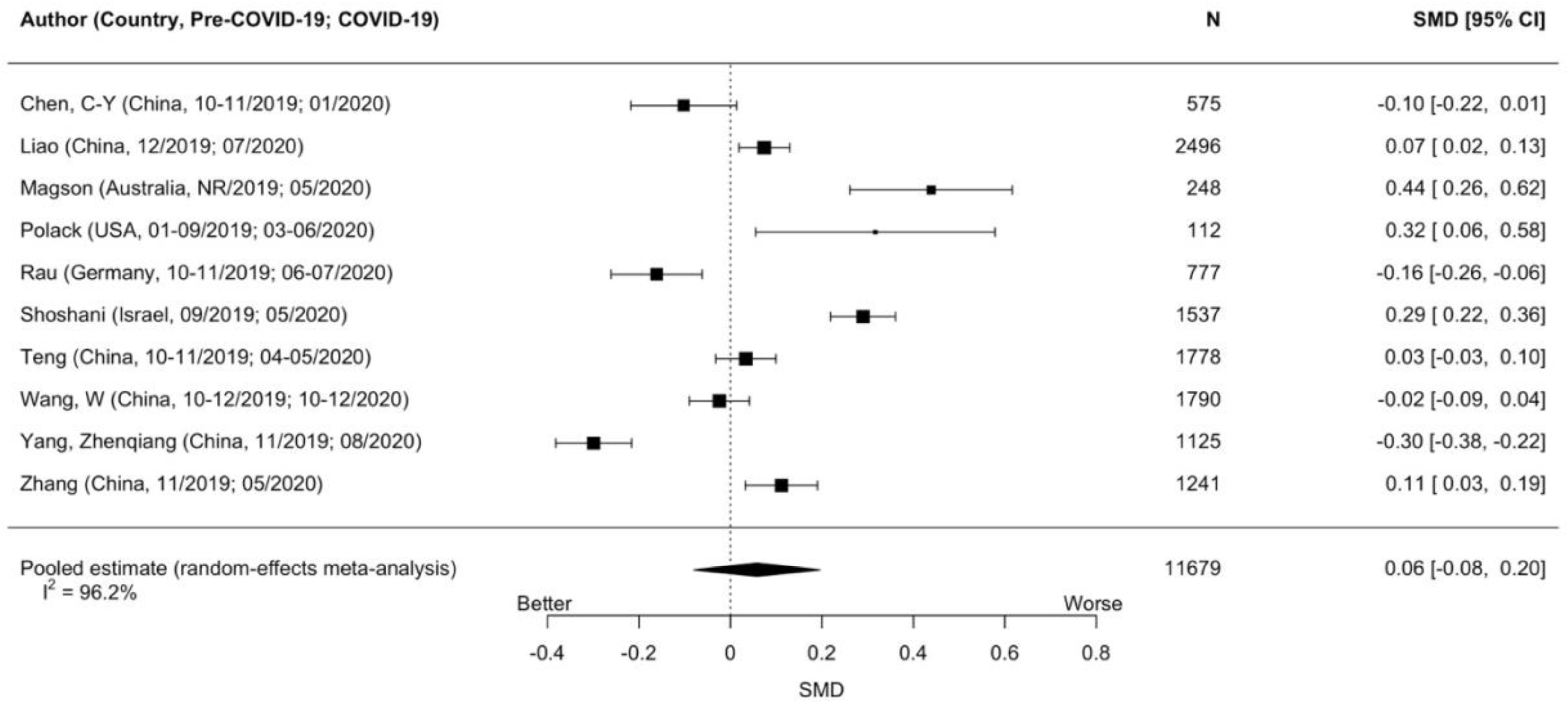
Forest plots of standardized mean difference changes in depression symptoms for studies of children and adolescents

**Figure 4h.**
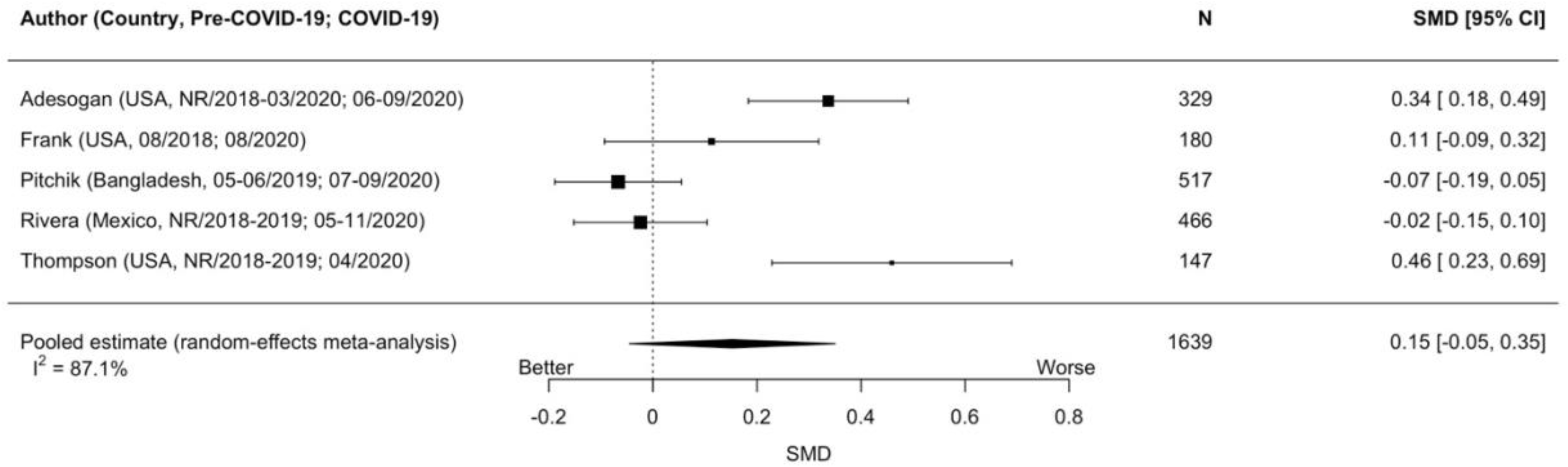
Forest plots of standardized mean difference changes in depression symptoms for studies of parents

**Figure 4i.**
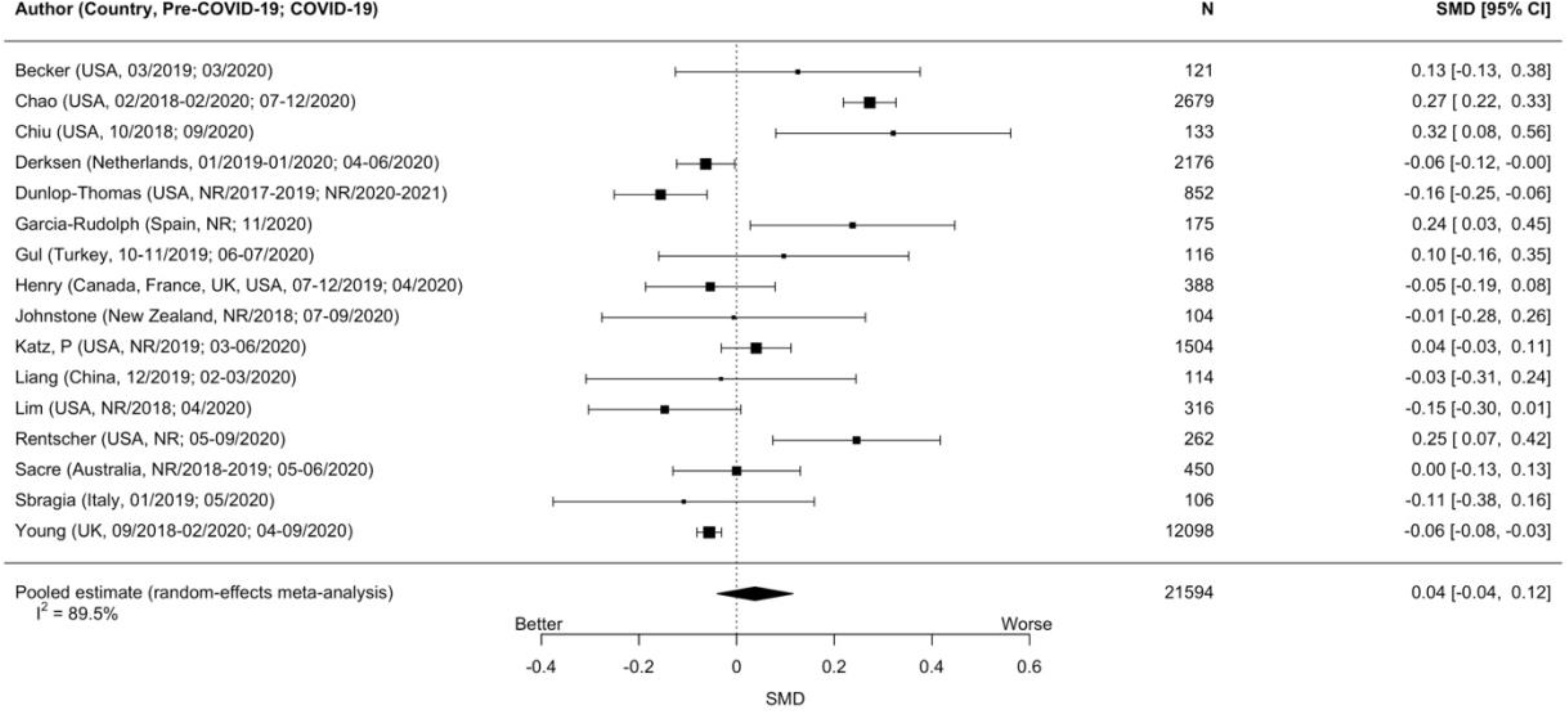
Forest plots of standardized mean difference changes in depression symptoms for studies of people with pre-existing medical conditions

**Figure 4j.**
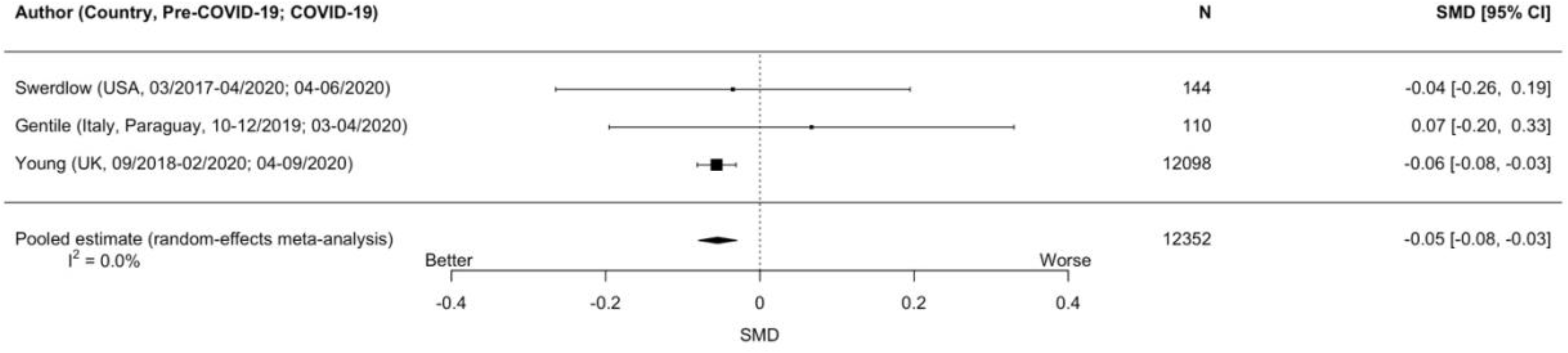
Forest plots of standardized mean difference changes in depression symptoms for studies of people with pre-existing mental health conditions

**Figure 4k.**
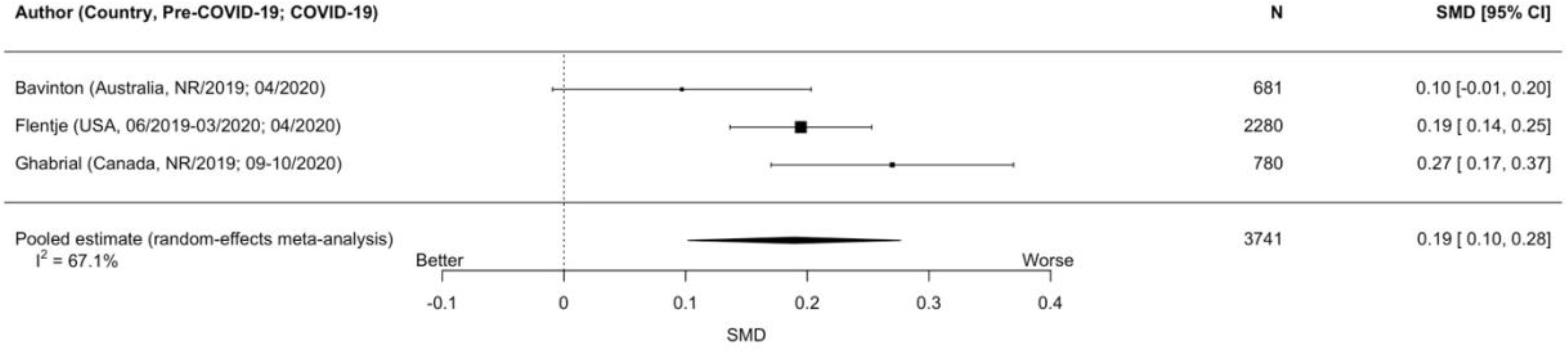
Forest plots of standardized mean difference changes in depression symptoms for studies of people who identify as sexual or gender minorities

They improved minimally for people with pre-existing mental health conditions (Figure 4j, 3 cohorts, N = 12,352; SMD_change_ = -0.05, 95% CI -0.08 to -0.03; I^2^ = 0%). I^2^ was 0% for people with pre-existing mental health conditions and 67% to 98% in all other analyses. Results did not change for people with pre-existing medical conditions in two sensitivity analyses (Supplementary Figures 4 and 5).

## DISCUSSION

### Principal Findings

We reviewed over 94,000 citations and included 137 studies from 134 cohorts that compared mental health during COVID-19 to assessments done prior to COVID-19. Included studies assessed changes in general mental health, anxiety symptoms, and depression symptoms in general population samples and among women or females, men or males, older adults, young adults, university students, children and adolescents, parents, people with pre-existing medical conditions, people with pre-existing mental health conditions, medical staff, and people who identified as sexual or gender minority individuals. All studies assessed COVID-19 symptoms during at least one time point in 2020, which in most cases was in the first half of the year. Only large population-based cohorts from the United Kingdom^S11,S12^ and the Netherlands^S16,S17,S33^ and a cohort of people with the rare autoimmune disease systemic sclerosis from 4 countries^S118^ assessed symptoms in both April 2020 and late 2020; the systemic sclerosis cohort also reported monthly results up to March 2021.

There was substantial heterogeneity in many of the general population and subgroup analyses that we conducted, which suggests that the point estimates in these meta-analyses should be interpreted cautiously. There was, however, general consistency across analyses in that most estimates of symptom changes were close to zero and not statistically significant, and changes that were identified were of minimal to small magnitudes. Among general population studies, we did not find changes in general mental health or anxiety symptoms, although depression symptoms worsened minimally.

Among subgroups, only women or females experienced consistent symptom worsening across outcome domains, all by small amounts (SMD_change_ = 0.20 to 0.22). Depression symptoms also worsened by minimal to small amounts for older adults, university students, and sexual or gender minorities but not for other groups. General mental health (3 studies, N = 932) and anxiety symptoms (1 study, 147 participants) worsened for parents, but this was based on very small numbers of studies and participants. General mental health and depression symptoms improved for people with pre-existing mental health conditions, but these findings were based on only 2 studies (N = 457) for general mental health, and improvement was negligible, even though statistically significant, for depression symptoms (SMD_change_ = 0.05).

There were 3 cohorts with analyses that assessed changes separately in early 2020 (March to June) and later 2020 (September to November).^S11,S12,S16,S17,S33,S118^ In a large Dutch general population sample (N = 3,983 to 4,064), change in general mental health was close to zero at both time points, and this was also the case for subgroups (women or females, men or males, older adults, younger adults).^S16,S17,S33^ In a United Kingdom general population study, there was a small worsening from pre-COVID-19 to April 2020 based on continuous^S11^ and dichotomous^S12^ results, but dichotomous results from September 2020 were not different from pre-COVID-19.^S12^ Similarly, in a cohort of people with systemic sclerosis,^S118^ anxiety symptoms were substantially higher in April 2020 compared to pre-COVID-19, but they were close to pre-COVID-19 levels in September to October 2020 and in March 2021.

## Findings in Context

The main finding that mental health was either largely unchanged or worsened by minimal to small amounts in the general population and subgroups is consistent with results from a more limited systematic review of 65 longitudinal studies from early in the pandemic.^17^ We know of only one study that has evaluated mental disorders using valid diagnostic methods. That study, from Norway,^32^ which was not eligible for our review, evaluated current mental disorders using the Composite International Diagnostic Interview (version 5.0) in a series of cross-sectional random samples accumulated from January 28 to March 11, 2020 (N = 563, 15.4%, 95% CI 12.5% to 18.8%), March 12 to May 31, 2020 (N = 691, 9.0%, 95% CI 7.1% to 11.4%), June 1 to July 31, 2020 (N = 530, 14.3%, 95% CI 11.5% to 17.5%), and August 1 to September 18, 2020 (N = 370, 11.9%, 95% CI 9.0% to 15.6%). The authors concluded that prevalence of mental health disorders during COVID-19, compared to January to March 2020, which they considered pre-COVID-19, was stable or slightly decreased.

The authors of the largest study to date on suicide in COVID-19^33,34^ initially analysed data from official government sources on suicide occurences at a monthly level from January 1, 2019 or earlier to July 31, 2020. They used an interrupted time-series analysis to model trends in monthly suicides before COVID-19 (January 1, 2019 or earlier to March 31, 2020) and compared the expected number from the model with the observed number of suicides from April 1 to July 31, 2020 for data from 21 countries. There was no evidence of a statistically significant increase in suicide risk in any country or area; there were, however, statistically significant decreases in 12 countries or areas.^32^ A subsequent update of 33 countries across the first 9 to 15 months of the pandemic found similar results.^33^

We found that women or females consistently experienced small negative changes, in aggregate, during the early part of the pandemic across general mental health, anxiety symptoms, and depression symptoms. This finding is consistent with a previous analysis of a subset of studies from those in the present review with direct comparisons between mental health of women or females and men or males (N = 10 unique cohorts).^35^ That study found significantly, albeit minimally, greater worsening of general mental health and anxiety symptoms among women (SMD_change_ = 0.15 for both); although, depressive symptoms were not significantly different but were worse for women (SMD_change_ = 0.12).^35^ Significant worsening of symptoms among women or females, although by a small amount, is of concern. This is an aggregate result, and even though it is small, it suggests that the pandemic has likely impacted some women or females substantively. By gender, the pandemic has arguably disproportionately affected women. In terms of vulnerabilities, most single-parent families are headed by women, and women earn less and are more likely to live in poverty than men.^36–40^ They are overrepresented in health care jobs and provide most family and elder care.^36–39^ Intimate partner violence incurred by women has increased during the pandemic.^41^ The small overall change in symptoms that we detected suggests that many women have been resilient to difficult circumstances but that there has likely been important symptom worsening among some. Indeed, although most of our analyses found no changes or minimal to small negative changes in mental health, they do suggest that the pandemic has taken a toll on some people, which is consistent with reports of increased visits for mental health and sustance use, for instance.^42, 43^

Nonetheless, the patterns of findings on mental health symptoms from our review, along with evidence on mental disorders and suicide, converge to suggest that the effects of COVID-19 on mental health are more nuanced than what had been described as a “tsunami” or other similar terms in many mainstream media articles.^15^ Short news cycles that emphasize dramatic events, anecdotes, and an uncritical reliance on unvalidated, difficult to interpret survey tools that inquire about mental health and well-being in COVID-19 among conveniently recruited volunteers might at least partially explain this discrepancy. Illustrating the pitfalls of interpreting studies that ask about COVID-19-specific angst, a study of 2,345 young men from Switzerland^44^ evaluated depression symptoms and stress during COVID-19 and found that they had significantly decreased compared to pre-pandemic levels. They also reported results from a series of unvalidated single items that queried about psychological status during COVID-19 and specifically assigned COVID-19 as the cause (e.g., “due to COVID-19, I experienced…”); these items suggested very high levels of distress, which became the focus of the study’s conclusions. Together with the findings from our systematic review, this suggests that many or most people are likely experiencing different aspects of COVID-19 as highly unpleasant or distressing, but that many people have been resilient and that evidence does not support the idea that population-level mental health has changed by large amounts, although it has changed negatively for some.

## Implications

The lack of evidence of a large-scale decline in mental health so far in COVID-19 could be because people are resilient and have made the best of a difficult situation. Indeed, although evidence is thin in this area, there are data to suggest, for instance, that suicide has generally declined during periods of societal conflict.^45–49^ War and pandemics have very different characteristics, but in both there is a shared threat and common focus on collective action to address that threat.

The lack of changes or small changes in mental health could also reflect steps that governments around the world have taken to support mental health. The World Health Organization, other pan-national organizations, and governments across the globe have produced strategies for addressing mental health and have invested in resources to support public mental health,^50, 51^ even in countries where mental health had not been a priority previously.^52, 53^ It is not known to what degree these efforts have been effective, but it is possible that government action has played an important role. On the other hand, the negative changes that we detected in some groups, particularly for women and females, early in the pandemic, underline the need for continued surveillance to determine the degree to which negative mental health changes may be ongoing and will require additional resources to address.

In terms of research, the results of our systematic review underline gaps in mental health surveillance across countries. Since early in the pandemic, the need for high-quality surveys with appropriately representative probabilistic sampling methods and pre-pandemic data has been emphasized.^54^ We found, however, few examples of mental health surveillance frameworks that generated high-quality data based on this type of sampling. As the authors of an editorial on this topic emphasized, “We can and must do better” (p. 568).^54^ This will require that governments invest in ongoing mental health surveillance mechanisms that can be used to improve our ability to address mental health needs at all times, as well as in times of crisis.This should include designs that allow us to identify patient-level factors that may influence risk of mental health changes.

## Strengths and Limitations

Strengths of our systematic review include using rigorous best-practice methods; searching 9 databases, including 2 Chinese databases; not restricting inclusion by language; and the ability to update rapidly as evidence emerges via our living systematic review approach. There are also limitations that suggest some level of caution in interpreting results. First, we did not peer review our search strategy given the urgency of generating synthesized mental health data early in the pandemic. However, the experienced librarian who developed the search strategy developed COVID-19 search terms in collaboration with other experienced librarians.

We reviewed studies included in other published reviews and did not identify any missed studies; to the contrary, our review identified more studies than in other similar reviews.^17^ Second, aside from several population-level randomly sampled surveys, most of the studies included in our systematic review had limitations related to study sampling frames and recruitment methods, response and follow-up rates, and management of missing follow-up data. Third, heterogeneity was high in most of the meta-analyses that we conducted. Heterogeneity of populations, conditions when they were conducted, along with generally low study quality and reporting did not allow us to evaluate factors that may have been associated with symptom change patterns. Fourth, only a handful of studies reported results from the fall months of 2020, and, although the few studies that did suggested that symptoms were stable or reduced from earlier in the pandemic, more data are needed. Fifth, although we were able to synthesize results from several vulnerable groups, including older adults and people with pre-existing medical conditions, there were few studies with results for other vulnerable groups, including people with low socio-economic status. It is possible that some groups may be experiencing important negative mental health effects of the pandemic and were not included in the studies we identified. Similarly, there was little evidence from low- or lower-middle-income countries or from some areas of the world, such as sub-Saharan Africa. Sixth, some potentially important outcomes, such as loneliness, were infrequently studied and not included in the present report. Seventh, we did not include studies with < 100 participants for feasibility, but we do not believe that this would have influenced results meaningfully. In a smaller systematic review with 65 total studies that did include such studies,^17^ studies with < 100 participants comprised only 1% of total participants from included studies. Eighth, the evidence base is rapidly evolving, and main results could change, although our living systematic review format will allow rapid updating as this occurs. Ninth, although we found some groups with minimal to small worsening in mental health, it is possible that this could have occurred over time regardless of the pandemic or that this was due to the combination of studies that happened to sample from certain groups. We do believe, however, that the consistent findings of no changes to minimal changes in the general population but small changes for women or females across domains are likely robust despite this possibility. Tenth, we did not assess for possible publication bias, though the largely null findings suggest that this was not likely an important factor.

## Conclusions

We reviewed 137 studies with data from 134 unique cohorts. Across population groups, results suggest that, rather than a mental health crisis, at a population level, there has been a high level of resilience during COVID-19, and changes in general mental health, anxiety symptoms, and depressive symptoms have been minimal to small with no changes detected in many analyses. There were few robust studies with vulnerable groups, however, and it is possible that there are population groups that are experiencing a different level of mental health effect than the general population or other groups. COVID-19 and long-term ramifications of the pandemic continue to affect societies across the world, and it will be important to continue to assess mental health. There were few studies with data beyond the initial months of the pandemic, and the negative changes we found in some groups, as well as economic and other ongoing burdens from the pandemic warrant continued mental health surveillance. The pandemic has upended the lives of many people around the world, and there is little doubt that some people who have not experienced mental health difficulties previously are experiencing them now. Governments should continue to ensure that mental health supports are available and respond to population needs. We will continue to review the COVID-19 mental health evidence base, as studies continue to be produced, with all study results posted online (https://www.depressd.ca/covid-19-mental-health) and produce future reports when more data at later points in the pandemic are available.

## Contributions

YS, YWu, DBR, AB, and BDT were responsible for the study conception and design. JTB was responsible for the design of the database searches. KL and AK carried out the searches. YS, YWu, SF, TDS, LL, XJ, KL, YWang, AT, AK, CH, OB, DBR, SM, MA, ITV, DN, EW, AY, and BDT contributed to data extraction, coding, and evaluation of included studies. YS and SF were responsible for study coordination. YS, YWu, BL, AB, and BDT were involved in data analysis. BA, CF, MSM, SS, and GT contributed to interpretation of results as knowledge translation partners. YS and BDT drafted the manuscript. All authors provided a critical review and approved the final manuscript. BDT is the guarantor; he had full access to all the data in the study and takes responsibility for the integrity of the data and the accuracy of the data analyses. BDT is the corresponding author and attests that all listed authors meet authorship criteria and that no others meeting the criteria have been omitted.

## Copyright for Authors

The Corresponding Author has the right to grant on behalf of all authors and does grant on behalf of all authors, to the Publishers and its licensees in perpetuity, in all forms, formats and media (whether known now or created in the future), to i) publish, reproduce, distribute, display and store the Contribution, ii) translate the Contribution into other languages, create adaptations, reprints, include within collections and create summaries, extracts and/or, abstracts of the Contribution, iii) create any other derivative work(s) based on the Contribution, iv) to exploit all subsidiary rights in the Contribution, v) the inclusion of electronic links from the Contribution to third party material where-ever it may be located; and, vi) licence any third party to do any or all of the above.

The Corresponding Author has the right to grant on behalf of all authors and does grant on behalf of all authors, an exclusive licence (or non-exclusive for government employees) on a worldwide basis to the BMJ Publishing Group Ltd to permit this article (if accepted) to be published in BMJ editions and any other BMJPGL products and sublicences such use and exploit all subsidiary rights, as set out in our licence.

## Ethical Approval

As this study was a systematic review of published results, it did not require ethical approval.

## Transparency Declaration

The manuscript’s guarantor affirms that the manuscript is an honest, accurate, and transparent account of the study being reported; that no important aspects of the study have been omitted; and that any discrepancies from the study as planned (and, if relevant, registered) have been explained.

## Data Sharing

All data used in the study are available in the manuscript and its tables or on online at https://www.depressd.ca/covid-19-mental-health.

## Dissemination to study participants or patient communities

There are no plans to disseminate the results of the research directly to a relevant patient community. However, all study results are available at https://www.depressd.ca/research-question-1-symptom-changes.

## Provenance and peer review

Not commissioned; externally peer reviewed.

## CC BY licence

The default licence, a CC BY NC licence, is needed.

## What is already known on this topic

- Large numbers of studies and media reports have concluded that COVID-19 has led to widespread decline in population mental health.
- Most existing evidence reviews have been based on cross-sectional studies and conclusions based on proportions of study respondents above thresholds on mental health measures, which are not intended for this purpose and can be highly misleading.

## What this study adds

- We synthesized evidence from 137 studies that compared general mental health, anxiety symptoms, or depression symptoms during COVID-19 to outcomes prior to COVID-19 in the same participant cohort.
- We did not identify negative changes in mental health at the general population level for general mental health or anxiety symptoms, but we did find a minimal worsening of depression symptoms.
- Among subgroups, women and females appear to have experienced worsening of general mental health, anxiety symptoms, and depression symptoms, which is consistent with evidence that women have experienced disproportionately greater burden from the pandemic.

## FIGURE LEGENDS

**Box 1 – Figure 1**. Illustration of change of 0.25 standardized mean difference effect size from hypothetical pre-COVID-19 (blue) to COVID-19 (black) symptom distributions. With a threshold located at one standard deviation above the pre-COVID-19 mean, the proportion of participants above the threshold would change from 16% to 23%. With a threshold two standard deviations above the pre-COVID-19 mean, the proportion would change from 2% to 4%.

**Figures 2a-2j.** Forest plots of standardized mean difference changes in general mental health for studies of the general population (2a), women or females (2b), men or males (2c), older adults (2d), young adults (2e) university students (2f), children and adolescents (2g), parents (2h), people with pre-existing medical conditions (2i), and people with pre-existing mental health conditions (2j).

**Figures 3a-3j.** Forest plots of standardized mean difference changes in anxiety symptoms for studies of the general population (3a), women or females (3b), men or males (3c), older adults (3d), young adults (3e), university students (3f), children and adolescents (3g), people with pre-existing medical conditions (3h), people with pre-existing mental health conditions (3i), and people who identify as sexual or gender minorities (3j).

**Figures 4a-4k.** Forest plots of standardized mean difference changes in depression symptoms for studies of the general population (4a), women or females (4b), men or males (4c), older adults (4d), young adults (4e), university students (4f), children and adolescents (4g), parents (4h), people with pre-existing medical conditions (4i), people with pre-existing mental health conditions (4j), and people who identify as sexual or gender minorities (4k).

