## Supplementary Material for "Comparison of Mental Health Symptoms Prior to and During COVID-19: Evidence from a Systematic Review and Meta-analysis of 134 Cohorts"

**Supplementary Material: Table of Contents**

The living systematic review was registered in the PROSPERO prospective register of systematic reviews (CRD 42020179703) and a protocol was uploaded to the Open Science Framework prior to initiation (https://osf.io/96csg/).

**Supplementary Material 1.** Search Strategies

**Supplementary Material 2.** Inclusion and Exclusion Coding Guides

**Supplementary Material 3.** Risk of Bias and Adequacy of Study Methods and Reporting

**Supplementary Table 1.** Characteristics of Included Studies with References

**Supplementary Table 2.** Risk of Bias and Adequacy of Methods and Reporting

**Supplementary Table 3.** Individual Study Results for General Mental Health

**Supplementary Table 4.** Individual Study Results for Anxiety Symptoms

**Supplementary Table 5.** Individual Study Results for Depression Symptoms

**Supplementary Figure 1.** Sensitivity Analysis of General Mental Health conducted with results from Savage et al. from October 2020 instead of April 2020

**Supplementary Figure 2.** Sensitivity Analysis of Anxiety Symptoms conducted with results from Henry et al. from September to October 2020 instead of April 2020

**Supplementary Figure 3.** Sensitivity Analysis of Anxiety Symptoms conducted with results from Henry et al. from March 2021 instead of April 2020

**Supplementary Figure 4.** Sensitivity Analysis of Depression Symptoms conducted with results from Henry et al. from September to October 2020 instead of April 2020

**Supplementary Figure 5.** Sensitivity Analysis of Depression Symptoms conducted with results from Henry et al. from March 2021 instead of April 2020

**References of Included Studies**

**Supplementary Material 1: Search Strategies**

**Ovid MEDLINE All**

†New subject heading added to original search on January 27, 2021

1. Quarantine/

2. social isolation/ or loneliness/ or physical distancing/†

3. psychology.fs. or psychology/

4. Mental Health/

5. mental disorders/

6. social stigma/

7. Fear/

8. Anxiety/

9. Depression/

10. Stress, Physiological/ or Stress, Psychological/

11. Anger/

12. Irritable Mood/

13. Grief/

14. burnout, psychological/ or burnout, professional/

15. or/1-15

16. (Quarantine* or Self-isolation or isolation or social distanc* or shelter*-in-place or psych* or mental health or mental illness* or mental disorder* or stigma or fear* or anxiety or anxious or depression or depressive or loneliness or stress* or trauma* or post-traumatic or posttraumatic or anger or mood* or irritability or irritable or emotional disturbance* or grief or burned out or burnout).tw,kf.

17. ((exp coronavirus/ or exp coronavirus infections/ or (betacoronavirus* or beta coronavirus* or coronavirus* or corona virus*).mp.) and (exp china/ or (china or chinese or hubei or wuhan).af.)) or (coronavirus* or corona virus* or betacoronavirus* or beta coronavirus*).mp.

18. (severe acute respiratory syndrome coronavirus 2 or "SARS CoV-2" or "SARSCoV 2" or SARSCoV2 or cov2 or "sars 2" or COVID or "coronavirus 2" or covid19 or nCov or ((new or Novel) adj3 coronavirus*) or ncp).mp. or ((exp pneumonia/ or pneumonia.mp.) and wuhan.af.)

19. 17 or 18

20. 15 or 16

21. 19 and 20

22. ("20191231" or 2020* or 2021* or 2022*).dt,ez,da.

23. 21 and 22

**Embase (Ovid)**

1. exp coronavirinae/

2. exp Coronavirus infection/

3. (betacoronavirus* or beta coronavirus* or coronavirus* or corona virus*).mp.

4. 1 or 2 or 3

5. exp China/

6. (china or chinese or hubei or wuhan).af.

7. 5 or 6

8. 4 and 7

9. (betacoronavirus* or beta coronavirus* or coronavirus* or corona virus*).mp.

10. (severe acute respiratory syndrome coronavirus 2 or "SARS CoV-2" or "SARSCoV 2" or SARSCoV2 or cov2 or "sars 2" or COVID or "coronavirus 2" or covid19 or nCov or ((new or Novel) adj3 coronavirus*) or ncp).mp.

11. (exp pneumonia/ or pneumonia.mp.) and wuhan.af.

12. 8 or 9 or 10 or 11

13. quarantine/

14. social isolation/ or isolation/ or patient isolation/

15. loneliness/

16. psychology/

17. mental health/

18. mental disease/

19. social stigma/

20. fear/

21. anxiety/

22. depression/

23. physiological stress/ or mental stress/

24. anger/

25. irritability/

26. exp grief/

27. exp burnout/

28. (mental disorder* or Quarantine* or Self-isolation or isolation or social distanc* or shelter*-in-place or psych* or mental health or mental illness* or stigma or fear* or anxiety or anxious or depression or depressive or loneliness or stress* or trauma* or post-traumatic or posttraumatic or anger or mood* or irritability or irritable or emotional disturbance* or grief or burned out or burnout).tw,kw.

29. or/13-27

30. 12 and 29

31. ("20191231" or 2020* or 2021* or 2022*).dc.

32. 30 and 31

**PsycINFO (Ovid)**

1. (coronavirus* or corona virus* or betacoronavirus* or beta coronavirus*).mp.

2. (severe acute respiratory syndrome coronavirus 2 or "SARS CoV-2" or "SARSCoV 2" or SARSCoV2 or cov2 or "sars 2" or COVID or "coronavirus 2" or covid19 or nCov or ((new or Novel) adj3 coronavirus*) or ncp).mp. or ((exp pneumonia/ or pneumonia.mp.) and wuhan.af.)

3. 1 or 2

4. ("20191231" or 2020* or 2021* or 2022*).up.

5. 3 and 4

**CINAHL**

| **Search ID#** | **Search Terms** |
| --- | --- |
| S26 | S11 AND S25 |
| S25 | S12 OR S13 OR S14 OR S15 OR S16 OR S17 OR S18 OR S19 OR S20 OR S21 OR S22 OR S23 OR S24 |
| S24 | TI ( (mental disorder* or Quarantine* or Self-isolation or isolation or social distanc* or shelter*-in-place or psych* or mental health or mental illness* or stigma or fear* or anxiety or anxious or depression or depressive or loneliness or stress* or trauma* or post-traumatic or posttraumatic or anger or mood* or irritability or irritable or emotional disturbance* or grief or burned out or burnout) ) OR AB ( (mental disorder* or Quarantine* or Self-isolation or isolation or social distanc* or shelter*-in-place or psych* or mental health or mental illness* or stigma or fear* or anxiety or anxious or depression or depressive or loneliness or stress* or trauma* or post-traumatic or posttraumatic or anger or mood* or irritability or irritable or emotional disturbance* or grief or burned out or burnout) ) |
| S23 | (MH "Burnout, Professional") |
| S22 | (MH "Grief+") |
| S21 | (MH "Anger") |
| S20 | (MH "Stress, Physiological") OR (MH "Stress, Psychological") |
| S19 | (MH "Depression") |
| S18 | (MH "Anxiety") |
| S17 | (MH "Fear") |
| S16 | (MH "Stigma") |
| S15 | (MH "Mental Health") or (MH "Mental Disorders") |
| S14 | (MH "Psychology") |
| S13 | (MH "Social Isolation") OR (MH "Loneliness") or (MH “Social Distancing”) or (MH “Stay at Home Orders”) † |
| S12 | (MH "Quarantine") |
| S11 | S7 OR S8 OR S9 OR S10 |
| S10 | ( (MH "Pneumonia+") or TI (pneumonia) OR AB (pneumonia) ) AND ( TI (wuhan) OR AB (wuhan) OR AF (wuhan) ) |
| S9 | TI ( (severe acute respiratory syndrome coronavirus 2 or "SARS CoV-2" or "SARSCoV 2" or SARSCoV2 or cov2 or "sars 2" or COVID or "coronavirus 2" or covid19 or nCov or ((new or Novel) N3 coronavirus*) ) OR AB ( (severe acute respiratory syndrome coronavirus 2 or "SARS CoV-2" or "SARSCoV 2" or SARSCoV2 or cov2 or "sars 2" or COVID or "coronavirus 2" or covid19 or nCov or ((new or Novel) N3 coronavirus*) ) or (MH “Covid 19”) † |
| S8 | TI ( (betacoronavirus* or beta coronavirus* or coronavirus* or corona virus*) ) OR AB ( (betacoronavirus* or beta coronavirus* or coronavirus* or corona virus*)) |
| S7 | S5 AND S6 |
| S6 | S1 OR S2 |
| S5 | S3 OR S4 |
| S4 | TI ( (china or chinese or hubei or wuhan) ) OR AB ( (china or chinese or hubei or wuhan) ) OR AF ( (china or chinese or hubei or wuhan) ) OR SO ( (china or chinese or hubei or wuhan) ) |
| S3 | (MH "China+") |
| S2 | TI ( (betacoronavirus* or beta coronavirus* or coronavirus* or corona virus*) ) OR AB ( (betacoronavirus* or beta coronavirus* or coronavirus* or corona virus*) ) |
| S1 | (MH "Coronavirus+") OR (MH "Coronavirus Infections+") |

**Web of Science**

TOPIC: (Quarantine* or "Self-isolation" or isolation or "social distanc*" or "shelter*-in-place" or psych* or "mental health" or "mental illness*" or "mental disorder*" or stigma or fear* or anxiety or anxious or depression or depressive or loneliness or stress* or trauma* or "post-traumatic" or posttraumatic or anger or mood* or irritability or irritable or "emotional disturbance*" or grief or "burned out" or burnout) AND TOPIC: ((coronavirus* or "corona virus*" or betacoronavirus* or "beta coronavirus*" or "severe acute respiratory syndrome coronavirus 2" or "SARS CoV-2" or "SARSCoV 2" or SARSCoV2 or cov2 or "sars 2" or COVID or "coronavirus 2" or covid19 or nCov or "Novel coronavirus*" or "new coronavirus*"))

Indexes=SCI-EXPANDED, SSCI, A&HCI, CPCI-S, CPCI-SSH, BKCI-S, BKCI-SSH, ESCI, CCR-EXPANDED, IC Timespan=Year to date

**China National Knowledge Infrastructure**

Restricted to disciplines: Medical and Public Health & Social science

TI=(隔离+封城+社交距离+方舱+心理+心理健康+精神卫生+精神疾病+心理疾病+污名+耻辱+羞辱+恐惧+焦虑+抑郁+孤独+压力+应激+创伤+创伤后+愤怒+情绪+心情+易怒+情绪障碍+心理障碍+哀伤+悲伤+悲痛+悲哀+忧郁+倦怠)*(新冠+新型冠状) OR AB=(隔离+封城+社交距离+方舱+心理+心理健康+精神卫生+精神疾病+心理疾病+污名+耻辱+羞辱+恐惧+焦虑+抑郁+孤独+压力+应激+创伤+创伤后+愤怒+情绪+心情+易怒+情绪障碍+心理障碍+哀伤+悲伤+悲痛+悲哀+忧郁+倦怠)*(新冠+新型冠状)

**Wanfang**

题名:("隔离"+封城+"社交距离"+方舱+心理+"心理健康"+"精神卫生"+"精神疾病"+"心理疾病"+污名+耻辱+羞辱+恐惧+焦虑+抑郁+孤独+压力+应激+创伤+"创伤后"+愤怒+情绪+心情+易怒+”情绪障碍"+"心理障碍”+哀伤+悲伤+悲痛+悲哀+忧郁+倦怠)*("新冠"+"新型冠状")+摘要:("隔离"+封城+"社交距离"+方舱+心理+"心理健康"+"精神卫生"+"精神疾病"+"心理疾病"+污名+耻辱+羞辱+恐惧+焦虑+抑郁+孤独+压力+应激+创伤+"创伤后"+愤怒+情绪+心情+易怒+"情绪障碍"+"心理障碍"+哀伤+悲伤+悲痛+悲哀+忧郁+倦怠)*("新冠"+"新型冠状")

We made several amendments to the original search strategies. Since the Wanfang database cannot export more than 5000 references at once, we broke the search strategies into two or more smaller search strings to get all the references. The four changes on September 1, 2020, September 28, 2020, October 15, 2020 and October 18, 2020 are all for this purpose.

To make this process more efficient, the disciplines of the China National Knowledge Infrastructure database were restricted to Medical and Public Health AND Social science subgroup 2 and those of Wanfang database were restricted to Medicine and Health AND Culture, Science, Education and PE disciplines on October 23, 2020.

September 1, 2020

**Wanfang**

题名:("隔离"+封城+"社交距离"+方舱+心理+"心理健康"+"精神卫生"+"精神疾病"+"心理疾病"+污名+耻辱+羞辱+恐惧+焦虑)*("新冠"+"新型冠状")+摘要:("隔离"+封城+"社交距离"+方舱+心理+"心理健康"+"精神卫生"+"精神疾病"+"心理疾病"+污名+耻辱+羞辱+恐惧+焦虑)*("新冠"+"新型冠状")

题名:("隔离"+封城+"社交距离"+方舱+抑郁+孤独+压力+应激+创伤+"创伤后"+愤怒+情绪+心情+易怒+"情绪障碍"+"心理障碍"+哀伤+悲伤+悲痛+悲哀+忧郁+倦怠)*("新冠"+"新型冠状")+摘要:("隔离"+封城+"社交距离"+方舱+抑郁+孤独+压力+应激+创伤+"创伤后"+愤怒+情绪+心情+易怒+"情绪障碍"+"心理障碍"+哀伤+悲伤+悲痛+悲哀+忧郁+倦怠)*("新冠"+"新型冠状")

September 28, 2020

**Wanfang**

题名:("隔离"+封城+"社交距离"+方舱+心理+"心理健康"+"精神卫生"+"精神疾病"+"心理疾病"+污名+耻辱+羞辱+恐惧+焦虑)*("新冠"+"新型冠状")+摘要:("隔离"+封城+"社交距离"+方舱+心理+"心理健康"+"精神卫生"+"精神疾病"+"心理疾病"+污名+耻辱+羞辱+恐惧+焦虑)*("新冠"+"新型冠状")

题名:("隔离"+封城+"社交距离"+方舱+抑郁+孤独+压力+应激+创伤+"创伤后")*("新冠"+"新型冠状")+摘要:("隔离"+封城+"社交距离"+方舱+抑郁+孤独+压力+应激+创伤+"创伤后")*("新冠"+"新型冠状")

题名:("隔离"+封城+"社交距离"+方舱+愤怒+情绪+心情+易怒+"情绪障碍"+"心理障碍"+哀伤+悲伤+悲痛+悲哀+忧郁+倦怠)*("新冠"+"新型冠状")+摘要:("隔离"+封城+"社交距离"+方舱+愤怒+情绪+心情+易怒+"情绪障碍"+"心理障碍"+哀伤+悲伤+悲痛+悲哀+忧郁+倦怠)*("新冠"+"新型冠状")

October 15, 2020

**Wanfang**

题名:("隔离"+封城+"社交距离"+方舱+心理+"心理健康"+"精神卫生"+"精神疾病"+"心理疾病")*("新冠"+"新型冠状")+摘要:("隔离"+封城+"社交距离"+方舱+心理+"心理健康"+"精神卫生"+"精神疾病"+"心理疾病")*("新冠"+"新型冠状")

题名:("隔离"+封城+"社交距离"+方舱+污名+耻辱+羞辱+恐惧+焦虑+抑郁+孤独+压力)*("新冠"+"新型冠状")+摘要:("隔离"+封城+"社交距离"+方舱+污名+耻辱+羞辱+恐惧+焦虑+抑郁+孤独+压力)*("新冠"+"新型冠状")

题名:("隔离"+封城+"社交距离"+方舱+应激+创伤+"创伤后"+愤怒+情绪+心情+易怒+"情绪障碍"+"心理障碍"+哀伤+悲伤+悲痛+悲哀+忧郁+倦怠)*("新冠"+"新型冠状")+摘要:("隔离"+封城+"社交距离"+方舱+应激+创伤+"创伤后"+愤怒+情绪+心情+易怒+"情绪障碍"+"心理障碍"+哀伤+悲伤+悲痛+悲哀+忧郁+倦怠)*("新冠"+"新型冠状")

October 18, 2020

**Wanfang**

题名:("隔离"+封城+"社交距离"+方舱+心理+"心理健康"+"精神卫生"+"精神疾病"+"心理疾病")*("新冠"+"新型冠状")+摘要:("隔离"+封城+"社交距离"+方舱+心理+"心理健康"+"精神卫生"+"精神疾病"+"心理疾病")*("新冠"+"新型冠状")

题名:("隔离"+封城+"社交距离"+方舱+污名+耻辱+羞辱+恐惧+焦虑+抑郁)*("新冠"+"新型冠状")+摘要:("隔离"+封城+"社交距离"+方舱+污名+耻辱+羞辱+恐惧+焦虑+抑郁)*("新冠"+"新型冠状")

题名:("隔离"+封城+"社交距离"+方舱+孤独+压力)*("新冠"+"新型冠状")+摘要:("隔离"+封城+"社交距离"+方舱+孤独+压力)*("新冠"+"新型冠状")

题名:("隔离"+封城+"社交距离"+方舱+应激+创伤+"创伤后"+愤怒+情绪+心情+易怒+"情绪障碍"+"心理障碍"+哀伤+悲伤+悲痛+悲哀+忧郁+倦怠)*("新冠"+"新型冠状")+摘要:("隔离"+封城+"社交距离"+方舱+应激+创伤+"创伤后"+愤怒+情绪+心情+易怒+"情绪障碍"+"心理障碍"+哀伤+悲伤+悲痛+悲哀+忧郁+倦怠)*("新冠"+"新型冠状")

October 23, 2020

**China National Knowledge Infrastructure**

Restricted to disciplines: Medical and Public Health & Social science subgroup 2

TI=(隔离+封城+社交距离+方舱+心理+心理健康+精神卫生+精神疾病+心理疾病+污名+耻辱+羞辱+恐惧+焦虑+抑郁+孤独+压力+应激+创伤+创伤后+愤怒+情绪+心情+易怒+情绪障碍+心理障碍+哀伤+悲伤+悲痛+悲哀+忧郁+倦怠)*(新冠+新型冠状) OR AB=(隔离+封城+社交距离+方舱+心理+心理健康+精神卫生+精神疾病+心理疾病+污名+耻辱+羞辱+恐惧+焦虑+抑郁+孤独+压力+应激+创伤+创伤后+愤怒+情绪+心情+易怒+情绪障碍+心理障碍+哀伤+悲伤+悲痛+悲哀+忧郁+倦怠)*(新冠+新型冠状)

**Wanfang**

Restricted to disciplines: Medicine and Health & Culture, Science, Education and PE

题名:("隔离" or 封城 or "社交距离" or 方舱 or 心理 or "心理健康" or "精神卫生" or "精神疾病" or "心 理疾病" or 污名 or 耻辱 or 羞辱 or 恐惧 or 焦虑 or 抑郁 or 孤独 or 压力 or 应激 or 创伤 or "创伤后" or 愤怒 or 情绪 or 心情 or 易怒 or "情绪障碍" or "心理障碍" or 哀伤 or 悲伤 or 悲痛 or 悲哀 or 忧郁 or 倦怠) and ("新冠" or "新型冠状") or 摘要:("隔离" or 封城 or "社交距离" or 方舱 or 心理 or "心理健康" or "精神卫生" or "精神疾病" or "心理疾病" or 污名 or 耻辱 or 羞辱 or 恐惧 or 焦虑 or 抑郁 or 孤独 or 压力 or 应激 or 创伤 or "创伤后" or 愤怒 or 情绪 or 心情 or 易怒 or "情绪障碍" or "心理障碍" or 哀伤 or 悲伤 or 悲痛 or 悲哀 or 忧郁 or 倦怠) and ("新冠" or "新型冠状")

**MedRxiv (pre-prints)**

Search 1: (isolation OR “mental health” OR “mental illness” OR “mental disorder”) AND (COVID OR covid19)

Search 2: (psychology OR psychological OR psychosocial OR anxiety OR depression OR stress or trauma) AND (COVID OR covid19)

**Open Science Framework (pre-prints)**

(isolation OR psychology OR psychological OR psychosocial OR “mental health” OR “mental illness” OR “mental disorder” OR anxiety OR depression OR stress or trauma) AND (coronavirus OR COVID OR covid19)

**Supplementary Material 2: Inclusion and Exclusion Coding Guides for Main Changes Review Plus Additional Criteria for Present Report**

Title and Abstract Review:

**Exclude: not original human data or a case study or case series. If it is clear from the title and abstract that the article is not an original report of primary data, but, for example, a letter, editorial, systematic review or meta-analysis, or it is a single case study or case series, then it is excluded. Studies reporting only on animal, cellular, or genetic data are also excluded. Conference abstracts are included.**

**Exclude: not a study of any population affected by the COVID-19 outbreak. If it is clear from the title or abstract that the study is not about any population affected by the COVID-19 outbreak, it is excluded. Studies that include fewer than 100 participants, are excluded. If a longitudinal study has baseline sample size with at least 100 participants, but no follow-up with at least 100 participants, then we exclude the study (and document); if its baseline and at least one follow-up have more than 100 participants, we include the study.**

**Exclude: not a study which reports mental health symptom changes longitudinally pre-COVID-19 to COVID-19 or during COVID-19. If it is clear from the title or abstract that the study does not report proportions of participants meeting diagnostic criteria using a validated diagnostic interview or validated mental health scale, or proportions of symptoms (based on a threshold or measured continuously) prior to and after the start of COVID-19, or longitudinally during COVID-19, then it will be excluded.**

**For pre-COVID versus during-COVID studies, pre- and during- samples must include the same cohort, not different representative samples. Pre- and during-samples should have less than 10% difference in the participants in the sample* or should statistically account for missing data, i.e., if N between the samples differs by more than 10%, modelling or imputation is needed to evaluate results for all participants. Pre-COVID data needs to be collected prior to 2020 (or at least 80% of the participants’ data need be collected prior to 2020 if collection spans from 2019 to 2020) and after 2018(or at least 80% of the participants’ data need to be collected after 2018 if collection spans from pre-2018 to 2018).**

For studies with multiple waves across COVID, if there are pre-pandemic time points, the most recent pre-pandemic wave needs to be in 2018 or later; if the most recent pre-pandemic wave spans from pre-2018 to 2018, at least 80% of the data need to be collected in 2018. Studies with multiple waves across COVID-19 must have at least two time points that have less than 10% difference in the participants in the sample*, or should statistically account for missing data, regardless of whether or not the study has pre-COVID assessments. If outcomes from the study are only shown graphically without eligible numerical values, exclude the study. At least 90% of participants in assessments from two time points need to be the same participants. In a three-wave survey, if N-T1 = 1000, N – T2 = 500, and N – T3 = 500, T2 and T3 would only be eligible if at least 90% of the participants at each time point were the same. It is not enough to just have a total N within 10%.

**Include: study eligible to be included in full-text review.**

Full-text Review:

**Exclude: not original human data or a case study or case series. If the article is not an original report of primary data, but, for example, a letter, editorial, systematic review or meta-analysis, or it is a single case study or case series, then it is excluded. Studies reporting only on animal, cellular, or genetic data are also excluded. Conference abstracts are included.**

**Exclude: not a study of any population affected by the COVID-19 outbreak. If it is clear from the full text that the study is not about any population affected by the COVID-19 outbreak, it is excluded. Studies that include fewer than 100 participants, are excluded. If a longitudinal study has baseline sample size with at least 100 participants, but no follow-up with at least 100 participants, then we exclude the study (and document); if its baseline and at least one follow-up have more than 100 participants, we include the study.**

**Exclude: not a study which reports mental health symptom changes longitudinally pre-COVID-19 to COVID-19 or during COVID-19. If it is clear from the title or abstract that the study does not report continuous scores of symptom levels or proportions of participants meeting the threshold on a validated scale, or diagnostic criteria using a validated diagnostic interview prior to and after the start of COVID-19, or longitudinally during COVID-19, then it will be excluded.**

For pre-COVID versus during-COVID studies, pre- and during- samples must include the same cohort, not different representative samples. Pre- and during-samples should have less than 10% difference in the participants in the sample* or should statistically account for missing data, i.e., if N between the samples differs by more than 10%, modelling or imputation is needed to evaluate results for all participants. Pre-COVID data needs to be collected prior to 2020 (or at least 80% of the participants’ data need be collected prior to 2020 if collection spans from 2019 to 2020) and after 2018 (or at least 80% of the participants’ data need to be collected after 2018 if collection spans from pre-2018 to 2018).

For studies with multiple waves across COVID, if there are pre-pandemic time points, the most recent pre-pandemic wave needs to be in 2018 or later; if the most recent pre-pandemic wave spans from pre-2018 to 2018, at least 80% of the data need to be collected in 2018. Studies with multiple waves across COVID-19 must have at least two time points that have less than 10% difference in the participants in the sample, or should statistically account for missing data, regardless of whether or not the study has pre-COVID assessments. If outcomes from the study are only shown graphically without eligible numerical values, exclude the study

**Include: study eligible for inclusion in systematic review.**

**Additional Criterion for Present Report: (1) Eligible pre-COVID-19 assessments had to be done** between January 1, 2018 and December 31, 2019; (2) Only studies that compared pre-COVID-19 and COVID-19 assessments were included but not studies with longitudinal data only during COVID-19.

**Supplementary Material 3: Risk of Bias and Adequacy of Study Methods and Reporting**

**Q1. Was the sample frame appropriate to address the target population?**

**Yes: The sampling frame was a true or close representation of the target population.**

**No: The sampling frame was NOT a true or close representation of the target population.**

**Unclear: Not enough information provided to determine.**

**Q2. Were study participants recruited in an appropriate way?**

**Yes:** A census was undertaken, OR, some form of random selection was used to select the sample (e.g. simple random sampling, stratified random sampling, cluster sampling, systematic sampling).

**No:** A census was NOT undertaken, AND some form of random selection was NOT used to select the sample.

**Unclear:** Not enough information provided to determine.

**Q3. Was the sample size adequate?**

**Yes:** There is evidence that the authors conducted a sample size calculation to determine an adequate sample size OR the study was large enough (e.g., a large national survey) whereby a sample size calculation is not required. In these cases, sample size can be considered adequate. If at least 200 participants are included for continuous outcomes and 250 for proportions, this is considered low risk.

**No:** The authors did not reach their intended sample size, or no sample size calculation is provided and there are < 100 participants for continuous outcomes, or < 125 for proportions.

**Unclear:** No sample size calculation is provided, and between 100-199 participants are included for continuous outcomes or between 125-249 for proportions.

**Q4. Were the study participants and setting described in detail?**

**Yes:** Data included age, sex, and at least 1 socioeconomic indicator (e.g., income, education, work status).

**No:** The minimum sociodemographic variables have not been reported.

**Unclear:** Not stated

**Q5. Was the response rate adequate and was the data analysis conducted with sufficient coverage?**

**Yes:** The overall response rate or response rate for intended subgroups was >/=75%, OR, an analysis was performed that established that there was not a substantive difference in relevant demographic characteristics between responders and non-responders within a subgroup (if non-response too high (e.g., > 50%), code “No”)

**No:** The overall response rate or response rate for subgroups was <75%, and if any analysis comparing responders and non-responders was done, it showed a meaningful difference in relevant demographic characteristics between responders and non-responders.

**Unclear:** Not enough information provided to determine.

**Q6. Were valid methods used for the identification of the outcome variable?**

**Yes:** The study instrument had been shown to have reliability and validity, e.g., test-retest, piloting, validation in a previous study, etc.

**No:** The study instrument had NOT been shown to have reliability or validity.

**Unclear:** Not stated.

**Q7. Was the mental health outcome measured in a standard, reliable way for all participants?**

**Yes:** All self-report data were collected directly from the participants. Any clinical interview data includes at least information about the interviewers’ level of education or training received. The same mode of data collection was used for all participants. All aspects of this question must be present (where relevant).

**No:** In some instances, data were collected from a proxy (e.g., a spouse). The qualifications of clinical interviewers are not reported or not appropriate. The same mode of data collection was NOT used for all participants. If any aspects of this item are absent, it is high risk.

**Unclear**: Not stated.

**Q8. Was there appropriate statistical analysis?**

**Yes:** Continuous variables report (1) mean (SD) of change or (2) pre mean (SD) and post mean (SD) with/out correlation between pre and post scores. For dichotomous variables, numerator, denominator, and percentages are clearly reported. Continuous variables are not artificially dichotomized. The statistical analyses section is detailed enough for readers to understand change scores (see STROBE reporting guidelines, if necessary).

**No:** Continuous variables do not include a report of the (1) mean (SD) of change or (2) pre mean (SD) and post mean (SD) with/out correlation between pre and post scores. For dichotomous variables, the numerator, denominator, or percentages are not clearly reported. The statistical analyses section does not clearly describe the methods used to assess change scores.

**Q9. Was the follow-up rate adequate, and if not, was the low follow-up rate managed appropriately?**

**Yes:** At least 75% of those who participated in the pre-COVID-19 assessment(s) provided follow-up responses and had their responses included in the follow-up, OR, an analysis was performed that showed no substantive difference in relevant demographic characteristics between participants who stayed in the study and drop-outs (if dropout too high (e.g. > 50%), code “No”).

**No:** Less than 75% of those participated in the pre-COVID-19 assessment(s) provided responses and had their responses included in the follow-up, and if any analysis comparing participants who stayed in the study and drop-outs was done, it showed a substantive difference in relevant demographic characteristics between the two groups.

**Unclear:** Not stated.

**Supplementary Table 1. Characteristics of unique studies (N=137) from included cohorts (N=134)^a^**

| **First Author** | **Outcome Domains** | | | **Description of Participants** | **Country(ies) of Participants** | **Pre- and Post-COVID-19 Data Collection** | **N Participants** | **Participant Age** | **% Female or Women** |
| --- | --- | --- | --- | --- | --- | --- | --- | --- | --- |
|  | **General Mental Health** | **Anxiety Symptoms** | **Depression Symptoms** |  |  |  |  | **Mean (SD) or % in Range of Years** |  |
| **General Population** | | | | | | | | | |
| Bulbulia^S1^ | K6 |  |  | Convenience sample of adults aged 18 to 65 from the New Zealand Attitudes and Values Study (NZAVS) | New Zealand | NR/2018 | 940 | 52 (13) | 65% |
|  |  |  |  |  |  | 03-04/2020 |  |  |  |
| Castellini^S2^ | BSI-GSI |  |  | Convenience sample of adults aged 18 to 60 years recruited via “convenience and snowballing” methods | Italy | 12/2019 | 130 | 34 (14)^b^ | 75% |
|  |  |  |  |  |  | 04-05/2020 |  |  |  |
| Chan^S3^ |  | HAI |  | Convenience sample of adults based in Hong Kong and participated in a previous study prior to the pandemic | Hong Kong, China | 07/2019 | 279 | 27 (9) | 74% |
|  |  |  |  |  |  | 07/2020 |  |  |  |
| Finucane^S4^ | K6 |  |  | Participants from the Pittsburgh Hill/Homewood Research on Neighborhood Changes and Health study | USA | 05-09/2018 | 419 | 62 (14) | 82% |
|  |  |  |  |  |  | 06-09/2020 |  |  |  |
| Ge^S5^ |  | GAD-7 | PHQ-9 | Convenience sample of adults recruited from the WeChat of China online social media platform | China | 01-12/2019 | 1,547-1,978 | Anxiety sample = 30 (10); Depression sample = 33 (11) | Anxiety sample: 29; Depression sample: 26 |
|  |  |  |  |  |  | 02-03/2020 |  |  |  |
| Haliwa^S6^ |  | Sample 1: GAD-7 Sample 2: DASS-21-Anxiety Sample 3: GAD-7 | Sample 1:PHQ-8 Sample 2: DASS-21-Depression Sample 3: PHQ-8 | U.S. residents recruited through Amazon’s Mechanical Turk | USA | 09-12/2019 | Sample 1: 300; Sample 2: 146; Sample 3: 142 | Sample 1: 41 (12); Sample 2: 44 (13); Sample 3: 41 (13) | Sample 1: 59; Sample 2: 53; Sample 3: 50 |
|  |  |  |  |  |  | 04-06/2020 |  |  |  |
| Kanbur^S7^ | SCL-90-R | SCL-90-R Anxiety | SCL-90-R Depression | Turkish office workers who were enrolled in another study before the pandemic | Turkey | NR/2019 | 400 | NR | NR |
|  |  |  |  |  |  | NR/2020 |  |  |  |
| Katz, B^S8^ | RRQ | DASS-21 Anxiety | DASS-21 Depression | Convenience sample of adults recruited via an online crowdsourcing research platform | Canada, Ireland, UK, USA | 04/2019 | 218 | 43 (13) | 54% |
|  | DToS |  |  |  |  |  |  |  |  |
|  |  |  |  |  |  | 04/2020 |  |  |  |
| Latikka^S9^ | GHQ-12 |  |  | Participants from the Social Media at Work in Finland Survey | Finland | 09-10/2019 | 840 | 44 (11) | 44% |
|  |  |  |  |  |  | 03-04/2020 |  |  |  |
| Megias-Robles^S10^ | PANAS-NA |  |  | Convenience sample of participants recruited from an adult community sample | Spain | 11/2019 | 102 | 30 (13) | 66% |
|  |  |  |  |  |  | 04/2020 |  |  |  |
| Pierce^S11^ | GHQ-12 |  |  | National probability-based sample of adults aged ≥ 18 years (United Kingdom Household Longitudinal Study) | UK | Pre-COVID-19 waves^c^ | 15,376^S11,d^  10,918^S12^ | 18-34 (12)^e^ | 47%^S11^ 58%^S12^ |
| Daly^S12^ |  |  |  |  |  |  |  | 35-49 (22)^e^ |  |
|  |  |  |  |  |  | 04-09/2020 |  | 50-64 (34)^e^ |  |
|  |  |  |  |  |  |  |  | 65+ (32)^e^ |  |
| Shimura^S13^ | BJSQ (Psychological and Physical) |  |  | Convenience sample of office workers who started remote work in 2020 | Japan | NR/2019 | 3,123 | 37 (11) | 43% |
|  |  |  |  |  |  | NR/2020 |  |  |  |
| Soltanzadeh^S14^ | GHQ-28 |  |  | Employees of three oil refineries in southern Iran who had at least 1 year of work experience | Iran | 11/2019 | 823-850 | 35 (13) | 19% |
|  |  |  |  |  |  | 07/2020 |  |  |  |
| Thygesen^S15^ | SWEMWBS |  |  | Participants from Danish Health and Wellbeing Survey | Denmark | 09-12/2019 | 4,234 | Age range (%): 15-44 (27); 45-59 (30); 60-74 (33); 75+ (10) | 58% |
|  |  |  |  |  |  | 09-11/2020 |  |  |  |
| van der Velden^S16^ | MHI-5 |  |  | National probability-based sample of adults aged ≥ 18 years (Longitudinal Internet Studies for the Social Sciences) | The Netherlands | 03/2019 | 3,983 | 18-34 (25)^f^ | 51% |
| van der Velden^S17^ |  |  |  |  |  | 11-12/2019 | 4,064 | 35-49 (23)^f^ |  |
|  |  |  |  |  |  |  |  | 50-64 (26)^f^ |  |
|  |  |  |  |  |  | 03/2020 |  | 65+ (26)^f^ |  |
|  |  |  |  |  |  | 11-12/2020 |  |  |  |
| Wanberg^S18^ |  |  | PHQ-8 | Individuals aged 30 to 80 years from the RANT American Life Panel | USA | 04-06/2019 | 1,143 | 53 (14) | 56% |
|  |  |  |  |  |  | 04/2020 |  |  |  |
| **Older Adults** | | | | | | | | | |
| Bartlett^S19^ |  | HADS-A | HADS-D | Adults aged ≥ 50 years from the Island Study Linking Ageing and Neurodegenerative Disease | Australia | 10/2019 | 1,671 | 63 (7) | 73% |
|  |  |  |  |  |  | 04-06/2020 |  |  |  |
| Briggs^S20^ |  |  | CES-D-8 | Nationally representative sample of community-dwelling older adults aged ≥ 50 years who took part in the Irish Longitudinal Study on Ageing (TILDA) | Ireland | NR/2018 | 3,490 | 70 (14) | 56% |
|  |  |  |  |  |  | 07-11/2020 |  |  |  |
| Creese^S21^ |  | GAD-7 | PHQ-9 | National convenience sample of adults aged ≥ 50 years recruited via publicity | UK | 10/2019 | 3,281 | 67 (7) | 80% |
|  |  |  |  |  |  | 05-06/2020 |  |  |  |
| Eliasen^S22^ | WHOQOL-BREF |  |  | Participants from the Faroese Septuagenarians cohort | Denmark | 12/2017-01/2019^g^ | 227 | 84 (1) | 52% |
|  | WHOQOL-BREF (psychological health) |  |  |  |  |  |  |  |  |
|  |  |  |  |  |  | 06-07/2020 |  |  |  |
| Herrera^S23^ |  | GAI-SF | PHQ-9 | Participants from the V National Survey on Quality of Life in Older Adults | Chile | 11/2019 | 721 | 72 (NR) | 70% |
|  |  |  |  |  |  | 09/2020 |  |  |  |
| Kera^S24^ | WHO-5-J |  |  | Community-dwelling older adults living in Itabashi Ward, Tokyo, who had participated in the Otassha Study | Japan | 09-10/2019 | 533 | 73 (6) | 62% |
|  |  |  |  |  |  | 06-07/2020 |  |  |  |
| Kivi^S25^ | SWLS |  |  | “Nationally representative” sample of older adults born 1949 to 1955 | Sweden | NR/2019 | 1,071 | 68 (2) | 47% |
|  |  |  |  |  |  | 03-04/2020 |  |  |  |
| Lee^S26^ |  |  | PHQ-9 | Participants from the PopulatiON HEalth and Eye Disease PRofile in Elderly Singaporeans study | Singapore | 12/2017-11/2019^g^ | 496 | 74 (8) | 55% |
|  |  |  |  |  |  | 05-06/2020 |  |  |  |
| Martínez^S27^ | PWBS |  | CES-D | Community-dwelling older adults aged 65 to 87 years | Spain | 10/2019 | 141 | 73 (5) | 60% |
|  | PERMA - PA |  |  |  |  |  |  |  |  |
|  | PERMA - NA |  |  |  |  | 04/2020 |  |  |  |
| Okely^S28^ | WEMWBS |  |  | Surviving members of cohort of all children born in 1936 and attending school in Scotland in 1947 | Scotland (UK) | NR/2017-NR/2019^g^ | 137 | 84 (NR) | 48% |
|  |  |  |  |  |  | 05-06/2020 |  |  |  |
| Rentscher^S29^ |  | STAI-State | CES-D | Women aged ≥ 60 years who were nonmetastatic breast cancer survivors | USA | 02-06/2019 | 262 | 68 (5) | 100% |
|  |  |  |  |  |  | 05-09/2020 |  |  |  |
| Rentscher^S29^ |  | STAI-State | CES-D | Women aged ≥ 60 years who were matched controls | USA | 02-06/2019 | 165 | 68 (6) | 100% |
|  |  |  |  |  |  | 05-09/2020 |  |  |  |
| Sardella^S30^ | SF-12 Mental Component Summary |  |  | Participants aged ≥ 65 | Italy | 10/2018-10/2019 | 104 | 80 (7) | 70% |
|  |  |  |  |  |  | 04/2020 |  |  |  |
| Siew^S31^ | WHOQOL-AGE | GAI-SF |  | Participants from the Community Health and Intergenerational study | Singapore | 02/2018-01/2020^h^ | 411 | 69 (6) | 65% |
|  |  |  |  |  |  | 05-06/2020 |  |  |  |
| van den Besselaar^S32^ |  | HADS-A | CES-D-10 | Participants from the Longitudinal Aging Study Amsterdam | The Netherlands | NR/2018-2019 | 984-1,068 | 74 (8) | 53% |
|  |  |  |  |  |  | 06-10/2020 |  |  |  |
| van Tilburg^S33^ | MHI-5 |  |  | National probability-based sample of adults aged ≥ 65 years (Longitudinal Internet Studies for the Social Sciences) | The Netherlands | 10-11/2019 | 1,679 | 73 (NR) | 49% |
|  |  |  |  |  |  | 05/2020 |  |  |  |
| Wang, Yi^S34^ | K10 |  |  | Adults aged ≥ 60 years who were part of the Shandong Rural Elderly Health Cohort (SREHC) | China | 05-06/2019 | 2,745 | Median (Age range): 70 (60-100) | 64% |
|  |  |  |  |  |  | 08-09/2020 |  |  |  |
| Wong, S^S35^ |  | GAD-7 | PHQ-9 | Adults aged ≥ 60 with ≥ 2 chronic medical conditions from 4 primary care clinics | Hong Kong, China | 04/2018-03/2019 | 583 | 71 (6) | 73% |
|  |  |  |  |  |  | 03-04/2020 |  |  |  |
| Yu^S36^ |  | GAI | GDS | Individuals aged 60 to 99 years living in the western region of Singapore | Singapore | 02/2018-01/2020^h^ | 419 | 69 (6) | 66% |
|  |  |  |  |  |  | 05-06/2020 |  |  |  |
| **Young Adults** | | | | | | | | | |
| Islam^S37^ | K10 |  |  | Individuals aged 20 to 21 who took part in the Longitudinal Study of Australian Children (LSAC) survey | Australia | NR/2018 | 1,110 | 21 (0) | 59% |
|  |  |  |  |  |  | 10-12/2020 |  |  |  |
| Marmet^S38^ |  |  | MDI | Swiss adult men who enrolled in a longitudinal cohort in 2010-2011 during medical evaluation for mandatory military service | Switzerland | 04/2019-02/2020^h^ | 2,345 | 29 (13) | 0% |
|  |  |  |  |  |  | 05-06/2020 |  |  |  |
| Rimfeld^S39^ |  | SMGAD-A | SMFQ | Adult twins born 1994-1996 who were enrolled in a longitudinal cohort at age 18 months | UK | NR/2018 | 3,563-3,694 | 24-26 (100%) | 63% |
|  |  |  |  |  |  | 04-05/2020 |  |  |  |
| Romm^S40^ |  |  | PHQ-2 | Adults aged 18 to 34 years in one of the six metropolitan statistical areas who participated in the Vape Shop Advertising, Place characteristics and Effects Surveillance study | USA | 09/2019 | 1,082 | 25 (5) | 51% |
|  |  |  |  |  |  | 03/2020 |  |  |  |
| Tanioka^S41^ | K6-J |  |  | Participants aged 15 to 30 years who took part in a comprehensive prospective research project on sleep behavior, sleep problems, psychological distress, and quality of life in young adults | Japan | 10/2019 | 2,222 | 21 (4) | 76% |
|  | SF-8 - MCS |  |  |  |  |  |  |  |  |
|  |  |  |  |  |  | 05/2020 |  |  |  |
| Villadsen^S42^ | K6 |  |  | Participants from the Millennium Cohort Study who were born in 2001 | UK | NR/2018 | 1,615 | Range: 19-20 | NR |
|  |  |  |  |  |  | 05/2020 |  |  |  |
| Watkins-Martin^S43^ |  | GAD-7 | CES-D-12 | Participants born in 1997-98 in Quebec, Canada who participated in the Québec Longitudinal Study of Child Development | Canada | NR/2018 | 1,039 | 22 (NR) | 60% |
|  |  |  |  |  |  | 08/2020 |  |  |  |
| **University Students** | | | | | | | | | |
| Conceição^S44^ |  | GAD-7 | PHQ-9 | First-year students at the University of Porto | Portugal | 10/2019 | 341 | 20 (2) | 75% |
|  |  |  |  |  |  | 06/2020 |  |  |  |
| Dong^S45^ | SCL-90-R |  |  | First-year undergraduate students from a single university recruited online | China | 09/2019 | 4,085-4,341 | 19 (1) | 77% |
|  |  |  |  |  |  | NR/2020 |  |  |  |
| Elmer^S46^ |  | GAD-7 | CES-D | Undergraduate students in engineering and natural sciences from a single university recruited by email invitation | Switzerland | 09/2019 | 209 | NR | 22% |
|  |  |  |  |  |  | 04/2020 |  |  |  |
| Evans^S47^ | WEMWBS | HADS-A | HADS-D | First or second year undergraduate psychology students | UK | 10/2019 | 254 | 20 (1) | 86% |
|  |  |  |  |  |  | 05/2020 |  |  |  |
| Fuller-Rowell^S48^ |  |  | BDI-II | Undergraduate students at a four-year university in the southeastern United States | USA | 09/2018-04/2019 | 263 | 19 (1) | 53% |
|  |  |  |  |  |  | 04-06/2020 |  |  |  |
| Gelezelyte^S49^ |  | DASS-21 Anxiety | DASS-21 Depression | First-year university students | Lithuania | 10-12/2019 | 474 | 19 (1) | 76% |
|  |  |  |  |  |  | 10-12/2020 |  |  |  |
| Gopalan^S50^ |  | CCAPS-62 - Anxiety | CES-D-10 | Undergraduate students from a large, multicampus public university | USA | 11/2019 | 1,004 | 19 (1) | 62% |
|  |  |  |  |  |  | 05/2020 |  |  |  |
| Hamza^S51^ |  | GAD-7 | CES-D-R | Undergraduate students from single university recruited by email invitation | Canada | 05/2019 | 733 | 19 (1) | 74% |
|  |  |  |  |  |  | 05/2020 |  |  |  |
| He^S52^ |  | STAI-Trait |  | Individuals who took part in the Behavioral Brain Research Project of Chinese Personality | China | 09-12/2019 | 589 | 19 (1) | 71% |
|  |  |  |  |  |  | 02/2020 |  |  |  |
| Koelen^S53^ |  | GAD-7 | CES-D | Students at the University of Amsterdam | The Netherlands | 01/2019-01/2020^h^ | 671-683 | 23 (6) | 70% |
|  |  |  |  |  |  | 04-05/2020 |  |  |  |
| Li, H^S54^ | PHQ-4 |  |  | Undergraduate students from a single university enrolled in an ongoing longitudinal study | China | 12/2019 | 555 | 20 (3) | 77% |
|  | PANAS-PA |  |  |  |  |  |  |  |  |
|  | PANAS-NA |  |  |  |  | 02/2020 |  |  |  |
| Li, R^S55^ | SCL-90-R |  |  | Undergraduate students from multiple universities in Szechuan province recruited online | China | 09/2019 | 2,603 | NR | 53% |
|  |  |  |  |  |  | 04/2020 |  |  |  |
| Li, Wendy Wen^S56^ |  | DASS-21 Anxiety | DASS-21 Depression | Undergraduate students from single university recruited by email invitation | China | 11/2019 | 173 | 20 (1) | 78% |
|  |  |  |  |  |  | 03/2020 |  |  |  |
| Liu^S57^ |  |  | PHQ-9 | First-year students of two medical universities located in three cities (Jining, Weifang, Rizhao) | China | 04-10/2018-04-10/2019 | 5,373-8,079 | 18 (1) | 60% |
|  |  |  | CIDI 3.0 |  |  |  |  |  |  |
|  |  |  |  |  |  | 09-10/2020 |  |  |  |
| Lu^S58^ |  | GAD-7 | PHQ-9 | Students from Chinese Undergraduate Cohort study | China | 09-10/2019 | 5,181 | 14-17: 0.5%; 18-24: 99.5% | 62% |
|  |  |  |  |  |  | 04/2020 |  |  |  |
| Mauer^S59^ |  | DASS-21 |  | Undergraduate students from 11 American universities | USA | 09-12/2019 | 1,434 | 20 (1) | 76% |
|  |  |  |  |  |  | 03-06/2020 |  |  |  |
| Mehus^S60^ |  | GAD-7 | PHQ-9 | First year college students aged 18 to 23 years | USA | 08,12/2019 | 727 | 18 (94%)  19 (6%) | 64% |
|  |  |  |  |  |  | 04/2020 |  |  |  |
| Ratner^S61^ |  |  | BDI-II | Fourth-year university students from Cornell University | USA | 09/2019 | 152 | 21 (1) | 72% |
|  |  |  |  |  |  | 04/2020 |  |  |  |
| Saraswathi^S62^ |  | DASS-21 Anxiety | DASS-21 Depression | Convenience sample of undergraduate university medical students | India | 12/2019 | 217 | 20 (2) | 64% |
|  |  |  |  |  |  | 06/2020 |  |  |  |
| Savage, 2020^S63,i^ | WEMWBS |  |  | Undergraduate students from single university recruited by email invitation and enrolled in an ongoing longitudinal study | UK | 10/2019 | 214 | 18-21 (64) | 72% |
|  |  |  |  |  |  |  |  | 22-25 (22) |  |
|  |  |  |  |  |  | 04/2020 |  | 26-35 (8) |  |
|  |  |  |  |  |  |  |  | 35+ (6) |  |
| Savage, 2021^S64,i^ | WEMWBS |  |  | Undergraduate students from single university recruited by email invitation and enrolled in an ongoing longitudinal study | UK | 10/2019 | 255 | 18 (0)  19 (16)  20 (28)  21 (26)  22-25 (20)  26-35 (8)  35+ (2) | 76% |
|  |  |  |  |  |  | 10/2020 |  |  |  |
| Shiratori^S65^ |  |  | PHQ-9 | Students enrolled at the University of Tsukuba | Japan | NR/2019 | 6,847 | 23 (6) | 41% |
|  |  |  |  |  |  | 06/2020 |  |  |  |
| Truskauskaite-Kuneviciene^S66^ | PMH | DASS-21 - Anxiety | DASS-21 - Depression | Emerging adults studying at a large university in the Ruhr region (Germany) or Vilnius (Lithuania) | Lithuania Germany | 10-12/2019 | Lithuania: 450; Germany: 325 | Lithuania: 19 (1);  Germany: 23 (3) | Lithuania: 79; Germany: 78 |
|  |  |  |  |  |  | 03-04/2020 |  |  |  |
| Voltmer^S67^ |  | BSI-18 Anxiety | BSI-18 Depression | Students attending the University of Lübeck | Germany | NR/2019 | 890 | 24 (3) | 79% |
|  |  |  |  |  |  | 06/2020 |  |  |  |
| Wang, Yitao^S68^ | SCL-90-R | SCL-90-R Anxiety | SCL-90-R Depression | First-year students at a medical university | China | 11/2019 | 2,559 | NR | NR |
|  |  |  |  |  |  | 06/2020 |  |  |  |
| Yang, X^S69^ |  |  | CES-D | First-year students at Wenzhou Medical University in Zhejiang province | China | 12/2018 | 195 | NR | 59% |
|  |  |  |  |  |  | 06/2020 |  |  |  |
| Yang, Ziyan^S70^ |  | DASS-21 Anxiety | DASS-21 Depression | College students from Zhejiang Ocean University | China | 10/2019 | 2,364 | 20 (1) | 54% |
|  |  |  |  |  |  | 05/2020 |  |  |  |
| Zimmerman^S71^ |  | GAD-7 | PHQ-9 | Undergraduate students at a single university enrolled in a mental health prevention program study | USA | 08/2019 | 205 | 18 (1) | 76% |
|  |  |  |  |  |  | 04/2020 |  |  |  |
| **Children and Adolescents** | | | | | | | | | |
| Achterberg^S72^ | SDQ-Internalizing Behaviors |  |  | Children aged 10 to 13 years who enrolled in a longitudinal twin study in 2015-2016 | The Netherlands | 01-11/2019 | 151 | 12 (1) | 47% |
|  | SDQ-Externalizing Behaviors |  |  |  |  |  |  |  |  |
|  |  |  |  |  |  | 04-05/2020 |  |  |  |
| Adachi^S73^ |  |  | PHQ-A | Fourth to seventh graders from the Assessment of Preschool to Adolescence—Longitudinal Epidemiological study | Japan | 09/2019 | 4,118-4,126 | Range: 9-12 | 50% |
|  |  |  |  |  |  | 07/2020 |  |  |  |
| Bado^S74^ | SDQ - Total score |  |  | Primarily adolescents from the Brazilian High-Risk Cohort for Mental Conditions | Brazil | NR/2018-2019 | 672 | Mean: 19  Range: 16-24 | 53% |
|  | SDQ - Emotion subscale |  |  |  |  |  |  |  |  |
|  |  |  |  |  |  | 04/2020-04/2021 |  |  |  |
| Bernasco^S75^ | RCADS - Parent  RCADS - Adolescents |  |  | Urban young adolescents in their final year of primary school | The Netherlands | 09-12/2019 | 245 | 12 (1) | 50% |
|  |  |  |  |  |  | 04-07/2020 |  |  |  |
| Bosch^S76^ | SDQ - Emotion Symptoms SDQ - Total |  |  | Students in secondary education from INSchool study | Spain | NR/2019 | 552 | 15 (1) | 61% |
|  |  |  |  |  |  | 05-06/2020 |  |  |  |
| Charmaraman^S77^ |  |  | CES-D-R-10 | Students in grades 6 to 9 in two school districts in northeastern United States | USA | NR/2019 | 586 | 14 (1) | 53% |
|  |  |  |  |  |  | 10-12/2020 |  |  |  |
| Chen, I-H^S78^ | DASS-21 |  |  | Primary school students | China | 10-11/2019 | 535 | 10 (1) | 50% |
|  |  |  |  |  |  | 03/2020 |  |  |  |
| Chen, C-Y^S79^ | DASS-21 | DASS-21 - Anxiety | DASS-21 - Depression | Schoolchildren in grades 3 to 6 enrolled in 3 primary schools in Sichuan province | China | 10-11/2019 | 575 | 11 (1) | NR |
|  |  |  |  |  |  | 01/2020 |  |  |  |
| Daniunaite^S80^ | SDQ - Emotional Symptoms |  |  | Adolescents aged 12 to 16 years from the Stress and Resilience in Adolescence study | Lithuania | 03-05/2019 | 331 | 14 (2) | 57% |
|  |  |  |  |  |  | 09-10/2020 |  |  |  |
| Ezpeleta^S81^ | SDQ-total Parent Version |  |  | Families of children who were enrolled in a longitudinal cohort at age 3 (parents responded to measure of child mental health) | Spain | NR/2019 | 197 | 14 (0) | 52% |
|  |  |  |  |  |  | 06/2020 |  |  |  |
| Fujihara^S82^ | K6 |  |  | Japanese junior high school students | Japan | 12/2019 | 1,854 | NR | 51% |
|  |  |  |  |  |  | 02/2020 |  |  |  |
| Hu^S83^ | SDQ - Emotion Problems |  |  | Adolescents aged 10 to 16 years who took part in the Understanding Society survey | UK | NR | 886 | 13 (1) | 52% |
|  |  |  |  |  |  | 07/2020 |  |  |  |
| Knowles^S84^ | SDQ score | GAD-7 | SMFQ | Adolescents recruited from twelve local secondary schools. Participants were part of the Resilience, Ethnicity, and AdolesCent Mental Health cohort study | UK | NR/2018-2019 | 1,047 | Range: 12-18 | 55% |
|  |  |  |  |  |  | 05-08/2020 |  |  |  |
| Li, Y^S85^ |  | ZSAS | BDI-II | Students aged 14 to 19 years from three public commuter secondary vocational schools in Southern China province | China | 12/2019 | 831 | 16 (1) | 61% |
|  |  |  |  |  |  | 03/2020 |  |  |  |
| Liao^S86^ |  |  | CES-DC | Students from 3 junior high schools in Sichuan province | China | 12/2019 | 2,496 | 13 (1) | 50% |
|  |  |  |  |  |  | 07/2020 |  |  |  |
| Magson^S87^ |  | SCAS | SMFQ | Adolescents aged 13 to 16 years who were enrolled in a longitudinal cohort 4 years prior | Australia | NR/2019 | 248 | 14 (1) | 51% |
|  |  |  |  |  |  | 05/2020 |  |  |  |
| Mastorci^S88^ | KIDSCREEN-52 (psychological wellbeing) |  |  | Students aged 10 to 14 years | Italy | 09-10/2019 | 1,019 | 13 (1) | 52% |
|  | KIDSCREEN-52 (mood/emotion) |  |  |  |  | 04/2020 |  |  |  |
| Meireles^S89^ | KIDSCREEN-10 |  |  | Adolescents aged 12 to 16 years who attended schools in the north of Portugal | Portugal | 04-07/2019 | 1,099 | 13 (1) | 53% |
|  |  |  |  |  |  | 05-06/2020 |  |  |  |
| Naumann^S90^ |  |  | STDS | Adolescents and young adults from the German Family Panel Pairfam study | Germany | 11/2018-07/2019 | 854 | Range: 16-19 | 58% |
|  |  |  |  |  |  | 05-07/2020 |  |  |  |
| Paizan^S91^ | SWLS |  |  | 6th to 10th graders in schools with a high proportion of ethnic minorities | Germany | 06-10/2019 | 226 | 14 (1) | 56% |
|  |  |  |  |  |  | 05-07/2020 |  |  |  |
| Polack^S92^ |  |  | CDI-S | Individuals aged 9 to 15 years | USA | 01-09/2019 | 112 | 13 (2) | 55% |
|  |  |  |  |  |  | 03-06/2020 |  |  |  |
| Rau^S93^ | KIDSCREEN-10 | RCADS - Anxiety | RCADS - Depression | 5^th^ to 11th graders from 3 German schools | Germany | 10-11/2019 | 777 | 13 (2) | 53% |
|  |  |  |  |  |  | 06-07/2020 |  |  |  |
| Shoshani^S94^ | PANAS-C - PE | BSI-18 - Anxiety | BSI-18 - Depression | 5th to 11th graders from 38 schools in three representative geographical areas in Israel | Israel | 09/2019 | 1,537 | 14 (2) | 52% |
|  | PANAS-C - NE |  |  |  |  |  |  |  |  |
|  | GSI-18 - BSI |  |  |  |  | 05/2020 |  |  |  |
| Teng^S95^ |  | STAI-Trait | CES-D | Primary and middle school students | China | 10-11/2019 | 1,778 | NR | 49% |
|  |  |  |  |  |  | 04-05/2020 |  |  |  |
| Vira^S96^ | SDQ-emotional problems |  |  | Middle school students from the Peer Relations In School from an Ecological perspective project | Sweden | 10/2019-01/2020^h^ | 849 | 10 (0) | 52% |
|  |  |  |  |  |  | 11/2020-02/2021 |  |  |  |
| Wang, Wanxin^S97^ |  | GAD-7 | CES-D | 7th and 10th graders from six middle schools and four high schools in Guangzhou | China | 10-12/2019 | 1,790-1,831 | 14 (1) | 50% |
|  |  |  |  |  |  | 10-12/2020 |  |  |  |
| Widnall^S98^ |  | HADS-A |  | Secondary students aged 13 to 15 years in South West England | UK | 10/2019 | 603 | Range: 13-15 | NR |
|  |  |  |  |  |  | 05/2020 |  |  |  |
| Wong, R^S99^ |  | DASS-21 - Anxiety | DASS-21 - Depression | Young adolescents from the Healthy Kids cohort | China | 04-08/2019 | 233 | 12 (0) | 61% |
|  |  |  |  |  |  | 02/2020 |  |  |  |
| Yang, Zhengqian^S100^ |  |  | CES-D | Adolescents from three public junior high schools in Heilongjiang, who were part of the Life History Strategies and Adolescents' Adaptation Project | China | 11/2019 | 1,125 | 14 (1) | 51% |
|  |  |  |  |  |  | 08/2020 |  |  |  |
| Zhang^S101^ |  | HBQ | MFQ | Students in grades 4 through 8 enrolled in an ongoing longitudinal cohort | China | 11/2019 | 1,241 | 13 (1) | 59% |
|  |  |  |  |  |  | 05/2020 |  |  |  |
| **Parents** | | | | | | | | | |
| Achterberg^S72^ | BSI |  |  | Parents of children aged 10 to 13 years who enrolled in a longitudinal twin study in 2015-2016 | The Netherlands | 01-11/2019 | 106 | 45 (5) | 93% |
|  |  |  |  |  |  | 04-05/2020 |  |  |  |
| Adesogan^S102^ |  |  | CES-D | Black men and women in the rural South who took part in the Protecting Strong African American Families project. | USA | NR/2018-03/2020^h^ | 329 | 43 (8) | 58% |
|  |  |  |  |  |  | 06-09/2020 |  |  |  |
| Bosch^S76^ | SDQ - Emotion Symptoms SDQ - Total |  |  | Parents with children aged 6 to 17 from the INSchool study | Spain | NR/2019 | 669 | 13 (3) | 48% |
|  |  |  |  |  |  | 05-06/2020 |  |  |  |
| Frank^S103^ |  |  | PHQ-9 | Physician parents enrolled in the Intern Health Study | USA | 08/2018 | 180 | 40 (4) | 53% |
|  |  |  |  |  |  | 08/2020 |  |  |  |
| Gagné^S104^ | K10 |  |  | Parents with at least one child aged 5 to 17 years previously enrolled in a parenting support program | Canada | 03-05/2019 | 127 | NR | 80% |
|  |  |  |  |  |  | 05-07/2020 |  |  |  |
| Loret de Mola^S105^ |  | GAD-7 | EPDS | Mothers from the Rio Grande birth cohort | Brazil | 01-12/2019 | 1,136 | 28 (7) | 100% |
|  |  |  |  |  |  | 05-07/2020 |  |  |  |
| Pitchik^S106^ |  |  | CES-D | Primary caregivers of children 6 to 24 months with no physical or cognitive disabilities | Bangladesh | 05-06/2019 | 517 | NR (NR) | 100% |
|  |  |  |  |  |  | 07-09/2020 |  |  |  |
| Rivera^S107^ |  |  | EDS | Women enrolled in the Programming Research in Obesity, Growth, Environment and Social Stressors study | Mexico | NR/2018-2019 | 466 | 39 (6) | 100% |
|  |  |  |  |  |  | 05-11/2020 |  |  |  |
| Thompson^S108^ |  | GAD-7 | CES-D | First-time mothers of young toddlers living in low-income contexts | USA | NR/2018-2019 | 147 | 27 (6) | 100% |
|  |  |  |  |  |  | 04/2020 |  |  |  |
| **People with Pre-existing Medical Conditions** | | | | | | | | | |
| Becker^S109^ | SF-36 (role emotional) |  | CESD-10 | Individuals with multiple sclerosis | USA | 03/2019 | 119-121 | 69 (8) | 86% |
|  |  |  |  |  |  | 03/2020 |  |  |  |
| Bonenkamp^S110^ | SF-12 Mental Component Summary |  |  | Dialysis patients from the ongoing Dutch nOcturnal and hoMe dialysis Study To Improve Clinical Outcomes | The Netherlands | 08/2019 | 177 | 65 (12) | 37% |
|  |  |  |  |  |  | 07/2020 |  |  |  |
| Chao^S111^ |  |  | PHQ-8 | Older adults with Type 2 diabetes who were enrolled in Look AHEAD. | USA | 02/2018-02/2020^h^ | 2,679-2,829 | 76 (6) | 63% |
|  |  |  |  |  |  | 07-12/2020 |  |  |  |
| Chiu^S112^ |  | HADS-A | HADS-D | Individuals with Multiple Sclerosis | USA | 10/2018 | 133 | 49 (12) | 86% |
|  | SPANE-P |  |  |  |  |  |  |  |  |
|  | SPANE-N |  |  |  |  | 09/2020 |  |  |  |
| Derksen^S113^ | EORTC QLQ-C30-Global quality of life | HADS-A | HADS-D | Patients included in the nationwide Prospective Dutch Colorectal Cancer cohort | The Netherlands | 01/2019-01/2020^h^ | 2,176 | 67 (10) | 37% |
|  | EORTC QLQ-C30-Emotional functioning |  |  |  |  | 04-06/2020 |  |  |  |
| Dunlop-Thomas^S114^ | PROMIS - Global mental health |  | PROMIS - Depression | Patients with systemic lupus erythematosus who were part of the Georgians Organized Against Lupus (GOAL) cohort | USA | NR/2017-2019^g^ | 852 | 48 (NR) | 94% |
|  |  |  |  |  |  | NR/2020-2021 |  |  |  |
| Fujiwara^S115^ | EQ-5D-5L | HADS-A | HADS-D | Outpatients with chronic pain undergoing treatment | Japan | 07-09/2019 | 245 | 73 (12) | 55% |
|  |  |  |  |  |  | 07-09/2020 |  |  |  |
| García-Rudolph^S116^ | WHOQOL-BREF | HADS-A | HADS-D | Adult community residents with diagnosed spinal cord injury | Spain | NR | 175 | 55 (14) | 30% |
|  |  |  |  |  |  | 11/2020 |  |  |  |
| Gul^S117^ |  |  | BDI | Patients with epilepsy . | Turkey | 10-11/2019 | 116 | Median: 33 Range: 18-65 | 56% |
|  |  |  |  |  |  | 06-07/2020 |  |  |  |
| Henry^S118^ |  | PROMIS Anxiety | PHQ-8 | People with systemic sclerosis enrolled in an ongoing longitudinal cohort | Canada, France, UK, USA | 07-12/2019 | 435 | 57 (13) | 89% |
|  |  |  |  |  |  | 04/2020^j^ |  |  |  |
|  |  |  |  |  |  | 12/2020^j^ |  |  |  |
|  |  |  |  |  |  | 03/2021^j^ |  |  |  |
| Johnstone^S119^ | QOLS | HADS-A | HADS-D | Individuals with rheumatoid arthritis or ankylosing spondylitis from the Patient Opinion Real-Time Anonymous Liaison study | New Zealand | NR/2018 | 104 | 57 (12) | 74% |
|  |  |  |  |  |  | 07-09/2020 |  |  |  |
| Katz, P^S120^ |  | GAD-2 | PHQ-2 | People with rheumatic diseases enrolled in a longitudinal registry (National Databank for Rheumatic Diseases) | USA | NR/2019  03-06/2020 | 1,504 | 66 (11) | 86% |
| Liang^S121^ |  | ZSAS | ZSDS | Patients with maintenance hemodialysis under medical quarantine in a single hospital | China | 12/2019 | 114 | 59 (16) | 32% |
|  |  |  |  |  |  | 02-03/2020 |  |  |  |
| Lim^S122^ | PROMIS Mental Health | PROMIS Anxiety | PROMIS Depression | People with systemic lupus erythematosus | USA | NR/2018 | 316 | 47 (13) | 93% |
|  |  |  |  |  |  | 04/2020 |  |  |  |
| Möller^S123^ | DASS-21 |  |  | Adults with coeliac disease | Australia, New Zealand | 08-10/2019 | 674 | 57 (14) | 83% |
|  | EUROHIS-QOL |  |  |  |  | 05-07/2020 |  |  |  |
| Park^S124^ | EQ-5D-3L | HADS-A | HADS-D | Adults with pulmonary arterial hypertension who were part of the PEPPAH-study | Germany | 09/2019–02/2020^h^ | 152 | Median: 58 IQR: 49-67 | 73% |
|  |  |  |  |  |  | 05-08/2020 |  |  |  |
| Rentscher^S29^ |  | STAI-State | CES-D | Women aged 60 years and older who were nonmetastatic breast cancer survivors | USA | 02-06/2019 | 262 | 68 (5) | 100% |
|  |  |  |  |  |  | 05-09/2020 |  |  |  |
| Sacre^S125^ | PAID | GAD-7 | PHQ-8 | Adults with type 2 diabetes from the PREDICT cohort study | Australia | NR/2018-2019 | 450 | 66 (9) | 31% |
|  |  |  |  |  |  | 05-06/2020 |  |  |  |
| Sbragia^S126^ | HADS | HADS-A | HADS-D | Patients with multiple sclerosis | Italy | 01/2019 | 106 | 43 (11) | 70% |
|  |  |  |  |  |  | 05/2020 |  |  |  |
| Ubara^S127^ |  |  | PHQ-9 | Patients from a sleep outpatient clinic from a single hospital | Japan | 04-07/2019 | 164 | 64 (14) | 13% |
|  |  |  |  |  |  | 05/2020 |  |  |  |
| Uchida^S128^ |  |  | CES-D-SF | Hemodialysis patients | Japan | 04/2019-03/2020^h^ | 142 | 66 (11) | 42% |
|  |  |  |  |  |  | 07/2020-03/2021 |  |  |  |
| Wong, S^S35^ |  | GAD-7 | PHQ-9 | Adults aged ≥ 60 years with ≥ 2 chronic medical conditions recruited from 4 primary care clinics | Hong Kong, China | 04/2018-03/2019 | 583 | 71 (6) | 73% |
|  |  |  |  |  |  | 03-04/2020 |  |  |  |
|  |  |  |  |  |  | 03-06/2020 |  |  |  |
| **People with Pre-existing Mental Health Conditions** | | | | | | | | | |
| Gentile^S129^ |  | HAM-A | HAM-D | Psychiatric outpatients based in a large area of central-southern Italy and Department of Psychiatry of University of Asuncion, Paraguay. | Italy, Paraguay | 10-12/2019 | 110^k^ | 39(14) | 55% |
|  |  |  |  |  |  | 03-04/2020 |  |  |  |
| Huong^S130^ | BSRS-5 |  |  | Patients with treatment-refractory depression referred by two psychiatrists in the study hospitals | Taiwan | 01-12/2018 | 114 | 57 (14) | 71% |
|  |  |  |  |  |  | 01-05/2020 |  |  |  |
| Swerdlow^S131^ |  | MASQ-30 - Anxiety | MASQ-30 - Depression | A community sample of adults with pre-existing mental health concerns | USA | NR/2018-04/2020^h^ | 144 | 29 (NR) | 74% |
|  |  |  |  |  |  | 04-06/2020 |  |  |  |
| Young^S132^ |  | GAD-7 | PHQ-9 | UK residents aged ≥ 16 years with current or history of depressive or anxiety disorder diagnosis from the Genetic Links to Anxiety and Depression study | UK | 09/2018-02/2020^h^ | 12,653 | Range (%): 16-18 (3); 19-25 (12); 26-35 (23); 36-45 (19); 46-55 (22); 56-65 (15); 66-70 (3); 71-75 (2); 76+ (1) | 80% |
|  |  |  |  |  |  | 04-09/2020 |  |  |  |
| **Medical Staff** | | | | | | | | | |
| Frank^S103^ |  |  | PHQ-9 | Physician parents enrolled in the Intern Health Study | USA | 08/2018 | 180 | 40 (4) | 53% |
|  |  |  |  |  |  | 08/2020 |  |  |  |
| Li, Weidong^S133^ |  | GAD-7 | PHQ-9 | Training physicians from 12 Shanghai hospitals | China | 10-11/2019 | 385 | Median: 25 IQR: 23-28 | 64% |
|  |  |  |  |  |  | 01-02/2020 |  |  |  |
| **Sexual or Gender Minority Individuals** | | | | | | | | | |
| Bavinton^S134^ |  | GAD-7 | PHQ-9 | Gay and bisexual men enrolled in a longitudinal cohort | Australia | NR/2019 | 681 | NR | 0% |
|  |  |  |  |  |  | 04/2020 |  |  |  |
| Flentje^S135^ |  | GAD-7 | PHQ-9 | Convenience sample of sexual and gender minority adults enrolled in a longitudinal cohort | USA | 06/2019 | 2,288 | 37 (15) | 63%^l^ |
|  |  |  |  |  |  | 03-04/2020 |  |  |  |
| Ghabrial^S136^ |  | OASIS | CES-D | Trans and non-binary individuals who were part of the Trans PULSE Canada (TPC) study | Canada | NR/2019 | 780 | 33 (12) | 25% |
|  |  |  |  |  |  | 09-10/2020 |  |  |  |
| **Immigrants** | | | | | | | | | |
| Gosselin^S137^ |  |  | PHQ-9 | Immigrants from sub-Saharan Africa | France | 04/2018-NR/2019 | 100 | Range (%): 19-29 (34); 30-39 (39); 40+ (27) | 21% |
|  |  |  |  |  |  | 06/2020 |  |  |  |

BDI-II = Beck Depression Inventory (second edition); BJSQ = Brief Job Stress Questionnaire; BSI = Brief Symptom Inventory; BSRS = Brief-Symptom Rating Scale; CCAPS-62 = Counseling Center Assessment of Psychological Symptoms; CDI-S = Children’s Depression Inventory-Short; CES-DC = Center for Epidemiologic Studies Depression Scale Children; CES-D(-R/10) = Center for Epidemiologic Studies Depression (– Revised/10); CIDI = The Composite International Diagnostic Interview; DASS-21 = Depression, Anxiety, and Stress Scale; DToS = Distress Tolerance Scale; EDS = Edinburgh Depression Scale; EORTC QLQ-C30 = European Organization for the Research and Treatment of Cancer Quality of Life Questionnaire; EPDS = Edinburgh Postnatal Depression Scale; EQ-5D-5L = The 5-level European Quality of Life 5-dimensions version; GAD-7 = Generalized Anxiety Disorder; GAI-SF = Geriatric Anxiety Inventory - Short Form; GDS = Geriatric Depression Scale; GHQ = General Health Questionnaire; GSI = Global Severity Index; HADS-A/D = Hospital Anxiety and Depression Scale-Anxiety/Depression; HAI = Health Anxiety Inventory; HAM-A/D = Hamilton Anxiety/Depression Rating Scale; HBQ = MacArthur Health and Behavior Questionnaire; K6/10 = Kessler Psychological Distress Scale-6/10; MASQ = Mood and Anxiety Symptom Questionnaire; MDI = Major Depression Inventory; (S)MFQ = (Short) Mood and Feelings Questionnaire; MHI-5 = Mental Health Index-5; OASIS = Overall Anxiety Severity and Impairment Scale; PAID = Problem Areas in Diabetes scale; PANAS-PA/NA = Positive and Negative Affect Schedule – Positive Affect/Negative Affect; PERMA – PA/NA = Positive emotion, Engagement, Relationships, Meaning, and Accomplishment Profiler – Positive/Negative Affect; PHQ-2/4/8/9/A = Patient Health Questionnaire 2/4/8/9/Adolescents; PMH = Positive Mental Health Scale; PROMIS = Patient-Reported Outcomes Measurement Information System; PWBS = Psychological Well-being scale; QOLS = Quality of Life Scale; RCADS = Revised Children's Anxiety and Depression Scale; RPOMIS = Patient-Reported Outcomes Measurement Information System; RRQ = Reflection and Rumination Scale; SCAS = Spence Children’s Anxiety Scale; SCL-90-R = Symptom Check List-90-Revised; SDQ = Strengths and Difficulties Questionnaire; SF-8/12/36(-MCS) = Short-Form-8/12/36 Health Survey (Mental Component Summary); SMGAD = Severity Measure for Generalized Anxiety Disorder; SPANE-P/N = Scale of Positive and Negative Experience-Positive/Negative; STAI = State-Trait Anxiety Inventory; STDS = State-Trait Depression Scale; SWLS = Satisfaction with Life Scale; UK = United Kingdom; USA = United States of America; (S)WEMWBS = (Short) Warwick Edinburgh Mental Wellbeing Scale; WHO-5-J = World Health Organization (Five) Wellbeing Index; WHOQOL-BREF/AGE = World Health Organization Quality of Life Questionnaire for Older adults; ZSAS = Zung Self-rating Anxiety Scale; ZSDS = Zung Self-rating Depression Scale.

^a^Studies with data on females or women and males or men are not listed separately because they represent subgroup data from other studies in the table. ^b^Based on 671 participants with data during COVID-19. ^c^Analyses compared COVID-19 symptom levels to preceding trends across multiple assessments. ^d^Number included in fixed effects regression analysis from where the majority of data were extracted. ^e^Age groups reported for Daly^S12^; for Pierce,^S11^ 16-24 = 9%, 25-34 = 11%, 35-44 = 16%, 45-54 = 20%, 55-69 = 29%, 70+ = 15%. ^f^Based on van der Velden.^S17^ ^g^Included because estimated that over 80% of pre-COVID-19 data would have been collected after January 01, 2018. ^h^Included because estimated that over 80% of pre-COVID-19 data would have been collected by December 31, 2019. ^i^Recruited participants from the same longitudinal cohort. Of cohort participants who completed pre-COVID-19 assessments, 946 agreed to be contacted again. 214^S63^ and 255^S64^ completed assessments during early 2020 and later 2020, but the authors did not report how many participants overlapped between COVID-19 assessments in 2020. ^j^Mental health assessments conducted at 15 time points during COVID-19. We have reported first (April 2020), last in 2020 (December 2020) and last in 2021 (March 2021). ^k^N = 60 from Italy and 50 from Paraguay (results not reported by country). ^l^Based on female sex assigned at birth; 12 gender categories listed in study.

**Supplementary Table 2. Risk of Bias and Adequacy of Methods and Reporting**

| **Author** | **Appropriate sample frame** | **Appropriate participant recruitment** | **Adequate sample size** | **Participants and setting adequately described** | **Adequate response rate and data analysis with sufficient coverage** | **Valid methods for identification of outcome variable** | **Standard, reliable outcome measurement** | **Appropriate statistical analysis** | **Adequate follow-up response rate/ appropriate management of low response rate** |
| --- | --- | --- | --- | --- | --- | --- | --- | --- | --- |
| **General Population** | | | | | | | | | |
| Bulbulia^S1^ | Yes | Yes | Yes | Yes | No | Yes | Yes | Yes | No |
| Castellini^S2^ | Unclear | No | Unclear | Yes | Unclear | Yes | Yes | Yes | Yes |
| Chan^S3^ | No | No | Yes | No | Unclear | Yes | Yes | Yes | Unclear |
| Finucane^S4^ | No | Yes | Yes | Yes | Unclear | Yes | Unclear | Yes | No |
| Ge^S5^ | No | No | Yes | No | Unclear | Yes | Yes | Yes | Unclear |
| Haliwa^S6^ | No | No | Unclear | Yes | Unclear | Yes | Yes | Yes | No |
| Kanbur^S7^ | No | Unclear | Yes | No | Unclear | Yes | Unclear | No | Unclear |
| Katz, B^S8^ | No | No | Yes | Yes | Unclear | Yes | Yes | Yes | Unclear |
| Latikka^S9^ | Unclear | Unclear | Yes | Yes | No | Yes | Yes | Yes | Yes |
| Megias-Robles^S10^ | Unclear | No | Unclear | No | Unclear | Yes | Yes | Yes | No |
| Pierce^S11^ | Yes | Yes | Yes | Yes | Unclear | Yes | Yes | Yes | Unclear |
| Daly^S12^ | Yes | Yes | Yes | Yes | Unclear | Yes | Yes | Yes | No |
| Shimura^S13^ | No | Unclear | Yes | No | Unclear | Yes | Yes | Yes | Yes |
| Soltanzadeh^S14^ | No | Yes | Yes | Yes | Yes | Yes | Unclear | Yes | Yes |
| Thygesen^S15^ | Yes | Unclear | Yes | Yes | No | Yes | Unclear | Yes | Unclear |
| van der Velden, 2020^S16^ | Yes | Yes | Yes | Yes | Yes | Yes | Yes | Yes | Yes |
| van der Velden, 2021^S17^ | Yes | Yes | Yes | Yes | Yes | Yes | Yes | Yes | Yes |
| Wanberg^S18^ | Yes | Yes | Yes | Yes | Unclear | Yes | Unclear | Yes | No |
| **Older Adults** | | | | | | | | | |
| Bartlett^S19^ | Yes | No | Yes | Yes | Unclear | Yes | Yes | Yes | No |
| Briggs^S20^ | Yes | Yes | Yes | Yes | Unclear | Yes | Yes | Yes | No |
| Creese^S21^ | Yes | No | Yes | Yes | No | Yes | Yes | Yes | Yes |
| Eliasen^S22^ | No | Yes | Yes | Yes | No | Yes | Yes | Yes | No |
| Herrera^S23^ | Yes | Yes | Yes | Yes | Unclear | Yes | No | No | No |
| Kera^S24^ | No | Unclear | Yes | No | Unclear | Yes | Yes | Yes | No |
| Kivi^S25^ | Yes | Unclear | Yes | Yes | No | Yes | Yes | Yes | No |
| Lee^S26^ | Yes | Yes | Yes | Yes | Unclear | Yes | Yes | No | Unclear |
| Martínez^S27^ | No | Unclear | Unclear | Yes | Unclear | Yes | Yes | Yes | Unclear |
| Okely^S28^ | No | No | Unclear | Yes | No | Yes | Yes | Yes | No |
| Rentscher^S29^ | Yes | Unclear | Yes | Yes | Unclear | Yes | No | Yes | Yes |
| Sardella^S30^ | No | No | Unclear | Yes | Unclear | Yes | Yes | Yes | No |
| Siew^S31^ | Yes | No | Yes | Yes | Unclear | Yes | No | Yes | No |
| van den Besselaar^S32^ | Unclear | No | Yes | Yes | Unclear | Yes | No | Yes | Unclear |
| van Tilburg^S33^ | Yes | Yes | Yes | Yes | Yes | Yes | Yes | Yes | Yes |
| Wang, Yi^S34^ | Yes | Yes | Yes | Yes | Unclear | Yes | Yes | Yes | Yes |
| Wong, S^S35^ | No | Unclear | Yes | Yes | Unclear | Yes | Yes | Yes | Yes |
| Yu^S36^ | Yes | No | Yes | Yes | Unclear | Yes | Yes | Yes | No |
| **Young Adults** | | | | | | | | | |
| Islam^S37^ | No | Yes | Yes | Yes | No | Yes | Yes | Yes | No |
| Marmet^S38^ | Yes | Yes | Yes | Yes | Unclear | Yes | Yes | Yes | No |
| Rimfeld^S39^ | Yes | Unclear | Yes | Yes | Unclear | Yes | Yes | Yes | Unclear |
| Romm^S40^ | No | No | Yes | Yes | Unclear | Yes | Yes | Yes | Unclear |
| Tanioka^S41^ | No | Unclear | Yes | Yes | Unclear | Yes | Yes | Yes | No |
| Villadsen^S42^ | Unclear | Unclear | Yes | Yes | Unclear | Yes | Yes | Yes | No |
| Watkins-Martin^S43^ | Yes | Unclear | Yes | Yes | Unclear | Yes | Yes | Yes | No |
| **University Students** | | | | | | | | | |
| Conceição^S44^ | No | Yes | Yes | Yes | Unclear | Yes | Unclear | Yes | No |
| Dong^S45^ | No | No | Yes | Yes | Yes | Yes | Yes | Yes | Yes |
| Elmer^S46^ | No | Yes | Yes | Yes | Unclear | Yes | Yes | Yes | Unclear |
| Evans^S47^ | No | Unclear | Yes | Yes | Unclear | Yes | No | Yes | Yes |
| Fuller-Rowell^S48^ | No | Unclear | Yes | Yes | Unclear | Yes | Unclear | Yes | Yes |
| Gelezelyte^S49^ | No | Unclear | Yes | Yes | No | Yes | Yes | Yes | No |
| Gopalan^S50^ | No | Unclear | Yes | Yes | Unclear | Yes | Yes | Yes | No |
| Hamza^S51^ | No | Unclear | Yes | Yes | Unclear | Yes | Yes | Yes | Yes |
| He^S52^ | Unclear | Unclear | Yes | Yes | Unclear | Yes | Unclear | Yes | Yes |
| Koelen^S53^ | No | Yes | Yes | Yes | Unclear | Yes | Yes | Yes | No |
| Li, H^S54^ | No | Unclear | Yes | Yes | Unclear | Yes | Yes | Yes | Yes |
| Li, R^S55^ | No | Yes | Yes | No | Unclear | Yes | Yes | Yes | Yes |
| Li, Wendy Wen^S56^ | No | No | Unclear | Yes | No | Yes | Yes | Yes | Yes |
| Liu^S57^ | No | Yes | Yes | Yes | Yes | Yes | Yes | Yes | Yes |
| Lu^S58^ | Unclear | Yes | Yes | Yes | Unclear | Yes | Unclear | Yes | Unclear |
| Mauer^S59^ | Yes | No | Yes | Yes | Unclear | Yes | Yes | Yes | No |
| Mehus^S60^ | No | Yes | Yes | Yes | Unclear | Yes | Yes | Yes | Unclear |
| Ratner^S61^ | No | Unclear | Unclear | Yes | Unclear | Yes | Yes | Yes | No |
| Saraswathi^S62^ | No | Yes | Unclear | Yes | Yes | Yes | Yes | Yes | Yes |
| Savage, 2020^S63^ | No | Yes | Yes | Yes | No | Yes | Yes | Yes | No |
| Savage, 2021^S64^ | Unclear | Unclear | Yes | Yes | No | Yes | Yes | No | No |
| Shiratori^S65^ | No | Unclear | Yes | Yes | Unclear | Yes | Yes | Yes | No |
| Truskauskaite-Kuneviciene^S66^ | No | Unclear | Yes | Yes | Unclear | Yes | Yes | Yes | No |
| Voltmer^S67^ | No | No | Yes | Yes | No | Yes | Yes | Yes | Unclear |
| Wang, Yitao^S68^ | No | Yes | Yes | No | Unclear | Yes | Yes | Yes | Yes |
| Yang, X^S69^ | No | Unclear | Unclear | No | Unclear | Yes | Yes | Yes | Unclear |
| Yang, Ziyan^S70^ | No | Unclear | Yes | Yes | Unclear | Yes | Unclear | Yes | Unclear |
| Zimmerman^S71^ | No | No | Yes | Yes | Unclear | Yes | Yes | Yes | No |
| **Children and Adolescents** | | | | | | | | | |
| Achterberg^S72,a^ | No | No | Unclear | Yes | Unclear | Yes | No | Yes | No |
| Adachi^S73^ | No | Unclear | Yes | Yes | Yes | Yes | Yes | Yes | Yes |
| Bado^S74^ | No | No | Yes | Yes | Unclear | Yes | Unclear | Yes | Unclear |
| Bernasco^S75^ | No | Unclear | Yes | Yes | Unclear | Yes | Yes | Yes | Unclear |
| Bosch^S76^ | Yes | Unclear | Yes | Yes | No | Yes | Yes | Yes | No |
| Charmaraman^S77^ | No | Unclear | Yes | Yes | Unclear | Yes | Unclear | Yes | No |
| Chen, I-H^S78^ | No | No | Yes | Yes | Yes | Yes | Yes | Yes | Yes |
| Chen, C-Y^S79^ | No | Unclear | Yes | No | Yes | Yes | Yes | Yes | No |
| Daniunaite^S80^ | No | Yes | Yes | Yes | Unclear | Yes | Yes | Yes | Yes |
| Ezpeleta^S81^ | No | No | Unclear | Yes | No | Yes | No | Yes | Unclear |
| Fujihara^S82^ | No | Unclear | Yes | No | No | Yes | Yes | Yes | Unclear |
| Hu^S83^ | Unclear | Unclear | Yes | Yes | Unclear | Yes | Yes | Yes | Unclear |
| Knowles^S84^ | Unclear | Unclear | Yes | Yes | No | Yes | Yes | Yes | Unclear |
| Li, Y^S85^ | No | Unclear | Yes | Yes | Unclear | Yes | Yes | Yes | Yes |
| Liao^S86^ | No | Yes | Yes | Yes | Unclear | Yes | Unclear | Yes | Yes |
| Magson^S87^ | No | Unclear | Yes | Yes | Unclear | Yes | Yes | Yes | No |
| Mastorci^S88^ | No | Yes | Yes | Yes | Unclear | Yes | Yes | Yes | No |
| Meireles^S89^ | No | Unclear | Yes | Yes | Unclear | Yes | Yes | Yes | Unclear |
| Naumann^S90^ | Yes | Unclear | Yes | Yes | Unclear | Yes | Yes | Yes | No |
| Paizan^S91^ | No | No | Yes | Yes | Unclear | Yes | Yes | Yes | Unclear |
| Polack^S92^ | No | No | Unclear | No | Unclear | Yes | Yes | Yes | Yes |
| Rau^S93^ | No | Unclear | Yes | Yes | No | Yes | Yes | Yes | No |
| Shoshani^S94^ | No | Unclear | Yes | Yes | Unclear | Yes | Yes | Yes | Yes |
| Teng^S95^ | No | Yes | Yes | Yes | Unclear | Yes | Yes | Yes | Yes |
| Vira^S96^ | Unclear | Unclear | Yes | Yes | Unclear | Yes | No | Yes | Unclear |
| Wang, Wanxin^S97^ | No | Yes | Yes | Yes | Yes | Yes | Unclear | Yes | Yes |
| Widnall^S98^ | No | Unclear | Yes | No | Unclear | Yes | Unclear | Yes | Unclear |
| Wong, R^S99^ | Unclear | Unclear | Yes | Yes | Unclear | Yes | Yes | Yes | Unclear |
| Yang, Zhengqian^S100^ | No | Unclear | Yes | Yes | Unclear | Yes | Yes | Yes | Unclear |
| Zhang^S101^ | No | No | Yes | Yes | Yes | Yes | Yes | Yes | Yes |
| **Parents** | | | | | | | | | |
| Achterberg^S72,a^ | No | No | Unclear | Yes | Unclear | Yes | Yes | Yes | No |
| Adesogan^S102^ | No | No | Yes | Yes | Unclear | Yes | No | Yes | Yes |
| Bosch^S76^ | Yes | Unclear | Yes | Yes | No | Yes | Yes | Yes | No |
| Frank^S103^ | No | Unclear | Unclear | Yes | Unclear | Yes | Yes | Yes | Unclear |
| Gagné^S104^ | Unclear | Unclear | Unclear | No | Unclear | Yes | Yes | Yes | Yes |
| Loret de Mola^S105^ | No | No | Yes | Yes | Unclear | Yes | Yes | Yes | No |
| Pitchik^S106^ | No | Yes | Yes | No | Unclear | Yes | Yes | Yes | No |
| Rivera^S107^ | No | Unclear | Yes | Yes | Unclear | Yes | Unclear | Yes | Unclear |
| Thompson^S108^ | Yes | Unclear | Unclear | Yes | Unclear | Yes | Yes | Yes | Yes |
| **People with Pre-existing Medical Conditions** | | | | | | | | | |
| Becker^S109^ | Yes | No | Yes | Yes | No | Yes | Yes | Yes | Unclear |
| Bonenkamp^S110^ | Yes | Unclear | Yes | Yes | Unclear | Yes | Unclear | Yes | Unclear |
| Chao^S111^ | No | Unclear | Yes | Yes | Unclear | Yes | Yes | Yes | Unclear |
| Chiu^S112^ | No | No | Unclear | Yes | Unclear | Yes | Yes | Yes | No |
| Derksen^S113^ | Yes | Unclear | Yes | Yes | Unclear | Yes | Unclear | Yes | No |
| Dunlop-Thomas^S114^ | Yes | Unclear | Yes | Yes | Unclear | Yes | Yes | Yes | Unclear |
| Fujiwara^S115^ | No | Unclear | Yes | No | Unclear | Yes | Unclear | Yes | Unclear |
| García-Rudolph^S116^ | No | No | Unclear | Yes | Unclear | Yes | Unclear | Yes | Yes |
| Gul^S117^ | No | Unclear | Unclear | Yes | Unclear | Yes | Unclear | Yes | Unclear |
| Henry^S118^ | Yes | No | Yes | Yes | Unclear | Yes | Yes | Yes | No |
| Johnstone^S119^ | No | No | Unclear | No | Unclear | Yes | Yes | Yes | Yes |
| Katz, P^S120^ | Unclear | Unclear | Yes | Yes | Unclear | Yes | Yes | Yes | No |
| Liang^S121^ | No | No | Unclear | Yes | Yes | Yes | Yes | Yes | Yes |
| Lim^S122^ | Yes | Unclear | Yes | Yes | Unclear | Yes | Unclear | Yes | Unclear |
| Möller^S123^ | Yes | No | Yes | Yes | No | Yes | Yes | Yes | Yes |
| Park^S124^ | No | No | Unclear | No | Unclear | Yes | Yes | Yes | Yes |
| Rentscher^S29^ | Yes | Unclear | Yes | Yes | Unclear | Yes | No | Yes | Yes |
| Sacre^S125^ | No | No | Yes | Yes | Unclear | Yes | No | Yes | Yes |
| Sbragia^S126^ | No | No | Unclear | Yes | Unclear | Yes | Unclear | Yes | No |
| Ubara^S127^ | No | Unclear | Unclear | No | Unclear | Yes | Yes | Yes | Unclear |
| Uchida^S128^ | No | Unclear | Unclear | Yes | Unclear | Yes | Unclear | Yes | Unclear |
| Wong, S^S35^ | No | Unclear | Yes | Yes | Unclear | Yes | Yes | Yes | Yes |
| **People with Pre-existing Mental Health Conditions** | | | | | | | | | |
| Gentile^S129^ | Yes | Unclear | Unclear | Yes | Unclear | Yes | No | Yes | Unclear |
| Huong^S130^ | No | Unclear | Unclear | Yes | Unclear | Yes | Yes | Yes | Yes |
| Swerdlow^S131^ | Yes | No | Unclear | Yes | Unclear | Yes | No | Yes | Yes |
| Young^S132^ | Yes | No | Yes | Yes | Unclear | Yes | Yes | Yes | No |
| **Medical Staff** | | | | | | | | | |
| Frank^S103^ | No | Unclear | Unclear | Yes | Unclear | Yes | Yes | Yes | Unclear |
| Li, Weidong^S133^ | No | Unclear | Yes | Yes | No | Yes | Yes | Yes | No |
| **Sexual or Gender Minority Individuals** | | | | | | | | | |
| Bavinton^S134^ | Yes | Unclear | Yes | No | Unclear | Yes | Yes | Yes | Unclear |
| Flentje^S135^ | Yes | No | Yes | Yes | Unclear | Yes | Yes | Yes | Unclear |
| Ghabrial^S136^ | Unclear | Unclear | Yes | Yes | Unclear | Yes | No | Yes | Unclear |
| **Immigrants** | | | | | | | | | |
| Gosselin^S137^ | No | No | Unclear | Yes | No | Yes | Yes | Yes | Unclear |

^a^Achterberg et al. has two samples, parents, and their children, with independent risk of bias coding.

**Supplementary Table 3. Individual Study Results for General Mental Health**

| **First Author**  **Study Country** | **Pre- and Post-COVID-19 Data Collection** | **N** | **Continuous**  **Outcome Measure** | **Pre-**  **COVID-19 Mean (SD)** | **Post-**  **COVID-19 Mean (SD)** | **Mean (SD) Change**^a^ | **Hedges’ g Standardized Mean Difference (95% CI)** | **Dichotomous Outcome Measure** | **% pre-COVID-19 (95% CI)** | **% post-COVID-19 (95% CI)** | **% Change with 95% CI**^a^ |
| --- | --- | --- | --- | --- | --- | --- | --- | --- | --- | --- | --- |
| **General Population** | | | | | | | | | | | |
| Bulbulia^S1^  New Zealand | NR/2018 | 940 | K6 | 5.42 (4.05) | 5.65 (3.78) | 0.23 (NR) | 0.06 (-0.03, 0.15) | ---------- | ---------- | ---------- | ---------- |
|  | 03-04/2020 |  |  |  |  |  |  |  |  |  |  |
| Castellini^S2^  Italy | 12/2019 | 130 | BSI-GSI | 0.51 (0.39) | 0.46 (0.46) | -0.05 (NR) | -0.12 (-0.36, 0.13) | ---------- | ---------- | ---------- | ---------- |
|  | 04-05/2020 |  |  |  |  |  |  |  |  |  |  |
| Finucane^S4^  USA | 05/11-2018 | 416 | K6 | 4.00 (4.40) | 4.70 (4.60) | 0.70 (NR) | 0.16 (0.02, 0.29) | K6 ≥ 13 | 6.0 (4.1, 8.7) | 6.5 (4.5, 9.2) | 0.5 (-2.0, 3.0) |
|  | 06-09/2020 |  |  |  |  |  |  |  |  |  |  |
| Kanbur^S7^  Turkey | NR/2019 | 400 | SCL-90-R | 0.36 (NR) | 0.78 (NR) | 0.42 (NR) | NR (NR) | ---------- | ---------- | ---------- | ---------- |
|  | NR/2020 |  |  |  |  |  |  |  |  |  |  |
| Katz, B^S8^  Canada, Ireland, UK, USA | 04/2019 | 218 | RRQ | 42.46 (11.74) | 41.66 (12.15) | -0.80 (7.43) | -0.07 (-0.25, 0.12) | ---------- | ---------- | ---------- | ---------- |
|  | 04/2020 |  | DToS | 41.95 (9.95) | 41.03 (10.08) | -0.92 (7.63) | -0.09 (-0.28, 0.10) | ---------- | ---------- | ---------- | ---------- |
| Latikka^S9^  Finland | 09/2019-10/2019 | 840 | GHQ-12 | 12.20 (5.67) | 12.41 (5.45) | 0.21 (NR) | 0.04 (-0.06, 0.13) | ---------- | ---------- | ---------- | ---------- |
|  | 03/2020-04/2020 |  |  |  |  |  |  |  |  |  |  |
| Megias-Robles^S10^  Spain | 11/2019 | 102 | PANAS-NA | 1.92 (0.65) | 2.22 (0.75) | 0.31 (0.80) | 0.44 (0.16, 0.71) | ---------- | ---------- | ---------- | ---------- |
|  | 04/2020 |  |  |  |  |  |  |  |  |  |  |
| Pierce^S11^  UK | Pre-COVID-19 waves | 15,376^30,b^ 10,918^34^ | GHQ-12 |  |  |  |  | GHQ-12 ≥ 4 |  |  |  |
| Daly^S12^  UK |  |  |  |  |  |  |  |  |  |  |  |
|  | 04/2020 |  |  | 11.50 (5.50) | 12.60 (6.60) | 1.10 (NR)^c^ | 0.18 (0.16, 0.21) |  | 20.8 (19.4, 22.2)^e^ | 29.5 (28.0, 31.0)^e^ | 8.7 (6.9, 10.4)^e^ |
|  |  |  |  |  |  | 0.48 (NR)^d^ | 0.08 (0.05, 0.10) |  |  |  |  |
|  | 09/2020 |  |  | ---------- | ---------- | ---------- | ---------- |  | 20.8 (19.4, 22.2)^e^ | 20.8 (19.5, 22.1)^e^ | 0.0 (-2.0, 1.9)^e^ |
| Shimura^S13^  Japan | NR/2019 | 3,123 | BJSQ (Psychological and Physical) | NR (NR) | NR (NR) | -0.31 (11.02) | -0.05 (-0.10, 0.00) | ---------- | ---------- | ---------- | ---------- |
|  | NR/2020 |  |  |  |  |  |  |  |  |  |  |
| Soltanzadeh^S14^  Iran | 11/2019 | 823 | GHQ-28 | 45.13 (11.65) | 51.41 (12.89) | 6.28 (NR) | 0.51 (0.41, 0.61) | ---------- | ---------- | ---------- | ---------- |
|  | 07/2020 |  |  |  |  |  |  |  |  |  |  |
| Thygesen^S15^  Denmark | 09-12/2019 | 4,234 | SWEMWBS | 25.50 (4.98) | 24.60 (4.98) | -0.90 (NR) | 0.18 (0.14, 0.22) | ---------- | ---------- | ---------- | ---------- |
|  | 09-11/2020 |  |  |  |  |  |  |  |  |  |  |
| van der Velden, 2020^S16^  Netherlands | 03/2019 | 3,983 | MHI-5^f^ |  |  |  |  | MHI-5 ≤ 59 |  |  |  |
| van der Velden, 2021^S17^  Netherlands | 11-12/2019 | 4,064 |  |  |  |  |  |  |  |  |  |
|  | 03/2020 |  |  | 74.20 (16.70) | 74.10 (16.40) | -0.10 (NR) | 0.01 (-0.04, 0.05) |  | ---------- | ---------- | ---------- |
|  | 11-12/2020 |  |  | ---------- | ---------- | ---------- | ---------- |  | 16.9 (15.8, 18.1) | 16.9 (15.8, 18.1) | 0.0 (-1.2, 1.3) |
| **Older Adults** | | | | | | | | | | | |
| Eliasen^S22^  Denmark | 12/2017-01/2019 | 225 | WHOQOL-BREF | 74.33 (14.96) | 71.88 (15.21) | -2.45 (NR) | 0.16 (-0.02, 0.35) | ---------- | ---------- | ---------- | ---------- |
|  |  |  | WHOQOL-BREF (psychological health) | 77.07 (11.52) | 80.53 (10.89) | 3.46 (NR) | -0.31 (-0.49, -0.12) |  |  |  |  |
|  | 06/2020-07/2020 |  |  |  |  |  |  |  |  |  |  |
| Kera^S24^  Japan | 10/2019 | 533 | WHO-5-J | 16.70 (4.79) | 15.10 (4.79) | -1.60 (NR) | 0.33 (0.21, 0.45) | ---------- | ---------- | ---------- | ---------- |
|  | 06-07/2020 |  |  |  |  |  |  |  |  |  |  |
| Kivi^S25^  Sweden | NR/2019 | 1,071 | SWLS^f^ | 5.12 (1.30) | 5.16 (1.26) | 0.04 (NR) | -0.03 (-0.12, 0.05) | ---------- | ---------- | ---------- | ---------- |
|  | 03-04/2020 |  |  |  |  |  |  |  |  |  |  |
| Martínez^S27^  Spain | 10/2019 | 141 | PWBS | 97.85 (21.30) | 99.50 (19.30) | 1.65 (11.53) | -0.08 (-0.32, 0.15) | ---------- | ---------- | ---------- | ---------- |
|  |  |  | PERMA - PA | 7.27 (1.38) | 7.20 (1.40) | -0.07 (1.14) | 0.05 (-0.18, 0.28) |  |  |  |  |
|  | 04/2020 |  | PERMA - NA | 4.12 (1.50) | 3.90 (1.60) | -0.22 (1.22) | -0.14 (-0.38, 0.09) |  |  |  |  |
| Okely^S28^  Scotland (UK) | NR/2017-NR/2019 | 137 | WEMWBS^f^ | 37.45 (8.37) | 36.45 (8.23) | -1.00 (NR) | 0.12 (-0.12, 0.36) | ---------- | ---------- | ---------- | ---------- |
|  | 05-06/2020 |  |  |  |  |  |  |  |  |  |  |
| Pierce^S11^  UK | Pre-COVID-19 waves | 2,491 (≥70 years)^30,b^ 3,447 (≥65 years)^34^ | GHQ-12 |  |  |  |  | GHQ-12 ≥ 4 |  |  |  |
| Daly^S12^  UK |  |  |  |  |  |  |  |  |  |  |  |
|  | 04/2020 |  |  | 10.10 (4.57) | 10.90 (5.35) | 0.80 (NR)^c^ | 0.16 (0.11, 0.21) |  | 12.7 (10.3, 15.1)^e^ | 19.4 (17.1, 21.8)^e^ | 6.8 (3.7, 9.8)^e^ |
|  |  |  |  |  |  | 0.05 (NR)^d^ | 0.01 (-0.04, 0.06) |  |  |  |  |
|  | 09/2020 |  |  | ---------- | ---------- | ---------- | ---------- |  | 12.7 (10.3, 15.1)^e^ | 14.9 (12.9, 16.9)^e^ | 2.2 (-0.8, 5.2)^e^ |
| Sardella^S30^  Italy | 10/2018-10/2019 | 104 | SF-12 Mental Component Summary | 49.99 (9.99) | 46.35 (10.06) | -3.64 (NR) | -0.36 (-0.64, -0.09) | ---------- | ---------- | ---------- | ---------- |
|  | 04/2020 |  |  |  |  |  |  |  |  |  |  |
| Siew^S31^  Singapore | 02/2018-01/2020 | 411 | WHOQOL-AGE | 50.36 (5.71) | 52.19 (6.42) | 1.83 (NR) | -0.30 (-0.44, -0.16) | ---------- | ---------- | ---------- | ---------- |
|  | 05-06/2020 |  |  |  |  |  |  |  |  |  |  |
| Thygesen^S15^  Denmark | 09-12/2019 | 423 | SWEMWBS | 25.70 (6.82) | 25.00 (5.77) | -0.70 (NR) | 0.11 (-0.02, 0.25) | ---------- | ---------- | ---------- | ---------- |
|  | 09-11/2020 |  |  |  |  |  |  |  |  |  |  |
| van der Velden, 2020^S16^  Netherlands | 03/2019 | 949-1,038 |  |  |  |  |  |  |  |  |  |
| van der Velden, 2021^S17^  Netherlands | 11-12/2019 | 968-1,052 |  |  |  |  |  |  |  |  |  |
| van Tilburg^S33^  Netherlands |  | 1,679 |  |  |  |  |  |  |  |  |  |
|  | 03/2020^36^ |  |  |  |  |  |  | MHI-5 ≤ 59 | 10.9 (9.0, 13.0) | 10.6 (8.9, 12.6) | -0.2 (-2.3, 1.9) |
|  | 05/2020^41^ |  |  | 4.93 (0.75) | 5.02 (0.73) | 0.09 (0.58) | -0.12 (-0.19, -0.05) | ---------- | ---------- | ---------- | ---------- |
|  | 11-12/2020^37^ |  |  |  |  |  |  | MHI-5 ≤ 59 | 12.1 (10.2, 14.3)^g^ | 10.5 (8.8, 12.5)^g^ | -1.7 (-3.7, 0.4) |
| Wang, Yi^S34^  China | 05-06/2019 | 2,745 | K10 | 16.64 (7.44) | 18.23 (8.06) | 1.35 (6.15) | 0.17 (0.12, 0.23) | ---------- | ---------- | ---------- | ---------- |
|  | 08-09/2020 |  |  |  |  |  |  |  |  |  |  |
| **Young Adults** | | | | | | | | | | | |
| Islam^S37^  Australia | NR/2018 | 1,110 | K10 | ---------- | ---------- | ---------- | ---------- | K10 ≥ 25 | 41.3 (38.4, 44.2) | 41.3 (38.4, 44.2) | 0.0 (-3.8, 3.8) |
|  | 10-12/2020 |  |  |  |  |  |  |  |  |  |  |
| Pierce^S11^  UK | Pre-COVID-19 waves | 1,999 (25-34 years)^30,b^ 1,260 (18-34 years)^34^ | GHQ-12 |  |  |  |  | GHQ-12 ≥ 4 |  |  |  |
| Daly^S12^  UK |  |  |  |  |  |  |  |  |  |  |  |
|  | 04/2020 |  |  | 12.10 (5.46) | 14.20 (6.32) | 2.10 (NR)^c^ | 0.36 (0.29, 0.42) |  | 25.4 (21.6, 29.2)^e^ | 39.9 (35.5, 44.4)^e^ | 14.5 (9.6,19.4)^e^ |
|  |  |  |  |  |  | 1.61 (NR)^d^ | 0.27 (0.21, 0.34) |  |  |  |  |
|  |  |  |  | ---------- | ---------- |  |  |  |  |  |  |
|  | 09/2020 |  |  |  |  | ---------- | ---------- |  | 25.4 (21.6, 29.2)^e^ | 23.7 (19.8, 27.6)^e^ | -1.7 (-5.9, 2.5)^e^ |
| Tanioka^S41^  Japan | 10/2019 | 2,222 | K6-J | 6.10 (5.70) | 6.10 (5.80) | 0.00 (NR) | 0.00 (-0.06, 0.06) | ---------- | ---------- | ---------- | ---------- |
|  |  |  | SF-8 - MCS | 46.60 (7.90) | 47.20 (7.80) | 0.60 (NR) | 0.08 (0.02, 0.14) |  |  |  |  |
|  | 05/2020 |  |  |  |  |  |  |  |  |  |  |
| van der Velden, 2020^S16^  Netherlands | 03/2019 | 993-1,062 | ---------- | ---------- | ---------- | ---------- | ---------- | MHI-5 ≤ 59 |  |  |  |
| van der Velden, 2021^S17^  Netherlands | 11-12/2019 | 1,018-1,083 |  |  |  |  |  |  |  |  |  |
|  | 03/2020 |  |  |  |  |  |  |  | 23.1 (20.6, 25.7) | 19.7 (17.4, 22.3) | -3.3 (-6.1, -0.6) |
|  | 11-12/2020 |  |  |  |  |  |  |  | 20.7 (18.4, 23.2)^g^ | 22.5 (20.0, 25.2)^g^ | 1.8 (-1.1, 4.7) |
| Villadsen^S42^  UK | NR/2018 | 1,615 | K6 | ---------- | ---------- | ---------- | ---------- | K6 ≥ 13 | 18.0 (15.2, 21.2) | 18.7 (15.7, 22.1) | 0.7 (-1.4, 2.8) |
|  | 05/2020 |  |  |  |  |  |  |  |  |  |  |
| **University Students** | | | | | | | | | | | |
| Dong^S45^  China | 09/2019 | 4,085-4,341 | ---------- | ---------- | ---------- | ---------- | ---------- | SCL-90-R ≥ 160 | 18.4 (17.3, 19.6) | 26.4 (25.1, 27.8) | 8.0 (6.4, 9.5) |
|  | NR/2020 |  |  |  |  |  |  |  |  |  |  |
| Evans^S47^  UK | 10/2019 | 251 | WEMWBSf | 23.04 (4.96) | 21.12 (4.87) | -1.92 (NR) | 0.39 (0.21, 0.57) | ---------- | ---------- | ---------- | ---------- |
|  | 05/2020 |  |  |  |  |  |  |  |  |  |  |
| Li, H^S54^  China | 12/2019 | 555 | PHQ-4 | 0.95 (0.65) | 0.76 (0.61) | -0.19 (0.66) | -0.30 (-0.42, -0.18) | ---------- | ---------- | ---------- | ---------- |
|  | 02/2020 |  | PANAS- PA^f^ | 3.21 (0.79) | 3.26 (0.79) | 0.06 (0.78) | -0.08 (-0.19, 0.04) |  |  |  |  |
|  |  |  | PANAS- NA | 2.38 (0.79) | 2.24 (0.80) | -0.15 (0.78) | -0.19 (-0.31, -0.07) |  |  |  |  |
| Li, R^S55^  China | 09/2019 | 2,603 | SCL-90-R | 1.60 (0.40) | 1.52 (0.41) | -0.08 (0.66) | -0.20 (-0.25, -0.14) | ---------- | ---------- | ---------- | ---------- |
|  | 04/2020 |  |  |  |  |  |  |  |  |  |  |
| Savage, 2020^S63^  UK | 10/2019 | 214 | WEMWBS^f^ | 44.12 (9.16) | 41.12 (10.14) | -3.00 (NR) | 0.31 (0.12, 0.50) | ---------- | ---------- | ---------- | ---------- |
|  | 04/2020 |  |  |  |  |  |  |  |  |  |  |
| Savage, 2021^S64^  UK | 10/2019 | 255 | WEMWBSf | 45.2 (9.39) | 42.3 (9.98) | -2.90 (NR) | 0.30 (0.12, 0.47) | ---------- | ---------- | ---------- | ---------- |
|  | 10/2020 |  |  |  |  |  |  |  |  |  |  |
| Truskauskaite-Kuneviciene^S66^  Lithuania Germany | 10-12/2019 | Lithuania: 450; Germany: 325 | PMH | Lithuania: 15.66 (5.57); Germany: 18.45 (5.68) | Lithuania: 16.44 (5.46); Germany: 18.64 (5.76) | Lithuania: 0.78 (NR); Germany: 0.19 (NR) | Lithuania: -0.14 (-0.27, -0.01); Germany: -0.03 (-0.19, 0.12) | ---------- | ---------- | ---------- | ---------- |
|  | 03-04/2020 |  |  |  |  |  |  |  |  |  |  |
| Wang, Yitao^S68^  China | 11/2019 | 2,559 | SCL-90-R | 139.64 (38.46) | 134.57 (40.44) | -5.07 (NR) | -0.13 (-0.18, -0.07) | ---------- | ---------- | ---------- | ---------- |
|  | 06/2020 |  |  |  |  |  |  |  |  |  |  |
| **Children and Adolescents** | | | | | | | | | | | |
| Achterberg^S72^  Netherlands | 01-11/2019 | 151 | SDQ-Internalizing Behaviors | 0.28 (0.35) | 0.29 (0.35) | 0.01 (NR) | 0.03 (-0.16, 0.22) | ---------- | ---------- | ---------- | ---------- |
|  | 04-05/2020 |  | SDQ-Externalizing Behaviors | 0.42 (0.39) | 0.39 (0.38) | -0.03 (NR) | -0.08 (-0.27, 0.11) | ---------- | ---------- | ---------- | ---------- |
| Bado^S74^  Brazil | NR/2018-2019 | 672 | SDQ - Total score | 13.93 (6.16) | 13.23 (6.43) | -0.70 (NR) | -0.11 (-0.22, 0.00) | ---------- | ---------- | ---------- | ---------- |
|  |  |  | SDQ - Emotion subscale | 4.21 (2.63) | 3.82 (2.74) | -0.39 (NR) | -0.15 (-0.25, -0.04) |  |  |  |  |
|  | 04/2020-04/2021 |  |  |  |  |  |  |  |  |  |  |
| Bernasco^S75^  Netherlands | NR/2019 | 245 | RCADS - Parents  RCADS - Adolescents | 1.17 (0.17) | 1.18 (0.19) | 0.01 (NR) | 0.06 (-0.12, 0.23) | ---------- | ---------- | ---------- | ---------- |
|  |  |  |  | 1.47 (0.31) | 1.43 (0.33) | -0.04 (NR) | -0.12 (-0.30, 0.05) |  |  |  |  |
|  | 04-07/2020 |  |  |  |  |  |  |  |  |  |  |
| Bosch^S76^  Spain | NR/2019 | 552 | SDQ - Emotion Symptoms | 2.42 (2.14) | 3.29 (2.29) | 0.87 (NR) | 0.39 (0.27, 0.51) | ---------- | ---------- | ---------- | ---------- |
|  |  |  | SDQ - Total | 8.62 (5.59) | 11.20 (5.62) | 2.58 (NR) | 0.46 (0.34, 0.58) |  |  |  |  |
|  | 05/2020-06/2020 |  |  |  |  |  |  |  |  |  |  |
| Chen, I-H^S78^  China | 10-11/2019 | 535 | DASS-21 | 0.46 (0.49) | 1.22 (0.30) | 0.76 (NR) | 1.87 (1.72, 2.01) | ---------- | ---------- | ---------- | ---------- |
|  | 03/2020 |  |  |  |  |  |  |  |  |  |  |
| Chen, C-Y^S79^  China | 10-11/2019 | 575 | DASS-21 | 21.85 (22.94) | 19.15 (22.13) | -2.70 (NR) | -0.12 (-0.24, 0.00) | ---------- | ---------- | ---------- | ---------- |
|  | 01/2020 |  |  |  |  |  |  |  |  |  |  |
| Daniunaite^S80^  Lithuania | 03-05/2019 | 331 | SDQ - Emotional Symptoms | 2.86 (2.29) | 3.27 (2.47) | 0.41 (NR) | 0.17 (0.02, 0.32) | ---------- | ---------- | ---------- | ---------- |
|  | 09-10/2020 |  |  |  |  |  |  |  |  |  |  |
| Ezpeleta^S81^  Spain | NR/2019 | 197 | SDQ-total Parent Version | 5.45 (4.65) | 6.20 (4.44) | 0.75 (3.75) | 0.16 (-0.03, 0.36) | ---------- | ---------- | ---------- | ---------- |
|  | 06/2020 |  |  |  |  |  |  |  |  |  |  |
| Fujihara^S82^  Japan | 12/2019 | 1,854 | K6 | 5.04 (5.07) | 5.73 (5.14) | 0.69 (NR) | 0.14 (0.07, 0.20) | ---------- | ---------- | ---------- | ---------- |
|  | 02/2020 |  |  |  |  |  |  |  |  |  |  |
| Hu^S83^  UK | NR | 886 | SDQ - Emotion Problems | 3.22 (2.44) | 3.45 (2.44) | 0.23 (NR) | 0.09 (0.00, 0.19) | ---------- | ---------- | ---------- | ---------- |
|  | 07/2020 |  |  |  |  |  |  |  |  |  |  |
| Knowles^S84^  UK | NR/2018-2019 | 958-1,055 | SDQ score | ---------- | ---------- | ---------- | ---------- | SDQ score ≥ 18 | 18.3 (13.9, 23.8) | 15.9 (13.0, 19.4) | -2.4 (-4.9, 0.1) |
|  | 05-08/2020 |  |  |  |  |  |  |  |  |  |  |
| Mastorci^S88^  Italy | 09-10/2019 | 1,019 | KIDSCREEN-52 (psychological wellbeing) | 50.23 (9.37) | 48.87 (9.83) | -1.36 (NR) | 0.14 (0.05, 0.23) | ---------- | ---------- | ---------- | ---------- |
|  |  |  | KIDSCREEN-52 (mood/emotion) | 48.62 (9.90) | 48.03 (9.82) | -0.59 (NR) | 0.06 (0.03, 0.15) |  |  |  |  |
|  | 04/2020 |  |  |  |  |  |  |  |  |  |  |
| Meireles^S89^  Portugal | 04-07/2019 | 1,099 | KIDSCREEN-10 | 3.72 (0.61) | 3.80 (0.56) | 0.08 (NR) | -0.14 (-0.22, -0.05) | ---------- | ---------- | ---------- | ---------- |
|  | 05-06/2020 |  |  |  |  |  |  |  |  |  |  |
| Paizan^S91^  Germany | 06-10/2019 | 226 | SWLS | 5.37 (1.19) | 5.11 (1.27) | -0.26 (NR) | 0.21 (0.03, 0.40) | ---------- | ---------- | ---------- | ---------- |
|  | 05-07/2020 |  |  |  |  |  |  |  |  |  |  |
| Rau^S93^  Germany | 10-11/2019 | 777 | KIDSCREEN-10 | 51.40 (13.10) | 52.30 (13.90) | 0.90 (NR) | -0.07 (-0.17, 0.03) | ---------- | ---------- | ---------- | ---------- |
|  | 06-07/2020 |  |  |  |  |  |  |  |  |  |  |
| Shoshani^S94^  Israel | 09/2019 | 1,537 | PANAS-C - PE | 18.15 (3.74) | 16.20 (3.88) | -1.95 (NR) | 0.51 (0.44, 0.58) | ---------- | ---------- | ---------- | ---------- |
|  |  |  | PANAS-C - NE | 9.58 (3.27) | 9.54 (3.29) | -0.04 (NR) | -0.01 (-0.08, 0.06) |  |  |  |  |
|  | 05/2020 |  | GSI-18 - BSI | 16.47 (11.26) | 19.18 (12.03) | 2.71 (NR) | 0.23 (0.16, 0.30) |  |  |  |  |
| Vira^S96^  Sweden | 10/2019-01/2020 | 849 | SDQ-emotional problems | 1.50 (0.45) | 1.53 (0.46) | 0.03 (NR) | 0.07 (-0.03, 0.16) | ---------- | ---------- | ---------- | ---------- |
|  | 11/2020-02/2021 |  |  |  |  |  |  |  |  |  |  |
| **Parents** | | | | | | | | | | | |
| Achterberg^S72^  Netherlands | 01-11/2019 | 106 | BSI | 0.19 (0.22) | 0.34 (0.32) | 0.15 (NR) | 0.54 (0.27, 0.82) | ---------- | ---------- | ---------- | ---------- |
|  | 04-05/2020 |  |  |  |  |  |  |  |  |  |  |
| Bosch^S76^  Spain | NR/2019 | 699 | SDQ - Emotion Symptoms | 1.63 (1.85) | 2.16 (2.03) | 0.53 (NR) | 0.27 (0.17, 0.38) | ---------- | ---------- | ---------- | ---------- |
|  |  |  | SDQ - Total | 7.02 (5.57) | 9.40 (5.61) | 2.38 (NR) | 0.43 (0.32, 0.53) |  |  |  |  |
|  | 05/2020-06/2020 |  |  |  |  |  |  |  |  |  |  |
| Gagné^S104^  Canada | 03-05/2019 | 127 | K10 | 1.75 (0.52) | 1.85 (0.61) | 0.10 (NR) | 0.18 (-0.07, 0.42) | K10 ≥ 9 | 41.7 (33.5, 50.4) | 40.2 (32.0, 48.9) | -1.6 (-12.3, 9.2) |
|  | 05-07/2020 |  |  |  |  |  |  |  |  |  |  |
| **People with Pre-existing Medical Conditions** | | | | | | | | | | | |
| Becker^S109^  USA | 03/2019 | 119 | SF-36 (role emotional) | 77.30 (26.30) | 73.70 (27.60) | -3.60 (NR) | -0.13 (-0.39, 0.12) | ---------- | ---------- | ---------- | ---------- |
|  | 03/2020 |  |  |  |  |  |  |  |  |  |  |
| Bonenkamp^S110^  Netherlands | 08/2019 | 177 | SF-12 Mental Component Summary | 48.08 (10.15) | 49.00 (10.04) | 0.91 (10.18) | 0.09 (-0.12, 0.30) | ---------- | ---------- | ---------- | ---------- |
|  | 07/2020 |  |  |  |  |  |  |  |  |  |  |
| Chiu^S112^  USA | 10/2018 | 133 |  |  |  |  |  | ---------- | ---------- | ---------- | ---------- |
|  |  |  | SPANE-P | 22.78 (3.88) | 21.11 (4.18) | -1.67 (5.31) | 0.41 (0.17, 0.66) |  |  |  |  |
|  | 09/2020 |  | SPANE-N | 14.50 (4.35) | 16.11 (4.51) | 1.61 (5.95) | 0.36 (0.12, 0.60) |  |  |  |  |
| Derksen^S113^  Netherlands | 01/2019-01/2020 | 2176 | EORTC QLQ-C30-Global quality of life | 79.83 (16.38) | 79.41 (16.18) | -0.42 (10.23) | 0.03 (-0.03, 0.09) | ---------- | ---------- | ---------- | ---------- |
|  | 04-06/2020 |  | EORTC QLQ-C30-Emotional functioning | 86.93 (17.37) | 87.92 (15.79) | 0.99 (14.95) | -0.06 (-0.12, -0.00) |  |  |  |  |
| Dunlop-Thomas^S114^  USA | NR/2017-2019 | 852 | PROMIS - Global mental health | 43.57 (9.34) | 43.75 (9.08) | 0.18 (NR) | 0.02 (-0.08, 0.11) | ---------- | ---------- | ---------- | ---------- |
|  | NR/2020-2021 |  |  |  |  |  |  |  |  |  |  |
| Fujiwara^S115^  Japan | 07-09/2019 | 245 | EQ-5D-5L | Median (IQR): 0.69 (0.27) | Median (IQR): 0.69 (0.30) | NR (NR) | NR (NR) | ---------- | ---------- | ---------- | ---------- |
|  | 07-09/2020 |  |  |  |  |  |  |  |  |  |  |
| García-Rudolph^S116^  Spain | NR | 175 | WHOQOL-BREF | 61.71 (19.75) | 57.95 (21.96) | -3.76 (NR) | 0.18 (-0.03, 0.39) | ---------- | ---------- | ---------- | ---------- |
|  | 11/2020 |  |  |  |  |  |  |  |  |  |  |
| Johnstone^S119^  New Zealand | NR/2018 | 104 | QOLS | 78.74 (14.18) | 73.17 (17.66) | -5.57 (NR) | 0.35 (0.07, 0.62) | ---------- | ---------- | ---------- | ---------- |
|  | 07-09/2020 |  |  |  |  |  |  |  |  |  |  |
| Lim^S122^  USA | NR/2018 | 316 | PROMIS Mental Health | 44.30 (9.30) | 44.50 (8.90) | 0.20 (7.40) | 0.02 (-0.13, 0.18) | ---------- | ---------- | ---------- | ---------- |
|  | 04/2020 |  |  |  |  |  |  |  |  |  |  |
| Möller^S123^  Australia, New Zealand | 08-10/2019 | 674 | DASS-21 | 18.95 (18.35) | 18.07 (17.53) | -0.88 (NR) | -0.05 (-0.16, 0.06) | ---------- | ---------- | ---------- | ---------- |
|  | 05-07/2020 |  | EUROHIS-QOL | 32.35 (5.34) | 32.70 (5.06) | 0.35 (NR) | -0.07 (-0.17, 0.04) |  |  |  |  |
| Park^S124^  Germany | 09/2019–02/2020 | 152 | EQ-5D-3L | Median (IQR): 8.00 (7.00, 9.00) | Median (IQR): 8.00 (6.00, 9.00) | NR (NR) | NR (NR) | ---------- | ---------- | ---------- | ---------- |
|  | 05-09/2020 |  |  |  |  |  |  |  |  |  |  |
| Sacre^S125^  Australia | NR/2018-2019 | 450 | PAID | ---------- | ---------- | ---------- | ---------- | PAID ≥ 40 | 14.7 (11.7, 18.2) | 7.8 (5.7, 10.6) | -6.9 (-9.6, 4.5) |
|  | 05-06/2020 |  |  |  |  |  |  |  |  |  |  |
| Sbragia^S126^  Italy | 01/2019 | 106 | HADS | 12.91 (6.52) | 11.60 (7.17) | -1.31 (NR) | -0.19 (-0.46, 0.08) | ---------- | ---------- | ---------- | ---------- |
|  | 05/2020 |  |  |  |  |  |  |  |  |  |  |
| Soltanzadeh^S14^  Iran | 11/2019 | 136 | GHQ-28 | 47.51 (11.37) | 54.61 (13.23) | 7.10 (NR) | 0.56 (0.09, 1.04) | ---------- | ---------- | ---------- | ---------- |
|  | 07/2020 |  |  |  |  |  |  |  |  |  |  |
| Thygesen^S15^  Denmark | 09-12/2019 | 1,543 | SWEMWBS | 24.20 (3.01) | 23.60 (2.00) | -0.60 (NR) | 0.23 (0.16, 0.31) | ---------- | ---------- | ---------- | ---------- |
|  | 09-11/2020 |  |  |  |  |  |  |  |  |  |  |
| **People with Pre-existing Mental Health Conditions** | | | | | | | | | | | |
| Huong^S130^  Taiwan | 01-12/2018 | 114 | BSRS-5 | 12.04 (6.19) | 10.58 (7.00) | -1.46 (NR) | -0.22 (-0.48, 0.04) | BSRS-5 ≥ 10 | 67.5 (58.5, 75.4) | 57.9 (48.7, 66.6) | -9.6 (-23.0, 4.1) |
|  | 01-05/2020 |  |  |  |  |  |  |  |  |  |  |
| Thygesen^S15^  Denmark | 09-12/2019 | 343 | SWEMWBS | 24.20 (3.01) | 23.60 (2.00) | -0.60 (NR) | -0.22 (-0.37, -0.07) | ---------- | ---------- | ---------- | ---------- |
|  | 09-11/2020 |  |  |  |  |  |  |  |  |  |  |
| **Women or Females** | | | | | | | | | | | |
| Dong^S45^  China | 09/2019 | 3,162-3,277 | ---------- | ---------- | ---------- | ---------- | ---------- | SCL-90-R ≥ 160 | 19.7 (18.4, 21.1) | 27.9 (26.4, 29.5) | 8.2 (6.3, 10.0) |
|  | NR/2020 |  |  |  |  |  |  |  |  |  |  |
| Fujihara^S82^  Japan | 12/2019 | 942 | K6 | 5.07 (NR) | 5.85 (NR) | 0.78 (NR) | NR (NR) | ---------- | ---------- | ---------- | ---------- |
|  | 02/2020 |  |  |  |  |  |  |  |  |  |  |
| Megias-Robles^S10^  Spain | 11/2019 | 67 | PANAS-NA | 1.93 (0.65) | 2.28 (0.79) | 0.34 (0.86) | 0.48 (0.13, 0.82) | ---------- | ---------- | ---------- | ---------- |
|  | 04/2020 |  |  |  |  |  |  |  |  |  |  |
| Meireles^S89^  Portugal | 04-07/2019 | 582 | KIDSCREEN-10 | 3.75 (0.61) | 3.73 (0.55) | -0.02 (NR) | 0.03 (-0.08, 0.15) | ---------- | ---------- | ---------- | ---------- |
|  | 05-06/2020 |  |  |  |  |  |  |  |  |  |  |
| Pierce^S11^  UK | Pre-COVID-19 waves | 7,181^30,b^ 6,380^34^ | GHQ-12 |  |  |  |  | GHQ-12 ≥ 4 |  |  |  |
| Daly^S12^  UK |  |  |  |  |  |  |  |  |  |  |  |
|  | 04/2020 |  |  | 12.00 (5.91) | 13.60 (7.14) | 1.60 (NR)^c^ | 0.24 (0.21, 0.28) |  | 24.5 (22.5, 26.4)^e^ | 36.8 (34.8, 38.9)^e^ | 12.4 (9.9, 14.9)^e^ |
|  |  |  |  |  |  | 0.88 (NR)^d^ | 0.13 (0.10, 0.17) |  |  |  |  |
|  | 09/2020 |  |  | ---------- | ---------- | ---------- | ---------- |  | 24.5 (22.5, 26.4)^e^ | 25.0 (23.3, 26.8)^e^ | 0.5 (-1.8, 2.9)^e^ |
| Savage, 2020^S63^  UK | 10/2019 | 154 | WEMWBS^f^ | 43.00 (9.00) | 40.00 (10.00) | -3.00 (NR) | 0.31 (0.09, 0.54) | ---------- | ---------- | ---------- | ---------- |
|  | 04/2020 |  |  |  |  |  |  |  |  |  |  |
| Soltanzadeh^S14^  Iran | 11/2019 | 161 | GHQ-28 | 45.11 (12.07) | 50.91 (12.69) | 5.80 (NR) | 0.47 (0.24, 0.69) | ---------- | ---------- | ---------- | ---------- |
|  | 07/2020 |  |  |  |  |  |  |  |  |  |  |
| Thygesen^S15^  Denmark | 09-12/2019 | 2,184 | SWEMWBS | 25.10 (9.54) | 24.10 (4.77) | -1.00 (NR) | 0.13 (0.07, 0.19) | ---------- | ---------- | ---------- | ---------- |
|  | 09-11/2020 |  |  |  |  |  |  |  |  |  |  |
| van der Velden, 2020^S16^  Netherlands | 03/2019 | 2,020 | MHI-5^f^ | ---------- | ---------- | ---------- | ---------- | MHI-5 ≤ 59 |  |  |  |
| van der Velden, 2021^S17^  Netherlands | 11-12/2019 | 2,062 |  |  |  |  |  |  |  |  |  |
|  | 03/2020 |  |  |  |  |  |  |  | 18.9 (17.3, 20.7) | 18.3 (16.7, 20.1) | -0.6 (-2.5, 1.3) |
|  | 11-12/2020 |  |  |  |  |  |  |  | 19.1 (17.4, 20.8) | 17.8 (16.2, 19.5) | -1.3 (-3.1, 0.6) |
| **Men or Males** | | | | | | | | | | | |
| Dong^S45^  China | 09/2019 | 923-1,064 | ---------- | ---------- | ---------- | ---------- | ---------- | SCL-90-R ≥ 160 | 14.3 (12.3, 16.5) | 21.2 (18.7, 24.0) | 6.9 (4.0, 9.9) |
|  | NR/2020 |  |  |  |  |  |  |  |  |  |  |
| Fujihara^S82^  Japan | 12/2019 | 912 | K6 | 5.01 (NR) | 5.61 (NR) | 0.60 (NR) | NR (NR) | ---------- | ---------- | ---------- | ---------- |
|  | 02/2020 |  |  |  |  |  |  |  |  |  |  |
| Megias-Robles^S10^  Spain | 11/2019 | 35 | PANAS-NA | 1.88 (0.67) | 2.11 (0.66) | 0.23 (0.66) | 0.34 (-0.14, 0.82) | ---------- | ---------- | ---------- | ---------- |
|  | 04/2020 |  |  |  |  |  |  |  |  |  |  |
| Meireles^S89^  Portugal | 04-07/2019 | 517 | KIDSCREEN-10 | 3.69 (0.61) | 3.88 (0.56) | 0.19 (NR) | -0.32 (-0.45, -0.20) | ---------- | ---------- | ---------- | ---------- |
|  | 05-06/2020 |  |  |  |  |  |  |  |  |  |  |
| Pierce^S11^  UK | Pre-COVID-19 waves | 8,195^30,b^ 4,538^34^ | GHQ-12 |  |  |  |  | GHQ-12 ≥ 4 |  |  |  |
| Daly^S12^  UK |  |  |  |  |  |  |  |  |  |  |  |
|  | 04/2020 |  |  | 10.80 (4.99) | 11.50 (5.75) | 0.70 (NR)^c^ | 0.13 (0.10, 0.16) |  | 16.7 (14.6, 18.7)^e^ | 21.1 (19.0, 23.3)^e^ | 4.5 (2.0, 7.0)^e^ |
|  |  |  |  |  |  | 0.03 (NR)^d^ | 0.01 (-0.03, 0.04) |  |  |  |  |
|  | 09/2020 |  |  | ---------- | ---------- | ---------- | ---------- |  | 16.7 (14.6, 18.7)^e^ | 16.0 (14.0, 17.9)^e^ | -0.7 (-2.9, 1.5)^e^ |
| Savage, 2020^S63^  UK | 10/2019 | 60 | WEMWBS^f^ | 47.00 (9.00) | 44.00 (10.00) | -3.00 (NR) | 0.31 (-0.04, 0.67) | ---------- | ---------- | ---------- | ---------- |
|  | 04/2020 |  |  |  |  |  |  |  |  |  |  |
| Soltanzadeh^S14^  Iran | 11/2019 | 689 | GHQ-28 | 46.12 (11.36) | 51.38 (12.34) | 5.26 (NR) | 0.44 (0.34, 0.55) | ---------- | ---------- | ---------- | ---------- |
|  | 07/2020 |  |  |  |  |  |  |  |  |  |  |
| Thygesen^S15^  Denmark | 09-12/2019 | 2,050 | SWEMWBS | 25.10 (5.78) | 24.40 (16.17) | -0.70 (NR) | 0.06 (-0.00, 0.12) | ---------- | ---------- | ---------- | ---------- |
|  | 09-11/2020 |  |  |  |  |  |  |  |  |  |  |
| van der Velden, 2020^S16^  Netherlands | 03/2019 | 1,962-1,963 | MHI-5^f^ | ---------- | ---------- | ---------- | ---------- | MHI-5 ≤ 59 |  |  |  |
| van der Velden, 2021^S17^  Netherlands | 11-12/2019 | 2,002 |  |  |  |  |  |  |  |  |  |
|  | 03/2020 |  |  |  |  |  |  |  | 14.6 (13.1, 16.3) | 15.6 (14.1, 17.3) | 1.0 (-0.8, 2.7) |
|  | 11-12/2020 |  |  |  |  |  |  |  | 14.7 (13.2, 16.3) | 15.9 (14.4, 17.6) | 1.2 (-0.5, 3.0) |

BSI = Brief Symptom Inventory; DToS = Distress Tolerance Scale; GHQ-12 = General Health Questionnaire-12; MHI-5 = Mental Health Index-5; PANAS – NA = Positive and Negative Affect Schedule – Negative Affect; PANAS – PA = Positive and Negative Affect Schedule – Positive Affect; PHQ-4 = Patient Health Questionnaire-4; RRQ = Reflection and Rumination Scale; SCL-90-R = Symptom Check List-90-Revised; SDQ – Extern = Strengths and Difficulties Questionnaire – Externalizing Behavior; SDQ – Intern = Strengths and Difficulties Questionnaire – Internalizing Behavior; SDQ – Total = Strengths and Difficulties Questionnaire – Total; SWLS = Satisfaction with Life Scale; WEMWBS = Warwick Edinburgh Mental Wellbeing Scale.

^a^Positive Hedges’ g effect sizes and increases in proportions above a threshold indicate worse mental health in COVID-19 compared to pre-COVID-19. Effects for measures where high scores = positive outcomes were reversed to reflect this. ^b^Number included in fixed effects regression analysis from where majority of data were extracted. ^c^Based on difference between 2020 and 2019 outcomes. ^d^Based on estimate from fixed effects regression model that estimates within-person change accounting for pre-COVID-19 trends. ^e^Included proportion outcomes from Daly,^31^ since Daly reported for two time points. ^f^Higher scale scores reflect better mental health; thus, direction of effect sizes reversed. ^g^Proportions in the study were calculated using age categories based on previous year’s age.

**Supplementary Table 4.** **Individual Study Results for Anxiety Symptoms**

| **First Author** | **Pre- and Post-COVID-19 Data Collection** | **N** | **Continuous**  **Outcome Measure** | **Pre-**  **COVID-19 Mean (SD)** | **Post-**  **COVID-19 Mean (SD)** | **Mean (SD) Change**^a^ | **Hedges’ g Standardized Mean Difference (95% CI)** | **Dichotomous Outcome Measure** | **% pre-COVID-19 (95% CI)** | **% post-COVID-19 (95% CI)** | **% Change with 95% CI**^a^ |
| --- | --- | --- | --- | --- | --- | --- | --- | --- | --- | --- | --- |
| **General Population** | | | | | | | | | | | |
| Chan^S3^  Hong Kong, China | 07/2019 | 279 | HAI | 15.33 (6.31) | 15.52 (6.70) | 0.19 (NR) | 0.03 (-0.14, 0.20) | ---------- | ---------- | ---------- | ---------- |
|  | 07/2020 |  |  |  |  |  |  |  |  |  |  |
| Ge^S5^  China | 01-12/2019 | 1,547-1,978 | GAD-7 | 9.24 (2.33) | 10.02 (2.28) | 0.78 (NR) | 0.1 (0.03, 0.18) | ---------- | ---------- | ---------- | ---------- |
|  | 02-03/2020 |  |  |  |  |  |  |  |  |  |  |
| Haliwa^S6^  USA | 09-12/2019 | Sample 1: 300; Sample 2: 146; Sample 3: 142 | Sample 1: GAD-7 Sample 2: DASS-21-Anxiety Sample 3: GAD-7 | Sample 1: 5.58 (5.02) Sample 2: 3.53 (4.89) Sample 3: 4.64 (5.35) | Sample 1: 6.55 (5.98) Sample 2: 3.25 (4.51) Sample 3: 4.82 (5.60) | Sample 1: 0.97 (4.93) Sample 2: -0.28 (2.82) Sample 3: 0.18 (4.21) | Sample 1: 0.18 (0.01, 0.34) Sample 2: -0.08 (-0.31, 0.15) Sample 3: 0.03 (-0.20, 0.27) | Sample 1: GAD-7 ≥ 10 Sample 2: DASS-21-Anxiety ≥ 6 Sample 3: GAD-7 ≥ 10 | Sample 1: 19.0 (15.0, 23.8) Sample 2: 21.9 (16.0, 29.3) Sample 3: 20.4 (14.6, 27.8) | Sample 1: 29.7 (24.8, 35.1) Sample 2: 23.3 (17.2, 30.8) Sample 3: 19.7 (14.0, 27.0) | Sample 1: 10.7 (4.6, 16.6) Sample 2: 1.4 (-6.5, 9.2) Sample 3: -0.7 (-8.0, 6.5) |
|  | 04-06/2020 |  |  |  |  |  |  |  |  |  |  |
| Kanbur^S7^  Turkey | NR/2019 | 400 | SCL-90-R Anxiety | 0.27 (NR) | 0.51 (NR) | 0.24 (NR) | NR (NR) | ---------- | ---------- | ---------- | ---------- |
|  | NR/2020 |  |  |  |  |  |  |  |  |  |  |
| Katz, B^S8^  Canada, Ireland, UK, USA | 04/2019 | 218 | DASS-21 Anxiety | 3.25 (3.91) | 2.83 (3.61) | -0.42 (3.13) | -0.11 (-0.30, 0.08) | ---------- | ---------- | ---------- | ---------- |
|  | 04/2020 |  |  |  |  |  |  |  |  |  |  |
| **Older Adults** | | | | | | | | | | | |
| Bartlett^S19^  Australia | 10/2019 | 1,671 | HADS-A | 5.56 (3.55) | 4.88 (3.34) | -0.68 (NR) | -0.20 (-0.27, -0.13) | ---------- | ---------- | ---------- | ---------- |
|  | 04-06/2020 |  |  |  |  |  |  |  |  |  |  |
| Creese^S21^  UK | 10/2019 | 3,281 | GAD-7 | 1.55 (2.64) | 1.94 (2.84) | 0.39 (NR) | 0.14 (0.09, 0.19) | GAD-7 ≥ 10 | 2.2 (1.8, 2.8) | 2.7 (2.2, 3.3) | 0.5 (-0.1, 1.1) |
|  | 05-06/2020 |  |  |  |  |  |  |  |  |  |  |
| Herrera^S23^  Chile | 11/2019 | 721 | GAI-SF | 2.04 (NR) | 2.26 (NR) | 0.22 (NR) | NR (NR) | GAI-SF ≥ 3 | 40.0 (36.5, 43.6) | 42.9 (39.3, 46.5) | 2.9 (-1.9, 7.7) |
|  | 09/2020 |  |  |  |  |  |  |  |  |  |  |
| Rentscher^S29^  USA | 02-06/2019 | 165 | STAI-State | 27.50 (7.10) | 30.60 (9.60) | 3.10 (NR) | 0.37 (0.15, 0.58) | ---------- | ---------- | ---------- | ---------- |
|  | 05-09/2020 |  |  |  |  |  |  |  |  |  |  |
| Rentscher^S29^  USA | 02-06/2019 | 262 | STAI-State | 27.90 (6.50) | 30.10 (9.10) | 2.20 (NR) | 0.28 (0.11, 0.45) | ---------- | ---------- | ---------- | ---------- |
|  | 05-09/2020 |  |  |  |  |  |  |  |  |  |  |
| Siew^S31^  Singapore | 02/2018-01/2020 | 411 | GAI-SF | 1.12 (2.63) | 1.40 (3.17) | 0.28 (NR) | 0.10 (-0.04, 0.23) | ---------- | ---------- | ---------- | ---------- |
|  | 05-06/2020 |  |  |  |  |  |  |  |  |  |  |
| van den Besselaar^S32^  Netherlands | NR/2018-2019 | 984 | HADS-A | 2.58 (2.70) | 3.35 (2.99) | 0.77 (NR) | 0.27 (0.18, 0.36) | ---------- | ---------- | ---------- | ---------- |
|  | 06-10/2020 |  |  |  |  |  |  |  |  |  |  |
| Wong, S^S35^  Hong Kong, China | 04/2018-03/2019 | 583 | GAD-7 | 2.50 (NR) | 3.00 (NR) | 0.48 (NR) | NR^b^ | ---------- | ---------- | ---------- | ---------- |
|  | 03-04/2020 |  |  |  |  |  |  |  |  |  |  |
| Yu^S36^  Singapore | 02/2018-01/2020 | 419 | GAI | 1.12 (2.58) | 1.38 (3.14) | 0.26 (2.31) | 0.09 (-0.05, 0.23) | ---------- | ---------- | ---------- | ---------- |
|  | 05-06/2020 |  |  |  |  |  |  |  |  |  |  |
| **Young Adults** | | | | | | | | | | | |
| Rimfeld^S39^  UK | NR/2018 | 3,563-3,694 | SMGAD | 7.48 (7.35) | 8.69 (7.54) | 1.21 (6.83) | 0.16 (0.12, 0.21) | ---------- | ---------- | ---------- | ---------- |
|  | 04-05/2020 |  |  |  |  |  |  |  |  |  |  |
| Watkins-Martin^S43^  Canada | NR/2018 | 1,039 | SMGAD | 4.73 (4.61) | 4.45 (4.70) | -0.28 (NR) | -0.06 (-0.15, 0.03) | GAD-7 ≥ 15 | 4.9 (3.8, 6.4) | 4.7 (3.6, 6.2) | -0.2 (-1.5, 1.1) |
|  | 08/2020 |  |  |  |  |  |  |  |  |  |  |
| **University Students** | | | | | | | | | | | |
| Conceição^S44^  Portugal | 10/2019 | 341 | GAD-7 | 9.89 (6.19) | 12.15 (6.50) | 2.26 (NR) | 0.36 (0.20, 0.51) | GAD-7 ≥ 10 | 46.0 (40.8, 51.4) | 64.5 (59.3, 69.4) | 18.5 (10.1, 26.5) |
|  | 06/2020 |  |  |  |  |  |  |  |  |  |  |
| Elmer^S46^  Switzerland | 09/2019 | 209 | GAD-7 | NR | NR | 0.60 (3.47) | 0.17 (-0.02, 0.36) | ---------- | ---------- | ---------- | ---------- |
|  | 04/2020 |  |  |  |  |  |  |  |  |  |  |
| Evans^S47^  UK | 10/2019 | 251 | HADS-A | 9.35 (4.28) | 9.42 (4.47) | 0.07 (NR) | 0.02 (-0.16, 0.19) | ---------- | ---------- | ---------- | ---------- |
|  | 05/2020 |  |  |  |  |  |  |  |  |  |  |
| Gelezelyte^S49^  Lithuania | 10-12/2019 | 474 | DASS-21 Anxiety | 6.99 (4.94) | 5.87 (4.58) | -1.12 (4.32) | -0.23 (-0.36, -0.11) | ---------- | ---------- | ---------- | ---------- |
|  | 10-12/2020 |  |  |  |  |  |  |  |  |  |  |
| Gopalan^S50^  USA | 11/2019 | 1,004 | CCAPS-62 - Anxiety | 1.31 (1.04) | 1.34 (1.07) | 0.03 (NR) | 0.03 (-0.06, 0.12) | ---------- | ---------- | ---------- | ---------- |
|  | 05/2020 |  |  |  |  |  |  |  |  |  |  |
| Hamza^S51^  Canada | 05/2019 | 733 | GAD-7 | 6.68 (5.53) | 6.39 (5.46) | -0.29 (NR) | -0.05 (-0.16, 0.05) | ---------- | ---------- | ---------- | ---------- |
|  | 05/2020 |  |  |  |  |  |  |  |  |  |  |
| He^S52^  China | 09-12/2019 | 589 | STAI-Trait | 43.27 (7.35) | 44.76 (8.78) | 1.49 (NR) | 0.18 (0.07, 0.30) | ---------- | ---------- | ---------- | ---------- |
|  | 02/2020 |  |  |  |  |  |  |  |  |  |  |
| Koelen^S53^  Netherlands | 01/2019-01/2020 | 683 | GAD-7 | ---------- | ---------- | ---------- | ---------- | GAD-7 ≥ 10 | 33.6 (30.1, 37.2) | 36.5 (32.9, 40.1) | 2.9 (-1.6, 7.4) |
|  | 04-05/2020 |  |  |  |  |  |  |  |  |  |  |
| Li, Wendy Wen^S56^  China | 11/2019 | 173 | DASS-21 Anxiety | 9.23 (6.16) | 5.09 (5.90) | -4.14 (NR) | -0.68 (-0.90, -0.47) | ---------- | ---------- | ---------- | ---------- |
|  | 03/2020 |  |  |  |  |  |  |  |  |  |  |
| Lu^S58^  China | 09/2019-10/2019 | 5,181 | GAD-7 | ---------- | ---------- | ---------- | ---------- | GAD-7 ≥ 10 | 3.5 (3.0, 4.0) | 3.7 (3.2, 4.2) | 0.2 (-0.3, 0.7) |
|  | 04/2020 |  |  |  |  |  |  |  |  |  |  |
| Mauer^S59^  USA | 09-12/2019 | 1,434 | DASS-21 | 5.42 (4.68) | 5.04 (4.30) | -0.38 (NR) | -0.08 (-0.16, -0.01) | DASS-21 ≥ 10 | 16.6 (14.8, 18.6) | 16.0 (14.2, 18.0) | -0.6 (-2.7, 1.4) |
|  | 03-06/2020 |  |  |  |  |  |  |  |  |  |  |
| Mehus^S60^  USA | 08,12/2019 | 727 | GAD-7 | 5.07 (4.68) | 5.67 (5.09) | 0.60 (NR) | 0.12 (0.02, 0.23) | GAD-7 ≥ 8 | 24.3 (21.4, 27.6) | 29.6 (26.3, 33.0) | 5.3 (1.3, 9.1) |
|  | 04/2020 |  |  |  |  |  |  |  |  |  |  |
| Saraswathi^S62^  India | 12/2019 | 217 | DASS-21 Anxiety | 4.60 (6.19) | 6.11 (7.13) | 1.51 (NR) | 0.23 (0.04, 0.41) | DASS-21 Anxiety > 7 | 21.2 (16.3, 27.1) | 33.2 (27.3, 39.7) | 12.0 (4.4, 19.4) |
|  | 06/2020 |  |  |  |  |  |  |  |  |  |  |
| Truskauskaite-Kuneviciene^S66^  Lithuania Germany | 10-12/2019 | Lithuania: 450; Germany: 325 | DASS-21 - Anxiety | Lithuania: 7.07 (4.92); Germany: 3.66 (3.66) | Lithuania: 4.16 (4.21); Germany: 2.33 (2.82) | Lithuania: -2.91 (NR); Germany: -1.33 (NR) | Lithuania: -0.63 (-0.77, -0.50); Germany: -0.41 (-0.56, -0.25) | ---------- | ---------- | ---------- | ---------- |
|  | 03-04/2020 |  |  |  |  |  |  |  |  |  |  |
| Voltmer^S67^  Germany | NR/2019 | 587 | BSI-18 Anxiety | 4.50 (4.40) | 4.10 (4.10) | -0.40 (NR) | -0.09 (-0.21, 0.02) | ---------- | ---------- | ---------- | ---------- |
|  | 06/2020 |  |  |  |  |  |  |  |  |  |  |
| Wang, Yitao^S68^  China | 11/2019 | 2,559 | SCL-90-R Anxiety | 1.55 (0.49) | 1.48 (0.50) | -0.07 (NR) | -0.14 (-0.20, -0.09) | ---------- | ---------- | ---------- | ---------- |
|  | 06/2020 |  |  |  |  |  |  |  |  |  |  |
| Yang, Ziyan^S70^  China | 10/2019 | 2,364 | DASS-21 Anxiety | 9.64 (2.88) | 8.92 (2.96) | -0.72 (NR) | -0.25 (-0.30, -0.19) | ---------- | ---------- | ---------- | ---------- |
|  | 05/2020 |  |  |  |  |  |  |  |  |  |  |
| Zimmerman^S71^  USA | 08/2019 | 205 | GAD-7 | 8.29 (6.28) | 9.71 (6.83) | 1.42 (0.41) | 0.22 (0.02, 0.41) | ---------- | ---------- | ---------- | ---------- |
|  | 04/2020 |  |  |  |  |  |  |  |  |  |  |
| **Children and Adolescents** | | | | | | | | | | | |
| Chen, C-Y^S79^  China | 10-11/2019 | 575 | DASS-21 - Anxiety | 7.98 (7.93) | 7.01 (7.43) | -0.97 (NR) | -0.13 (-0.24, -0.01) | ---------- | ---------- | ---------- | ---------- |
|  | 01/2020 |  |  |  |  |  |  |  |  |  |  |
| Knowles^S84^  UK | NR/2018-2019 | 958-1,055 | GAD-7 | ---------- | ---------- | ---------- | ---------- | GAD-7 ≥ 10 | 20.5 (17.3, 24.3) | 17.3 (14.0, 21.0) | -3.1 (-5.8, -0.5) |
|  | 05-08/2020 |  |  |  |  |  |  |  |  |  |  |
| Li, Y^S85^  China | 09/2019 | 831 | ZSAS | ---------- | ---------- | ---------- | ---------- | ZSAS > 50 | 27.7 (24.7, 30.8) | 23.0 (20.3, 26.0) | -4.7 (-7.9, -1.4) |
|  | 03/2020 |  |  |  |  |  |  |  |  |  |  |
| Magson^S87^  Australia | NR/2019 | 248 | SCAS Generalized Anxiety | 4.60 (3.74) | 5.10 (4.05) | 0.50 (1.50) | 0.13 (-0.05, 0.30) | ---------- | ---------- | ---------- | ---------- |
|  | 05/2020 |  |  |  |  |  |  |  |  |  |  |
| Rau^S93^  Germany | 10-11/2019 | 777 | RCADS - Anxiety | 24.40 (17.70) | 21.10 (17.00) | -3.30 (NR) | -0.19 (-0.29, -0.09) | ---------- | ---------- | ---------- | ---------- |
|  | 06-07/2020 |  |  |  |  |  |  |  |  |  |  |
| Shoshani^S94^  Israel | 09/2019 | 1,537 | BSI-18 - Anxiety | 3.93 (2.68) | 5.24 (3.14) | 1.31 (NR) | 0.45 (0.38, 0.52) | ---------- | ---------- | ---------- | ---------- |
|  | 05/2020 |  |  |  |  |  |  |  |  |  |  |
| Teng^S95^  China | 10-11/2019 | 1,778 | STAI-Trait | 1.95 (0.65) | 1.98 (0.66) | 0.03 (NR) | 0.05 (-0.02, 0.11) | ---------- | ---------- | ---------- | ---------- |
|  | 04-05/2020 |  |  |  |  |  |  |  |  |  |  |
| Wang, Wanxin^S97^  China | 10-12/2019 | 1,790 | GAD-7 | 3.60 (4.32) | 3.56 (4.22) | -0.04 (NR) | -0.01 (-0.07,0.06) | GAD-7 ≥ 5 | 31.6 (29.5, 33.8) | 32.9 (30.7, 35.1) | 1.3 (-1.4, 3.9) |
|  | 10-12/2020 |  |  |  |  |  |  |  |  |  |  |
| Widnall^S98^  UK | 10/2019 | 603 | HADS-A | Median (IQR): 7.00 (4.00-11.00) | Median (IQR): 6.00 (3.00-10.00) | NR (NR) | NR (NR) | ---------- | ---------- | ---------- | ---------- |
|  | 05/2020 |  |  |  |  |  |  |  |  |  |  |
| Wong, R^S99^  China | 04-08/2019 | 233 | DASS-21 - Anxiety | NR (NR) | NR (NR) | 0.13 (5.42) | NR (NR) | ---------- | ---------- | ---------- | ---------- |
|  | 02/2020 |  |  |  |  |  |  |  |  |  |  |
| Zhang^S101^  China | 11/2019 | 1,241 | HBQ Anxiety | 3.06 (0.90) | 3.02 (1.05) | -0.05 (0.90) | -0.05 (-0.13, 0.03) | ---------- | ---------- | ---------- | ---------- |
|  | 05/2020 |  |  |  |  |  |  |  |  |  |  |
| **Parents** | | | | | | | | | | | |
| Loret de Mola^S105^  Brazil | 01-12/2019 | 1,028 | GAD-7 | ---------- | ---------- | ---------- | ---------- | GAD-7 ≥ 10 | 9.7 (8.1, 11.7) | 25.9 (23.3, 28.6) | 16.2 (13.2, 19.1) |
|  | 05-07/2020 |  |  |  |  |  |  |  |  |  |  |
| Thompson^S108^  USA | NR/2018-2019 | 147 | GAD-7 | 6.05 (4.70) | 7.42 (5.92) | 1.37 (6.02) | 0.25 (0.02, 0.49) | ---------- | ---------- | ---------- | ---------- |
|  | 04/2020 |  |  |  |  |  |  |  |  |  |  |
| **People with Pre-existing Medical Conditions** | | | | | | | | | | | |
| Chiu^S112^  USA | 10/2018 | 133 | HADS-A | 6.89 (3.92) | 6.95 (3.85) | 0.06 (3.30) | 0.02 (-0.22, 0.25) | ---------- | ---------- | ---------- | ---------- |
|  | 09/2020 |  |  |  |  |  |  |  |  |  |  |
| Derksen^S113^  Netherlands | 01/2019-01/2020 | 2176 | HADS-A | 3.24 (3.20) | 3.18 (3.16) | -0.06 (2.34) | -0.02 (-0.08, 0.04) | ---------- | ---------- | ---------- | ---------- |
|  | 04-06/2020 |  |  |  |  |  |  |  |  |  |  |
| Fujiwara^S115^  Japan | 07-09/2019 | 245 | HADS-A | Median (IQR): 6.00 (5.00) | Median (IQR): 6.00 (6.00) | NR (NR) | NR (NR) | ---------- | ---------- | ---------- | ---------- |
|  | 07-09/2020 |  |  |  |  |  |  |  |  |  |  |
| García-Rudolph^S116^  Spain | NR | 175 | HADS-A | 6.21 (4.28) | 6.52 (4.64) | 0.31 (NR) | 0.07 (-0.14, 0.28) | ---------- | ---------- | ---------- | ---------- |
|  | 11/2020 |  |  |  |  |  |  |  |  |  |  |
| Henry^S118^  Canada, France, UK, USA | 07-12/2019 | 435 | PROMIS Anxiety |  |  |  |  | ---------- | ---------- | ---------- | ---------- |
|  | 04/2020 |  |  | 52.66 (10.41) | 57.54 (8.79) | 4.88 (NR) | 0.51 (0.37, 0.64) |  |  |  |  |
|  | 09-10/2020 |  |  | 52.66 (10.41) | 53.75 (9.46) | 1.09 (NR) | 0.11 (-0.02, 0.24) |  |  |  |  |
|  | 03/2021 |  |  | 52.66 (10.41) | 53.19 (9.70) | 0.53 (NR) | 0.05 (-0.08, 0.18) |  |  |  |  |
| Johnstone^S119^  New Zealand | NR/2018 | 104 | HADS-A | 5.88 (4.12) | 5.55 (4.23) | -0.33 (NR) | -0.08 (-0.35, 0.19) | ---------- | ---------- | ---------- | ---------- |
|  | 07-09/2020 |  |  |  |  |  |  |  |  |  |  |
| Katz, P^S120^  USA | NR/2019 | 1,504 | GAD-2 | 0.66 (1.18) | 0.99 (1.35) | 0.33 (NR) | 0.26 (0.19, 0.33) | ---------- | ---------- | ---------- | ---------- |
|  | 03-06/2020 |  |  |  |  |  |  |  |  |  |  |
| Liang^S121^  China | 12/2019 | 114 | ZSAS | 32.80 (7.20) | 32.80 (7.20) | 0.00 (NR) | 0.00 (-0.27, 0.27) | ---------- | ---------- | ---------- | ---------- |
|  | 02-03/2020 |  |  |  |  |  |  |  |  |  |  |
| Lim^S122^  USA | NR/2018 | 316 | PROMIS Anxiety | 50.30 (11.30) | 50.30 (11.10) | 0.00 (10.20) | 0.00 (-0.16, 0.16) | ---------- | ---------- | ---------- | ---------- |
|  | 04/2020 |  |  |  |  |  |  |  |  |  |  |
| Park^S124^  Germany | 09/2019–02/2020 | 152 | HADS-A | Median (IQR): 6.00 (2.00, 9.00) | Median (IQR): 6.00 (3.00, 8.00) | NR (NR) | NR (NR) | HADS-A ≥ 8 | 17.0 (12.0, 23.9) | 11.0 (6.6, 16.4) | -5.9 (-11.5, -0.6) |
|  | 05-09/2020 |  |  |  |  |  |  |  |  |  |  |
| Rentscher^S29^  USA | 02-06/2019 | 262 | STAI-State | 27.90 (6.50) | 30.10 (9.10) | 2.20 (NR) | 0.28 (0.11, 0.45) | ---------- | ---------- | ---------- | ---------- |
|  | 05-09/2020 |  |  |  |  |  |  |  |  |  |  |
| Sacre^S125^  Australia | NR/2018-2019 | 450 | GAD-7 | 3.30 (4.10) | 3.10 (4.30) | -0.20 (NR) | -0.05 (-0.18, 0.08) | GAD-7 ≥ 10 | 8.4 (6.2, 11.4) | 8.4 (6.2, 11.4) | 0.0 (-2.8, 2.8) |
|  | 05-06/2020 |  |  |  |  |  |  |  |  |  |  |
| Sbragia^S126^  Italy | 01/2019 | 106 | HADS-A | 7.02 (3.62) | 6.09 (4.05) | -0.93 (NR) | -0.24 (-0.51, 0.03) | HADS-A > 8 | 54.0 (44.3, 63.0) | 46.0 (37.0, 55.7) | -8.0 (-19.9, 5.1) |
|  | 05/2020 |  |  |  |  |  |  |  |  |  |  |
| Wong, S^S35^  Hong Kong, China | 04/2018-03/2019 | 583 | GAD-7 | 2.50 (NR) | 3.00 (NR) | 0.48 (NR) | NR^b^ | ---------- | ---------- | ---------- | ---------- |
|  | 03-04/2020 |  |  |  |  |  |  |  |  |  |  |
| **People with Pre-existing Mental Health Conditions** | | | | | | | | | | | |
| Gentile^S129^  Italy, Paraguay | 10-12/2019  03-04/2020 | 110 | HAM-A | 16.60 (9.47) | 18.50 (9.68) | 1.90 (NR) | 0.20 (-0.07, 0.46) | ---------- | ---------- | ---------- | ---------- |
| Swerdlow^S131^  USA | 03/2017-04/2020 | 144 | MASQ-30 - Anxiety | 16.01 (5.29) | 17.89 (6.80) | 1.88 (NR) | 0.31 (0.08, 0.54) | ---------- | ---------- | ---------- | ---------- |
|  | 04-06/2020 |  |  |  |  |  |  |  |  |  |  |
| Young^S132^  UK | 09/2018-02/2020  04-09/2020 | 12108 | GAD-7 | 8.78 (5.96) | 8.48 (5.83) | -0.30 (NR) | -0.05 (-0.08, -0.03) | ---------- | ---------- | ---------- | ---------- |
| **Medical Staff** | | | | | | | | | | | |
| Li, Weidong^S133^  China | 10-11/2019 | 385 | GAD-7 | 4.33 (NR) | 5.43 (NR) | 1.10 (NR) | NR^b^ | ---------- | ---------- | ---------- | ---------- |
|  | 01-02/2020 |  |  |  |  |  |  |  |  |  |  |
| **Sexual or Gender Minority Individuals** | | | | | | | | | | | |
| Bavinton^S134^  Australia | NR/2019 | 681 | GAD-7 | 4.54 (4.95) | 4.96 (5.07) | 0.42 (NR) | 0.08 (-0.02, 0.19) | ---------- | ---------- | ---------- | ---------- |
|  | 04/2020 |  |  |  |  |  |  |  |  |  |  |
| Flentje^S135^  USA | 06/2019 | 2,282 | GAD-7 | 5.78 (5.21) | 8.89 (6.22) | 3.11 (5.32) | 0.54 (0.48, 0.60) | ---------- | ---------- | ---------- | ---------- |
|  | 03-04/2020 |  |  |  |  |  |  |  |  |  |  |
| Ghabrial^S136^  Canada | NR/2019 | 780 | OASIS | 10.13 (4.70) | 10.35 (4.42) | 0.22 (NR) | 0.05 (-0.05, 0.15) | ---------- | ---------- | ---------- | ---------- |
|  | 09-10/2020 |  |  |  |  |  |  |  |  |  |  |
| **Women or Females** | | | | | | | | | | | |
| Li, Y^S85^  China | 09/2019 | 328 | ZSAS | EMM (SE): 47.50 (0.50) | EMM (SE): 45.70 (0.52) | NR (NR) | NR (NR) | ---------- | ---------- | ---------- | ---------- |
|  | 03/2020 |  |  |  |  |  |  |  |  |  |  |
| Lim^S122^  USA | NR/2018 | 295 | PROMIS Anxiety | 50.20 (11.50) | 50.40 (11.10) | 0.20 (10.10) | 0.02 (-0.14, 0.18) | ---------- | ---------- | ---------- | ---------- |
|  | 04/2020 |  |  |  |  |  |  |  |  |  |  |
| Loret de Mola^S105^  Brazil | 01-12/2019 | 1,028 | GAD-7 | ---------- | ---------- | ---------- | ---------- | GAD-7 ≥ 10 | 9.7 (8.1, 11.7) | 25.9 (23.3, 28.6) | 16.2 (13.2, 19.1) |
|  | 05-07/2020 |  |  |  |  |  |  |  |  |  |  |
| Magson^S87^  Australia | NR/2019 | 126 | SCAS Generalized Anxiety | 5.55 (4.05) | 6.52 (4.31) | 0.97 (NR) | 0.23 (-0.02, 0.48) | ---------- | ---------- | ---------- | ---------- |
|  | 05/2020 |  |  |  |  |  |  |  |  |  |  |
| Rentscher^S29^  USA | 02-06/2019 | 165 | STAI-State | 27.50 (7.10) | 30.60 (9.60) | 3.10 (NR) | 0.37 (0.15, 0.58) | ---------- | ---------- | ---------- | ---------- |
|  | 05-09/2020 |  |  |  |  |  |  |  |  |  |  |
| Rentscher^S29^  USA | 02-06/2019 | 262 | STAI-State | 27.90 (6.50) | 30.10 (9.10) | 2.20 (NR) | 0.28 (0.11, 0.45) | ---------- | ---------- | ---------- | ---------- |
|  | 05-09/2020 |  |  |  |  |  |  |  |  |  |  |
| Rimfeld^S39^  UK | NR/2018 | 2,513 | SMGAD | 8.15 (7.53) | 9.69 (7.69) | 1.54 (7.61) | 0.20 (0.14, 0.26) | ---------- | ---------- | ---------- | ---------- |
|  | 04-05/2020 |  |  |  |  |  |  |  |  |  |  |
| Saraswathi^S62^  India | 12/2019 | 139 | DASS-21 | 4.59 (6.29) | 5.94 (6.93) | 1.35 (NR) | 0.20 (-0.03, 0.44) | DASS-21 Anxiety > 7 | 18.7 (13.1, 26.0) | 32.4 (25.2, 40.5) | 13.7 (4.4, 22.7) |
|  |  |  | Anxiety |  |  |  |  |  |  |  |  |
|  | 06/2020 |  |  |  |  |  |  |  |  |  |  |
| **Men or Males** | | | | | | | | | | | |
| Li, Y^S85^  China | 09/2019 | 503 | ZSAS | EMM (SE): 44.70 (0.59) | EMM (SE): 44.80 (0.62) | NR (NR) | NR (NR) | ---------- | ---------- | ---------- | ---------- |
|  | 03/2020 |  |  |  |  |  |  |  |  |  |  |
| Lim^S122^  USA | NR/2018 | 21 | PROMIS Anxiety | 50.50 (9.90) | 48.60 (11.00) | -1.90 (12.10) | -0.17 (-0.79, 0.45) | ---------- | ---------- | ---------- | ---------- |
|  | 04/2020 |  |  |  |  |  |  |  |  |  |  |
| Magson^S87^  Australia | NR/2019 | 122 | SCAS Generalized Anxiety | 3.63 (3.13) | 3.64 (3.16) | 0.01 (NR) | 0.00 (-0.25, 0.25) | ---------- | ---------- | ---------- | ---------- |
|  | 05/2020 |  |  |  |  |  |  |  |  |  |  |
| Rimfeld^S39^  UK | NR/2018 | 1,050 | SMGAD | 5.88 (6.66) | 6.30 (6.58) | 0.42 (6.62) | 0.06 (-0.02, 0.15) | ---------- | ---------- | ---------- | ---------- |
|  | 04-05/2020 |  |  |  |  |  |  |  |  |  |  |
| Saraswathi^S62^  India | 12/2019 | 78 | DASS-21 Anxiety | 4.62 (6.04) | 6.41 (7.50) | 1.79 (NR) | 0.26 (-0.05, 0.57) | DASS-21 Anxiety > 7 | 25.6 (17.3, 36.3) | 34.6 (25.0, 45.7) | 9.0 (-4.0, 21.5) |
|  | 06/2020 |  |  |  |  |  |  |  |  |  |  |

BSI-18-Anxiety = Brief Symptom Inventory - Anxiety; DASS-21 Anxiety = Depression, Anxiety, and Stress Scale – Anxiety subscale; GAD-2 = Generalized Anxiety Disorder-2; GAD-7 = Generalized Anxiety Disorder-; HBQ = MacArthur Health and Behavior Questionnaire; SCAS = Spence Children's Anxiety Scale; SMGAD= Severity Measure for Generalized Anxiety Disorder; ZSAS = Zung Self-rating Anxiety Scale.

^a^Positive Hedges’ g effect sizes and increases in proportions above a threshold indicate worse mental health in COVID-19 compared to pre-COVID-19. Effects for measures where high scores = positive outcomes were reversed to reflect this. ^b^Not enough information reported to calculate. ^c^Provided by authors. ^d^Included because it is estimated that over 80% of pre-COVID-19 data would have been collected by December 31, 2019.

**Supplementary Table 5.** **Individual Study Results for Depression Symptoms**

| **First Author** | **Pre- and Post-COVID-19 Data Collection** | **N** | **Continuous**  **Outcome Measure** | **Pre-**  **COVID-19 Mean (SD)** | **Post-**  **COVID-19 Mean (SD)** | **Mean (SD) Change**^a^ | **Hedges’ g Standardized Mean Difference (95% CI)** | **Dichotomous Outcome Measure** | **% pre-COVID-19 (95% CI)** | **% post-COVID-19 (95% CI)** | **% Change with 95% CI**^a^ |
| --- | --- | --- | --- | --- | --- | --- | --- | --- | --- | --- | --- |
| **General Population** | | | | | | | | | | | |
| Ge^S5^  China | 01-12/2019 | 1,547-1,978 | PHQ-9 | 12.93 (2.71) | 13.58 (2.46) | 0.65 (NR) | 0.25 (0.19, 0.31) | ---------- | ---------- | ---------- | ---------- |
|  | 02-03/2020 |  |  |  |  |  |  |  |  |  |  |
| Haliwa^S6^  USA | 09-12/2019 | Sample 1: 300; Sample 2: 146; Sample 3: 142 | Sample 1:PHQ-8 Sample 2: DASS-21-Depression Sample 3: PHQ-8 | Sample 1: 5.92 (5.26) Sample 2: 4.81 (5.92) Sample 3: 5.15 (5.81) | Sample 1: 5.79 (6.04) Sample 2: 4.79 (5.67) Sample 3: 5.32 (6.08) | Sample 1: -0.13 (4.09) Sample 2: -0.02 (4.83) Sample 3: 0.17 (4.13) | Sample 1: -0.02 (-0.18, 0.14) Sample 2: 0.00 (-0.23, 0.23) Sample 3: 0.03 (-0.20, 0.26) | Sample 1:PHQ-8 ≥ 10 Sample 2: DASS-21-Depression ≥ 7 Sample 3: PHQ-8 ≥ 10 | Sample 1: 21.3 (17.1, 26.3) Sample 2: 30.8 (23.9, 38.7) Sample 3: 24.6 (18.3, 32.3) | Sample 1: 27.0 (22.3, 32.3) Sample 2: 32.9 (25.8, 40.9) Sample 3: 23.9 (17.7, 31.6) | Sample 1: 5.7 (-0.2, 11.5) Sample 2: 2.1 (-7.1, 11.2) Sample 3: -0.7 (-8.6, 7.2) |
|  | 04-06/2020 |  |  |  |  |  |  |  |  |  |  |
| Kanbur^S7^  Turkey | NR/2019 | 400 | SCL-90-R Depression | 0.33 (NR) | 0.69 (NR) | 0.36 (NR) | NR (NR) | ---------- | ---------- | ---------- | ---------- |
|  | NR/2020 |  |  |  |  |  |  |  |  |  |  |
| Katz, B^S8^  Canada, Ireland, UK, USA | 04/2019 | 218 | DASS-21 Depression | 5.85 (5.64) | 6.28 (5.50) | 0.43 (4.38) | 0.08 (-0.11, 0.26) | ---------- | ---------- | ---------- | ---------- |
|  | 04/2020 |  |  |  |  |  |  |  |  |  |  |
| Wanberg^S18^  USA | 04-06/2019 | 1,117 | PHQ-8 | 4.18 (4.60) | 4.77 (4.83) | 0.59 (NR) | 0.13 (0.04, 0.21) | ---------- | ---------- | ---------- | ---------- |
|  | 04/2020 |  |  |  |  |  |  |  |  |  |  |
| **Older Adults** | | | | | | | | | | | |
| Bartlett^S19^  Australia | 10/2019 | 1,671 | HADS-D | 2.07 (2.09) | 2.05 (2.19) | -0.02 (NR) | -0.01 (-0.08, 0.06) | ---------- | ---------- | ---------- | ---------- |
|  | 04-06/2020 |  |  |  |  |  |  |  |  |  |  |
| Briggs^S20^  Ireland | NR/2018 | 3,490 | CES-D-8 | ---------- | ---------- | ---------- | ---------- | CES-D-8 ≥ 9 | 5.9 (5.1, 6.8) | 19.8 (18.5, 21.2) | 13.9 (12.5, 15.3) |
|  | 07-11/2020 |  |  |  |  |  |  |  |  |  |  |
| Creese^S21^  UK | 10/2019 | 3,281 | PHQ-9 | 2.51 (3.29) | 3.07 (3.58) | 0.56 (NR) | 0.16 (0.11, 0.21) | PHQ-9 ≥ 10 | 4.1 (3.5, 5.0) | 5.6 (4.9, 6.4) | 1.5 (0.6, 2.3) |
|  | 05-06/2020 |  |  |  |  |  |  |  |  |  |  |
| Herrera^S23^  Chile | 11/2019 | 721 | PHQ-9 | 4.25 (NR) | 5.05 (NR) | 0.80 (NR) | NR (NR) | PHQ-9 ≥ 7 | 23.8 (20.8, 27.0) | 30.2 (26.9, 33.6) | 6.4 (2.4, 10.4) |
|  | 09/2020 |  |  |  |  |  |  |  |  |  |  |
| Lee^S26^  Singapore | 12/2017-11/2019 | 496 | PHQ-9 | 0.95 (2.47) | 0.64 (1.49) | -0.31 (NR) | -0.15 (-0.28, -0.03) | PHQ-9 ≥ 6 | 4.8 (3.3, 7.1) | 2.2 (1.2, 3.9) | -2.6 (-4.4, -1.3) |
|  | 05-06/2020 |  |  |  |  |  |  |  |  |  |  |
| Martínez^S27^  Spain | 10/2019 | 141 | CES-D | 11.90 (8.90) | 14.20 (9.10) | 2.30 (9.42) | 0.25 (0.02, 0.49) | ---------- | ---------- | ---------- | ---------- |
|  | 04/2020 |  |  |  |  |  |  |  |  |  |  |
| Rentscher^S29^  USA | NR | 262 | CES-D | 6.30 (7.00) | 8.10 (7.60) | 1.80 (NR) | 0.25 (0.07, 0.42) | ---------- | ---------- | ---------- | ---------- |
|  | 05-09/2020 |  |  |  |  |  |  |  |  |  |  |
| Rentscher^S29^  USA | NR | 165 | CES-D | 4.50 (5.40) | 7.60 (8.00) | 3.10 (NR) | 0.45 (0.23, 0.67) | ---------- | ---------- | ---------- | ---------- |
|  | 05-09/2020 |  |  |  |  |  |  |  |  |  |  |
| Uchida^S128^  Japan | 04/2019-03/2020 | 35 | CES-D-SF | Median (IQR): 8.00 (5.00, 11.00) | Median (IQR): 7.00 (6.00, 9.00) | NR (NR) | NR (NR) | CES-D-SF ≥ 10 | 31.4 (18.6, 48.0) | 22.9 (12.1, 39.0) | -8.6 (-23.0, 6.1) |
|  | 07/2020-03/2021 |  |  |  |  |  |  |  |  |  |  |
| van den Besselaar^S32^  Netherlands | NR/2018-2019 | 984 | CES-D-10 | 4.49 (4.05) | 5.92 (4.11) | 1.43 (NR) | 0.35 (0.26, 0.44) | ---------- | ---------- | ---------- | ---------- |
|  | 06-10/2020 |  |  |  |  |  |  |  |  |  |  |
| Wong, S^S35^  Hong Kong, China | 04/2018-03/2019 | 583 | PHQ-9 | 4.40 (NR) | 4.50 (NR) | 0.19 (NR) | NR^c^ | ---------- | ---------- | ---------- | ---------- |
|  | 03-04/2020 |  |  |  |  |  |  |  |  |  |  |
| Yu^S36^  Singapore | 02/2018-01/2020 | 419 | GDS-15 | 1.02 (1.76) | 2.11 (2.30) | 1.09 (2.10) | 0.53 (0.39, 0.67) | ---------- | ---------- | ---------- | ---------- |
|  | 05-06/2020 |  |  |  |  |  |  |  |  |  |  |
| **Young Adults** | | | | | | | | | | | |
| Marmet^S38^  Switzerland | 04/2019-02/2020^f^ | 2,228 | MDI | 9.07 (7.69) | 7.60 (7.79) | -1.47 (NR) | -0.19 (-0.25, -0.13) | ---------- | ---------- | ---------- | ---------- |
|  | 05-06/2020 |  |  |  |  |  |  |  |  |  |  |
| Rimfeld^S39^  UK | NR/2018 | 3,563-3,694 | SMFQ | 4.36 (4.07) | 4.36 (3.94) | 0.00 (3.82) | 0.00 (-0.05, 0.05) | ---------- | ---------- | ---------- | ---------- |
|  | 04-05/2020 |  |  |  |  |  |  |  |  |  |  |
| Romm^S40^  USA | 09/2019 | 1,082 | PHQ-2 | 1.71 (1.72) | 2.10 (1.74) | 0.38 (1.80) | 0.22 (0.14, 0.30) | ---------- | ---------- | ---------- | ---------- |
|  | 03/2020 |  |  |  |  |  |  |  |  |  |  |
| Watkins-Martin^S43^  Canada | NR/2018 | 1,039 | CES-D-12 | 9.30 (6.42) | 9.59 (6.79) | 0.29 (NR) | 0.04 (-0.04, 0.13) | CES-D-12 ≥ 21 | 6.2 (4.9, 7.8) | 8.1 (6.2, 9.5) | 1.9 (0.2, 3.7) |
|  | 08/2020 |  |  |  |  |  |  |  |  |  |  |
| **University Students** | | | | | | | | | | | |
| Conceição^S44^  Portugal | 10/2019 | 341 | PHQ-9 | 9.66 (7.45) | 12.89 (6.99) | 3.23 (NR) | 0.45 (0.29, 0.60) | PHQ-9 ≥ 15 | 22.6 (18.7, 27.6) | 37.0 (32.3, 42.5) | 14.4 (8.1, 20.5) |
|  | 06/2020 |  |  |  |  |  |  |  |  |  |  |
| Elmer^S46^  Switzerland | 09/2019 | 209 | CES-D | NR | NR | 4.44 (7.23) | 0.53 (0.33, 0.72) | ---------- | ---------- | ---------- | ---------- |
|  | 04/2020 |  |  |  |  |  |  |  |  |  |  |
| Evans^S47^  UK | 10/2019 | 259 | HADS-D | 4.33 (3.26) | 6.31 (3.74) | 1.97 (NR) | 0.56 (0.38, 0.73) | ---------- | ---------- | ---------- | ---------- |
|  | 05/2020 |  |  |  |  |  |  |  |  |  |  |
| Fuller-Rowell^S48^  USA | 09/2018-04/2019 | 263 | BDI-II | 6.50 (7.25) | 10.75 (8.95) | 4.25 (NR) | 0.52 (0.35, 0.69) | ---------- | ---------- | ---------- | ---------- |
|  | 04-06/2020 |  |  |  |  |  |  |  |  |  |  |
| Gelezelyte^S49^  Lithuania | 10-12/2019 | 474 | DASS-21 Depression | 7.88 (5.64) | 7.26 (5.27) | -0.62 (5.25) | -0.11 (-0.24, 0.01) | ---------- | ---------- | ---------- | ---------- |
|  | 10-12/2020 |  |  |  |  |  |  |  |  |  |  |
| Gopalan^S50^  USA | 11/2019 | 1,004 | CES-D-10 | 10.34 (6.21) | 13.12 (6.93) | 2.78 (NR) | 0.42 (0.33, 0.51) | CES-D-10 > 10 | 44.2 (41.2, 47.3) | 60.9 (57.8, 63.8) | 16.6 (11.9 , 21.3) |
|  | 05/2020 |  |  |  |  |  |  |  |  |  |  |
| Hamza^S51^  Canada | 05/2019 | 733 | CES-D-R | 17.62 (13.46) | 18.44 (13.24) | 0.82 (NR) | 0.06 (-0.04, 0.16) | ---------- | ---------- | ---------- | ---------- |
|  | 05/2020 |  |  |  |  |  |  |  |  |  |  |
| Koelen^S53^  Netherlands | 01/2019-01/2020 | 671 | CES-D | ---------- | ---------- | ---------- | ---------- | CES-D ≥ 16 | 48.7 (45.0, 52.5) | 55.3 (51.5, 59.0) | 6.6 (0.9, 12.1) |
|  | 04-05/2020 |  |  |  |  |  |  |  |  |  |  |
| Li, Wendy Wen^S56^  China | 11/2019 | 173 | DASS-21 Depression | 6.25 (6.15) | 4.99 (6.15) | -1.26 (NR) | -0.20 (-0.41, 0.01) | ---------- | ---------- | ---------- | ---------- |
|  | 03/2020 |  |  |  |  |  |  |  |  |  |  |
| Liu^S57^  China | (04-10/2018)-(04-10/2019) | 8079 | PHQ-9 | 4.64 (3.39) | 3.33 (3.90) | -1.31 (NR) | -0.36 (-0.39, -0.33) | PHQ-9 ≥ 10 | 6.6 (6.1, 7.2) | 6.3 (5.7, 7.0) | -0.3 (-0.8, 0.3) |
|  |  |  |  |  |  |  |  | CIDI 3.0 | 2.7 (2.3, 3.1) | 2.1 (1.7, 2.5) | -0.6 (-0.9, -0.3) |
|  | 09-10/2020 |  |  |  |  |  |  |  |  |  |  |
| Lu^S58^  China | 09/2019-10/2019 | 5,181 | PHQ-9 | ---------- | ---------- | ---------- | ---------- | PHQ-9 ≥ 10 | 5.8 (5.1, 6.4) | 7.2 (6.6, 8.0) | 1.4 (0.7, 2.1) |
|  | 04/2020 |  |  |  |  |  |  |  |  |  |  |
| Mehus^S60^  USA | 08,12/2019 | 727 | PHQ-9 | 5.70 (5.09) | 6.83 (5.50) | 1.13 (NR) | 0.21 (0.11, 0.32) | PHQ-9 ≥ 10 | 19.3 (16.6, 22.3) | 27.8 (24.7, 31.2) | 8.5 (4.7, 12.3) |
|  | 04/2020 |  |  |  |  |  |  |  |  |  |  |
| Ratner^S61^  USA | 09/2019 | 152 | BDI-II | 0.38 (0.40) | 0.43 (0.45) | 0.05 (NR) | 0.12 (-0.11, 0.34) | ---------- | ---------- | ---------- | ---------- |
|  | 04/2020 |  |  |  |  |  |  |  |  |  |  |
| Saraswathi^S62^  India | 12/2019 | 217 | DASS-21 Depression | 7.55 (7.86) | 8.16 (8.9) | 0.61 (NR) | 0.07 (-0.12, 0.26) | DASS-21 Depression > 9 | 33.2 (27.3, 39.7) | 35.5 (29.4, 42.1) | 2.3 (-5.6, 10.2) |
|  | 06/2020 |  |  |  |  |  |  |  |  |  |  |
| Shiratori^S65^  Japan | NR/2019 | 6,847 | PHQ-9 | 2.89 (3.44) | 4.05 (4.17) | 1.16 (NR) | 0.30 (0.27, 0.34) | PHQ-9 ≥ 10 | 5.2 (4.7, 5.8) | 9.8 (9.1, 10.5) | 4.6 (3.9, 5.3) |
|  | 06/2020 |  |  |  |  |  |  |  |  |  |  |
| Truskauskaite-Kuneviciene^S66^  Lithuania Germany | 10-12/2019 | Lithuania: 450; Germany: 325 | DASS-21 - Depression | Lithuania: 7.72 (5.66); Germany: 5.09 (4.57) | Lithuania: 6.54 (5.18); Germany: 4.71 (4.36) | Lithuania: -1.18 (NR); Germany: -0.38 (NR) | Lithuania: -0.22 (-0.35, -0.09); Germany: -0.08 (-0.24, 0.07) | ---------- | ---------- | ---------- | ---------- |
|  | 03-04/2020 |  |  |  |  |  |  |  |  |  |  |
| Voltmer^S67^  Germany | NR/2019 | 588 | BSI-18 Depression | 4.80 (5.00) | 4.50 (4.80) | -0.30 (NR) | -0.06 (-0.18, 0.05) | ---------- | ---------- | ---------- | ---------- |
|  | 06/2020 |  |  |  |  |  |  |  |  |  |  |
| Wang, Yitao^S68^  China | 11/2019 | 2,559 | SCL-90-R Depression | 1.55 (0.53) | 1.51 (0.54) | -0.04 (NR) | -0.07 (-0.13, -0.02) | ---------- | ---------- | ---------- | ---------- |
|  | 06/2020 |  |  |  |  |  |  |  |  |  |  |
| Yang, X^S69^  China | 12/2018 | 195 | CES-D | 15.93 (9.97) | 19.08 (6.63) | 3.15 (NR) | 0.37 (0.17, 0.57) | ---------- | ---------- | ---------- | ---------- |
|  | 06/2020 |  |  |  |  |  |  |  |  |  |  |
| Yang, Ziyan^S70^  China | 10/2019 | 2,364 | DASS-21 Depression | 8.87 (2.62) | 8.67 (2.92) | -0.20 (NR) | -0.07 (-0.13, -0.02) | ---------- | ---------- | ---------- | ---------- |
|  | 05/2020 |  |  |  |  |  |  |  |  |  |  |
| Zimmerman^S71^  USA | 08/2019 | 205 | PHQ-9 | 8.91 (6.59) | 12.09 (7.73) | 3.19 (0.51) | 0.44 (0.25,0.64) | ---------- | ---------- | ---------- | ---------- |
|  | 04/2020 |  |  |  |  |  |  |  |  |  |  |
| **Children and Adolescents** | | | | | | | | | | | |
| Adachi^S73^  Japan | 09/2019 | 4,118 | PHQ-A | 4.14 (4.60) | 3.84 (4.24) | -0.30 (NR) | -0.07 (-0.11, -0.02) | PHQ-A ≥ 10 | 12.2 (11.2, 13.2) | 9.9 (9.1, 10.9) | -2.3 (-3.2, -1.3) |
|  | 07/2020 |  |  |  |  |  |  |  |  |  |  |
| Charmaraman^S77^  USA | NR/2019 | 586 | CESDR-10 | NR (NR) | NR (NR) | Change estimate (B): 2.23 | NR (NR) | ---------- | ---------- | ---------- | ---------- |
|  | 10-12/2020 |  |  |  |  |  |  |  |  |  |  |
| Chen, C-Y^S79^  China | 10-11/2019 | 575 | DASS-21 - Depression | 5.86 (7.89) | 5.08 (7.38) | -0.78 (NR) | -0.10 (-0.22, 0.01) | ---------- | ---------- | ---------- | ---------- |
|  | 01/2020 |  |  |  |  |  |  |  |  |  |  |
| Knowles^S84^  UK | NR/2018-2019 | 958-1,055 | SMFQ | ---------- | ---------- | ---------- | ---------- | SMFQ ≥ 12 | 27.8 (22.6, 33.7) | 22.6 (19.3, 26.4) | -5.1 (-8.1, -2.1) |
|  | 05-08/2020 |  |  |  |  |  |  |  |  |  |  |
| Li, Y^S85^  China | 09/2019 | 831 | BDI-II | ---------- | ---------- | ---------- | ---------- | BDI-II > 13 | 35.4 (32.2, 38.7) | 27.8 (24.9, 30.9) | -7.6 (-11.1, -4.0) |
|  | 03/2020 |  |  |  |  |  |  |  |  |  |  |
| Liao^S86^  China | 12/2019 | 2,496 | CES-DC | 15.10 (10.50) | 15.90 (11.10) | 0.80 (NR) | 0.07 (0.02, 0.13) | ---------- | ---------- | ---------- | ---------- |
|  | 07/2020 |  |  |  |  |  |  |  |  |  |  |
| Magson^S87^  Australia | NR/2019 | 248 | SMFQ | 3.81 (4.31) | 6.12 (6.04) | 2.31 (5.81) | 0.44 (0.26, 0.62) | ---------- | ---------- | ---------- | ---------- |
|  | 05/2020 |  |  |  |  |  |  |  |  |  |  |
| Naumann^S90^  Germany | 11/2018-07/2019 | 854 | STDS | ---------- | ---------- | ---------- | ---------- | STDS > 25 | 10.4 (8.4, 12.5) | 25.3 (22.4, 28.2) | 14.9 (11.7, 18.1) |
|  | 05/2020-07/2020 |  |  |  |  |  |  |  |  |  |  |
| Polack^S92^  USA | 01-09/2019 | 112 | CDI-S | 2.85 (3.14) | 3.96 (3.79) | 1.11 (2.77) | 0.32 (0.06, 0.58) | ---------- | ---------- | ---------- | ---------- |
|  | 03-06/2020 |  |  |  |  |  |  |  |  |  |  |
| Rau^S93^  Germany | 10-11/2019 | 777 | RCADS - Depression | 6.27 (5.30) | 5.41 (5.35) | -0.86 (NR) | -0.16 (-0.26, -0.06) | ---------- | ---------- | ---------- | ---------- |
|  | 06-07/2020 |  |  |  |  |  |  |  |  |  |  |
| Shoshani^S94^  Israel | 09/2019 | 1,537 | BSI-18 - Depression | 6.14 (4.73) | 7.59 (5.25) | 1.45 (NR) | 0.29 (0.22, 0.36) | ---------- | ---------- | ---------- | ---------- |
|  | 05/2020 |  |  |  |  |  |  |  |  |  |  |
| Teng^S95^  China | 10-11/2019 | 1,778 | CES-D | 0.81 (0.59) | 0.83 (0.60) | 0.02 (NR) | 0.03 (-0.03, 0.10) | ---------- | ---------- | ---------- | ---------- |
|  | 04-05/2020 |  |  |  |  |  |  |  |  |  |  |
| Wang, Wanxin^S97^  China | 10-12/2019 | 1,790 | CES-D | 13.69 (10.53) | 13.44 (10.28) | -0.25 (NR) | -0.02 (-0.09, 0.04) | CES-D ≥ 16 | 30.0 (27.9, 32.2) | 29.2 (27.1, 31.4) | -0.8 (-3.3, 1.7) |
|  | 10-12/2020 |  |  |  |  |  |  |  |  |  |  |
| Wong, R^S99^  China | 04-08/2019 | 233 | DASS-21 - Depression | NR (NR) | NR (NR) | 0.84 (6.22) | NR (NR) | ---------- | ---------- | ---------- | ---------- |
|  | 02/2020 |  |  |  |  |  |  |  |  |  |  |
| Yang, Zhengqian^S100^  China | 11/2019 | 1,125 | CES-D | 0.94 (0.63) | 0.75 (0.64) | -0.19 (NR) | -0.30 (-0.38, -0.22) | ---------- | ---------- | ---------- | ---------- |
|  | 08/2020 |  |  |  |  |  |  |  |  |  |  |
| Zhang^S101^  China | 11/2019 | 1,241 | MFQ | 16.6 (12.20) | 17.7 (14.40) | 1.49 (11.41) | 0.11 (0.03, 0.19) | ---------- | ---------- | ---------- | ---------- |
|  | 05/2020 |  |  |  |  |  |  |  |  |  |  |
| **Parents** | | | | | | | | | | | |
| Adesogan^S102^  USA | NR/2018-03/2020 | 329 | CES-D | 6.56 (4.22) | 8.22 (5.52) | 1.66 (NR) | 0.34 (0.18, 0.49) | ---------- | ---------- | ---------- | ---------- |
|  | 06-09/2020 |  |  |  |  |  |  |  |  |  |  |
| Frank^S103^  USA | 08/2018 | 180 | PHQ-9 | 3.65 (5.77) | 4.33 (6.24) | 0.68 (NR) | 0.11 (-0.09, 0.32) | ---------- | ---------- | ---------- | ---------- |
|  | 08/2020 |  |  |  |  |  |  |  |  |  |  |
| Loret de Mola^S105^  Brazil | 01-12/2019 | 1,042 | EPDS | ---------- | ---------- | ---------- | ---------- | EPDS ≥ 13 | 5.1 (3.8, 6.5) | 29.5 (26.6, 32.1) | 24.4 (21.3, 27.2) |
|  | 05-07/2020 |  |  |  |  |  |  |  |  |  |  |
| Pitchik^S106^  Bangladesh | 05-06/2019 | 517 | CES-D | 13.40 (8.70) | 12.80 (9.20) | -0.60 (NR) | -0.07 (-0.19, 0.05) | ---------- | ---------- | ---------- | ---------- |
|  | 07-09/2020 |  |  |  |  |  |  |  |  |  |  |
| Rivera^S107^  Mexico | NR/2018-2019 | 466 | EDS | 7.48 (5.80) | 7.34 (5.83) | -0.14 (NR) | -0.02 (-0.15, 0.10) | EDS > 12 | 19.5 (16.4, 23.6) | 19.1 (16.0, 23.1) | -0.4 (-4.4, 3.5) |
|  | 05-11/2020 |  |  |  |  |  |  |  |  |  |  |
| Thompson^S108^  USA | NR/2018-2019 | 147 | CES-D | 14.22 (10.13) | 19.28 (11.74) | 5.06 (13.08) | 0.46 (0.23, 0.69) | ---------- | ---------- | ---------- | ---------- |
|  | 04/2020 |  |  |  |  |  |  |  |  |  |  |
| **People with Pre-existing Medical Conditions** | | | | | | | | | | | |
| Becker^S109^  USA | 03/2019 | 121 | CESD-10 | 8.40 (5.50) | 9.10 (5.60) | 0.70 (NR) | 0.13 (-0.13, 0.38) | ---------- | ---------- | ---------- | ---------- |
|  | 03/2020 |  |  |  |  |  |  |  |  |  |  |
| Chao^S111^  USA | 02/2018-02/2020 | 2,679 | PHQ-8 | 2.50 (3.30) | 3.50 (4.00) | 1.00 (NR) | 0.27 (0.22, 0.33) | PHQ-8 ≥ 10 | 4.6 (3.8, 5.5) | 8.5 (7.4, 9.6) | 3.9 (2.8, 5.0) |
|  | 07-12/2020 |  |  |  |  |  |  |  |  |  |  |
| Chiu^S112^  USA | 10/2018 | 133 | HADS-D | 4.61 (3.65) | 5.82 (3.85) | 1.21 (3.50) | 0.32 (0.08, 0.56) | ---------- | ---------- | ---------- | ---------- |
|  | 09/2020 |  |  |  |  |  |  |  |  |  |  |
| Derksen^S113^  Netherlands | 01/2019-01/2020 | 2176 | HADS-D | 2.98 (3.22) | 2.78 (3.10) | -0.20 (3.20) | -0.06 (-0.12, 0.00) | ---------- | ---------- | ---------- | ---------- |
|  | 04-06/2020 |  |  |  |  |  |  |  |  |  |  |
| Dunlop-Thomas^S114^  USA | NR/2017-2019 | 852 | PROMIS - Depression | 51.40 (10.65) | 49.80 (9.87) | -1.60 (NR) | -0.16 (-0.25, -0.06) | ---------- | ---------- | ---------- | ---------- |
|  | NR/2020-2021 |  |  |  |  |  |  |  |  |  |  |
| Fujiwara^S115^  Japan | 07-09/2019 | 245 | HADS-D | Median (IQR): 7.00 (6.00) | Median (IQR): 7.00 (6.00) | NR (NR) | NR (NR) | ---------- | ---------- | ---------- | ---------- |
|  | 07-09/2020 |  |  |  |  |  |  |  |  |  |  |
| García-Rudolph^S116^  Spain | NR | 175 | HADS-D | 4.63 (4.25) | 5.73 (4.95) | 1.10 (NR) | 0.24 (0.03, 0.45) | ---------- | ---------- | ---------- | ---------- |
|  | 11/2020 |  |  |  |  |  |  |  |  |  |  |
| Gul^S117^  Turkey | 10-11/2019 | 116 | BDI | 11.53 (9.40) | 12.54 (11.30) | 1.01 (NR) | 0.10 (-0.16, 0.35) | BDI ≥ 19 | 17.2 (11.5, 25.1) | 23.3 (16.5, 31.8) | 6.0 (-2.8, 14.8) |
|  | 06-07/2020 |  |  |  |  |  |  |  |  |  |  |
| Henry^S118^  Canada, France, UK, USA | 07-12/2019 | 388 | PHQ-8 |  |  |  |  | ---------- | ---------- | ---------- | ---------- |
|  | 04/2020 |  |  | 6.73 (5.73) | 6.44 (5.44) | -0.29 (NR) | -0.05 (-0.19, 0.09) |  |  |  |  |
|  | 09-10/2020 |  |  | 6.73 (5.73) | 5.59 (5.05) | -1.14 (NR) | -0.21 (-0.35, -0.07) |  |  |  |  |
|  | 03/2021 |  |  | 6.73 (5.73) | 5.60 (5.28) | -1.13 (NR) | -0.20 (-0.35, -0.06) |  |  |  |  |
| Johnstone^S119^  New Zealand | NR/2018 | 104 | HADS-D | 3.82 (3.12) | 3.80 (3.39) | -0.02 (NR) | -0.01 (-0.28, 0.26) | ---------- | ---------- | ---------- | ---------- |
|  | 07-09/2020 |  |  |  |  |  |  |  |  |  |  |
| Katz, P^S120^  USA | NR/2019 | 1,504 | PHQ-2 | 0.79 (1.25) | 0.84 (1.24) | 0.05 (NR) | 0.04 (-0.03, 0.11) | ---------- | ---------- | ---------- | ---------- |
|  | 03-06/2020 |  |  |  |  |  |  |  |  |  |  |
| Liang^S121^  China | 12/2019 | 114 | ZSDS | 37.70 (9.10) | 37.40 (9.50) | -0.3 (NR) | -0.03 (-0.31, 0.24) | ---------- | ---------- | ---------- | ---------- |
|  | 02-03/2020 |  |  |  |  |  |  |  |  |  |  |
| Lim^S122^  USA | NR/2018 | 316 | PROMIS Depression | 50.80 (10.50) | 49.30 (9.80) | -1.50 (9.20) | -0.15 (-0.30, 0.01) | ---------- | ---------- | ---------- | ---------- |
|  | 04/2020 |  |  |  |  |  |  |  |  |  |  |
| Park^S124^  Germany | 09/2019–02/2020 | 152 | HADS-D | Median (IQR): 5.00 (2.00, 7.00) | Medina (IQR): 4.00 (2.00, 7.00) | NR (NR) | NR (NR) | HADS-D ≥ 8 | 13.0 (8.7, 19.5) | 8.0 (4.6, 13.3) | -5.3 (-10.3, -0.8) |
|  | 05-08/2020 |  |  |  |  |  |  |  |  |  |  |
| Rentscher^S29^  USA | NR | 262 | CES-D | 6.30 (7.00) | 8.10 (7.60) | 1.80 (NR) | 0.25 (0.07, 0.42) | ---------- | ---------- | ---------- | ---------- |
|  | 05-09/2020 |  |  |  |  |  |  |  |  |  |  |
| Sacre^S125^  Australia | NR/2018-2019 | 450 | PHQ-8 | 4.10 (4.70) | 4.10 (4.70) | 0.00 (NR) | 0.00 (-0.13, 0.13) | PHQ-8 ≥ 10 | 5.3 (3.6, 7.8) | 5.6 (3.8, 8.1) | 0.3 (-2.1, 2.5) |
|  | 05-06/2020 |  |  |  |  |  |  |  |  |  |  |
| Sbragia^S126^  Italy | 01/2019 | 106 | HADS-D | 5.93 (3.77) | 5.51 (3.93) | -0.42 (NR) | -0.11 (-0.38, 0.16) | HADS-D > 8 | 38.1 (29.1, 47.2) | 34.0 (25.7, 43.4) | -3.8 (-14.6, 7.2) |
|  | 05/2020 |  |  |  |  |  |  |  |  |  |  |
| Ubara^S127^  Japan | 04-07/2019 | 164 | PHQ-9 | Median (IQR): | Median (IQR): | NR^c^ | NR^c^ | ---------- | ---------- | ---------- | ---------- |
|  |  |  |  | 2.00 (1.00-5.00) | 3.00 (0.25-6.00) |  |  |  |  |  |  |
|  | 05/2020 |  |  |  |  |  |  |  |  |  |  |
| Uchida^S128^  Japan | 04/2019-03/2020 | 142 | CES-D-SF | Median (IQR): 6.00 (4.80, 10.00) | Median (IQR): 7.00 (5.00, 10.00) | NR (NR) | NR (NR) | CES-D-SF ≥ 10 | 26.1 (19.5, 33.8) | 26.1 (19.5, 33.8) | 0.0 (-8.3, 8.3) |
|  | 07/2020-03/2021 |  |  |  |  |  |  |  |  |  |  |
| Wong, S^S35^  Hong Kong, China | 04/2018-03/2019 | 583 | PHQ-9 | 4.40 (NR) | 4.50 (NR) | 0.19 (NR) | NR^c^ | ---------- | ---------- | ---------- | ---------- |
|  | 03-04/2020 |  |  |  |  |  |  |  |  |  |  |
| Young^S132^  UK | 09/2018-02/2020 | 12,098 | PHQ-9 | 11.18 (6.86) | 10.80 (6.68) | -0.38 (NR) | -0.06 (-0.08, -0.03) | ---------- | ---------- | ---------- | ---------- |
|  | 04-09/2020 |  |  |  |  |  |  |  |  |  |  |
| **People with Pre-existing Mental Health Conditions** | | | | | | | | | | | |
| Gentile^S129^  Italy, Paraguay | 10-12/2019 | 110 | HAM-D | 11.40 (7.26) | 11.90 (7.56) | 0.50 (NR) | 0.07 (-0.20, 0.33) | ---------- | ---------- | ---------- | ---------- |
|  | 03-04/2020 |  |  |  |  |  |  |  |  |  |  |
| Swerdlow^S131^  USA | NR/2018-04/2020 | 144 | MASQ-30 - Depression | 35.99 (6.83) | 35.74 (7.36) | -0.25 (NR) | -0.04 (-0.26, 0.19) | ---------- | ---------- | ---------- | ---------- |
|  | 04-06/2020 |  |  |  |  |  |  |  |  |  |  |
| Young^S132^  UK | 09/2018-02/2020 | 12,098 | PHQ-9 | 11.18 (6.86) | 10.80 (6.68) | -0.38 (NR) | -0.06 (-0.08, -0.03) | ---------- | ---------- | ---------- | ---------- |
|  | 04-09/2020 |  |  |  |  |  |  |  |  |  |  |
| **Medical Staff** | | | | | | | | | | | |
| Frank^S103^  USA | 08/2018 | 180 | PHQ-9 | 3.65 (5.77) | 4.33 (6.24) | 0.68 (NR) | 0.11 (-0.09, 0.32) | ---------- | ---------- | ---------- | ---------- |
|  | 08/2020 |  |  |  |  |  |  |  |  |  |  |
| Li, Weidong^S133^  China | 10-11/2019 | 385 | PHQ-9 | 5.17 (NR) | 5.77 (NR) | 0.60 (NR) | NR^c^ | ---------- | ---------- | ---------- | ---------- |
|  | 01-02/2020 |  |  |  |  |  |  |  |  |  |  |
| **Sexual or Gender Minority Individuals** | | | | | | | | | | | |
| Bavinton^S134^  Australia | NR/2019 | 681 | PHQ-9 | 5.98 (5.93) | 6.56 (6.03) | 0.58 (NR) | 0.10 (-0.01, 0.20) | ---------- | ---------- | ---------- | ---------- |
|  | 04/2020 |  |  |  |  |  |  |  |  |  |  |
| Flentje^S135^  USA | 06/2019 | 2,280 | PHQ-9 | 7.10 (5.99) | 8.31 (6.43) | 1.21 (5.10) | 0.19 (0.14, 0.25) | ---------- | ---------- | ---------- | ---------- |
|  | 03-04/2020 |  |  |  |  |  |  |  |  |  |  |
| Ghabrial^S136^  Canada | NR/2019 | 780 | CES-D | 14.47 (7.69) | 16.50 (7.34) | 2.03 (NR) | 0.27 (0.17, 0.37) | ---------- | ---------- | ---------- | ---------- |
|  | 09-10/2020 |  |  |  |  |  |  |  |  |  |  |
| **Women or Females** | | | | | | | | | | | |
| Adesogan^S102^  USA | NR/2018-03/2020 | 191 | CES-D | 6.82 (4.70) | 9.23 (5.77) | 2.41 (NR) | 0.46 (0.25, 0.66) | ---------- | ---------- | ---------- | ---------- |
|  | 06-09/2020 |  |  |  |  |  |  |  |  |  |  |
| Frank^S103^  USA | 08/2018 | 95 | PHQ-9 | 3.69 (5.26) | 5.05 (6.64) | 1.36 (NR) | 0.23 (-0.06, 0.51) | ---------- | ---------- | ---------- | ---------- |
|  | 08/2020 |  |  |  |  |  |  |  |  |  |  |
| Li, Y^S85^  China | 09/2019 | 328 | BDI-II | EMM (SE): 13.02 (0.54) | EMM (SE): 10.77 (0.55) | NR (NR) | NR (NR) | ---------- | ---------- | ---------- | ---------- |
|  | 03/2020 |  |  |  |  |  |  |  |  |  |  |
| Lim^S122^  USA | NR/2018 | 295 | PROMIS Depression | 50.80 (10.70) | 49.30 (9.80) | -1.50 (9.30) | -0.15 (-0.31, 0.02) | ---------- | ---------- | ---------- | ---------- |
|  | 04/2020 |  |  |  |  |  |  |  |  |  |  |
| Loret de Mola^S105^  Brazil | 01-12/2019 | 1,042 | EPDS | ---------- | ---------- | ---------- | ---------- | EPDS ≥ 13 | 5.1 (3.8, 6.5) | 29.5 (26.6, 32.1) | 24.4 (21.3, 27.2) |
|  | 05-07/2020 |  |  |  |  |  |  |  |  |  |  |
| Magson^S87^  Australia | NR/2019 | 126 | SMFQ | 4.77 (5.00) | 8.16 (6.46) | 3.39 (NR) | 0.58 (0.33, 0.83) | ---------- | ---------- | ---------- | ---------- |
|  | 05/2020 |  |  |  |  |  |  |  |  |  |  |
| Rentscher^S29^  USA | NR | 165 | CES-D | 4.50 (5.40) | 7.60 (8.00) | 3.10 (NR) | 0.45 (0.23, 0.67) | ---------- | ---------- | ---------- | ---------- |
|  | 05-09/2020 |  |  |  |  |  |  |  |  |  |  |
| Rentscher^S29^  USA | NR | 262 | CES-D | 6.30 (7.00) | 8.10 (7.60) | 1.80 (NR) | 0.25 (0.07, 0.42) | ---------- | ---------- | ---------- | ---------- |
|  | 05-09/2020 |  |  |  |  |  |  |  |  |  |  |
| Rimfeld^S39^  UK | NR/2018 | 2,578 | SMFQ | 4.65 (4.20) | 4.81 (4.07) | 0.16 (4.14) | 0.04 (-0.02, 0.09) | ---------- | ---------- | ---------- | ---------- |
|  | 04-05/2020 |  |  |  |  |  |  |  |  |  |  |
| Saraswathi^S62^  India | 12/2019 | 139 | DASS-21 Depression | 7.71 (7.57) | 7.94 (8.77) | 0.23 (NR) | 0.03 (-0.21, 0.26) | DASS-21 Depression > 9 | 36.7 (29.1, 45.0) | 34.5 (27.1, 42.8) | -2.2 (-11.7, 7.4) |
|  | 06/2020 |  |  |  |  |  |  |  |  |  |  |
| Uchida^S128^  Japan | 04/2019-03/2020 | 60 | CES-D-SF | Median (IQR): 7.00 (5.00, 9.00) | Median (IQR): 7.00 (5.00, 10.00) | NR (NR) | NR (NR) | CES-D-SF ≥ 10 | 23.3 (14.4, 35.4) | 28.3 (18.5, 40.8) | 5.0 (-8.4, 18.2) |
|  | 07/2020-03/2021 |  |  |  |  |  |  |  |  |  |  |
| **Men or Males** | | | | | | | | | | | |
| Adesogan^S102^  USA | NR/2018-03/2020 | 138 | CES-D | 6.20 (3.46) | 6.86 (5.17) | 0.66 (NR) | 0.15 (-0.09, 0.38) | ---------- | ---------- | ---------- | ---------- |
|  | 06-09/2020 |  |  |  |  |  |  |  |  |  |  |
| Frank^S103^  USA | 08/2018 | 85 | PHQ-9 | 3.60 (6.30) | 3.52 (5.75) | -0.08 (NR) | -0.01 (-0.32, 0.29) | ---------- | ---------- | ---------- | ---------- |
|  | 08/2020 |  |  |  |  |  |  |  |  |  |  |
| Li, Y^S85^  China | 09/2019 | 503 | BDI-II | EMM (SE): 9.25 (0.63) | EMM (SE): 7.87 (0.66) | NR (NR) | NR (NR) | ---------- | ---------- | ---------- | ---------- |
|  | 03/2020 |  |  |  |  |  |  |  |  |  |  |
| Lim^S122^  USA | NR/2018 | 21 | PROMIS Depression | 50.70 (8.60) | 49.90 (9.70) | -0.80 (8.20) | -0.08 (-0.70, 0.54) | ---------- | ---------- | ---------- | ---------- |
|  | 04/2020 |  |  |  |  |  |  |  |  |  |  |
| Magson^S87^  Australia | NR/2019 | 122 | SMFQ | 2.81 (3.18) | 4.02 (4.76) | 1.21 (NR) | 0.30 (0.05, 0.55) | ---------- | ---------- | ---------- | ---------- |
|  | 05/2020 |  |  |  |  |  |  |  |  |  |  |
| Marmet^S38^  Switzerland | 04/2019-02/2020 | 2,345 | MDI | 9.07 (7.69) | 7.60 (7.79) | -1.47 (NR) | -0.19 (-0.25, -0.13) | ---------- | ---------- | ---------- | ---------- |
|  | 05-06/2020 |  |  |  |  |  |  |  |  |  |  |
| Rimfeld^S39^  UK | NR/2018 | 1,116 | SMFQ | 3.71 (3.70) | 3.33 (3.40) | -0.38 (3.55) | -0.11 (-0.19, -0.02) | ---------- | ---------- | ---------- | ---------- |
|  | 04-05/2020 |  |  |  |  |  |  |  |  |  |  |
| Saraswathi^S62^  India | 12/2019 | 78 | DASS-21 Depression | 7.28 (8.40) | 8.54 (9.17) | 1.26 (NR) | 0.14 (-0.17, 0.45) | DASS-21 Depression > 9 | 26.9 (18.3, 37.7) | 37.2 (27.3, 48.3) | 10.3 (-2.9, 22.9) |
|  | 06/2020 |  |  |  |  |  |  |  |  |  |  |
| Uchida^S128^  Japan | 04/2019-03/2020 | 82 | CES-D-SF | Median (IQR): 6.00 (4.00, 10.00) | Median (IQR): 7.00 (4.80, 9.30) | NR (NR) | NR (NR) | CES-D-SF ≥ 10 | 28.1 (19.5, 38.6) | 24.4 (16.4, 34.7) | -3.7 (-14.0, 6.7) |
|  | 07/2020-03/2021 |  |  |  |  |  |  |  |  |  |  |
| **Immigrants** |  |  |  |  |  |  |  |  |  |  |  |
| Gosselin^S137^  France | 04/2018-NR/2019 | 100 | PHQ-9 | ---------- | ---------- | ---------- | ---------- | PHQ-9 ≥ 10 | 65.0 (55.3, 73.6) | 72.0 (62.5, 79.9) | 7.0 (-9.4, 23.0) |
|  | 06/2020 |  |  |  |  |  |  |  |  |  |  |

BSI-18-Depression = Brief Symptom Inventory - Depression; CES-D= Center for Epidemiologic Studies Depression Scale; DASS-21 Depression = Depression, Anxiety, and Stress Scale – Depression subscale; MDI= Major Depression Inventory; MFQ = Mood and Feelings Questionnaire; PHQ-2 = Patient Health Questionnaire-2; PHQ-8 = Patient Health Questionnaire-8; PHQ-9 = Patient Health Questionnaire-9; SMFQ = Short Mood and Feelings Questionnaire; ZSDS= Zung Self-rating Depression Scale.

^a^Positive Hedges’ g effect sizes and increases in proportions above a threshold indicate worse mental health in COVID-19 compared to pre-COVID-19. Effects for measures where high scores = positive outcomes were reversed to reflect this. ^b^Not enough information reported to calculate. ^c^Provided by authors. ^d^Included because it is estimated that over 80% of pre-COVID-19 data would have been collected by December 31, 2019.

**Supplementary Figure 1**. Sensitivity Analysis of General Mental Health conducted with results from Savage et al. from October 2020 instead of April 2020

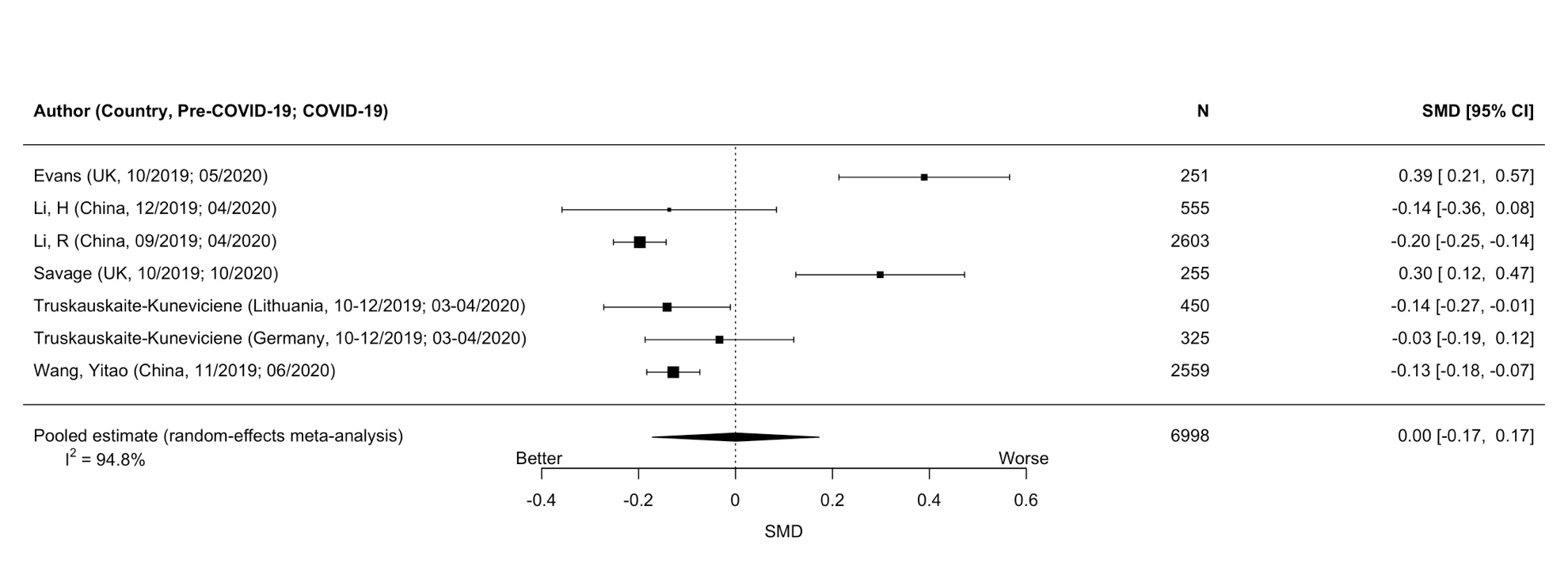

**Supplementary Figure 2.** Sensitivity Analysis of Anxiety Symptoms conducted with results from Henry et al. from September to October 2020 instead of April 2020

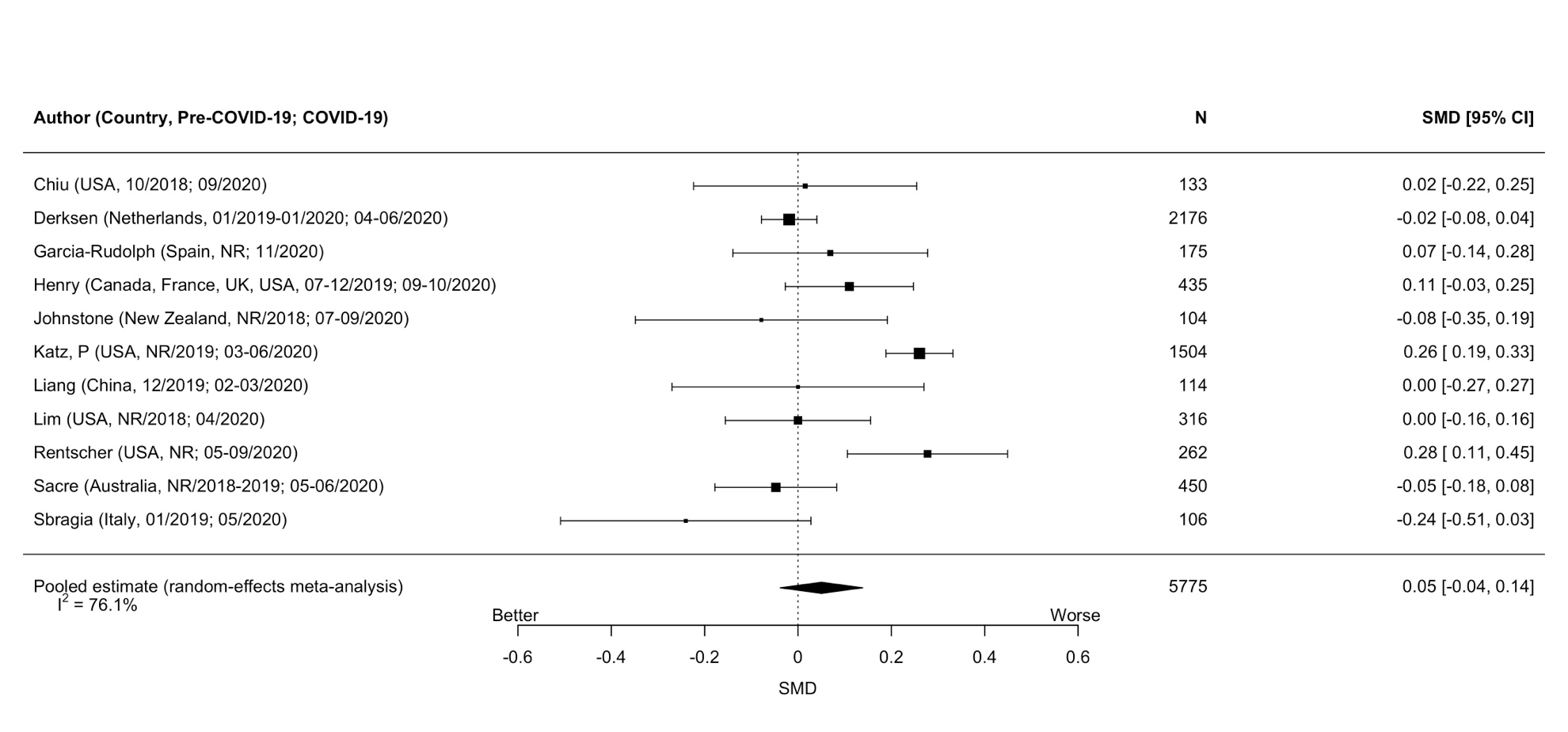

**Supplementary Figure 3.** Sensitivity Analysis of Anxiety Symptoms conducted with results from Henry et al. from March 2021 instead of April 2020
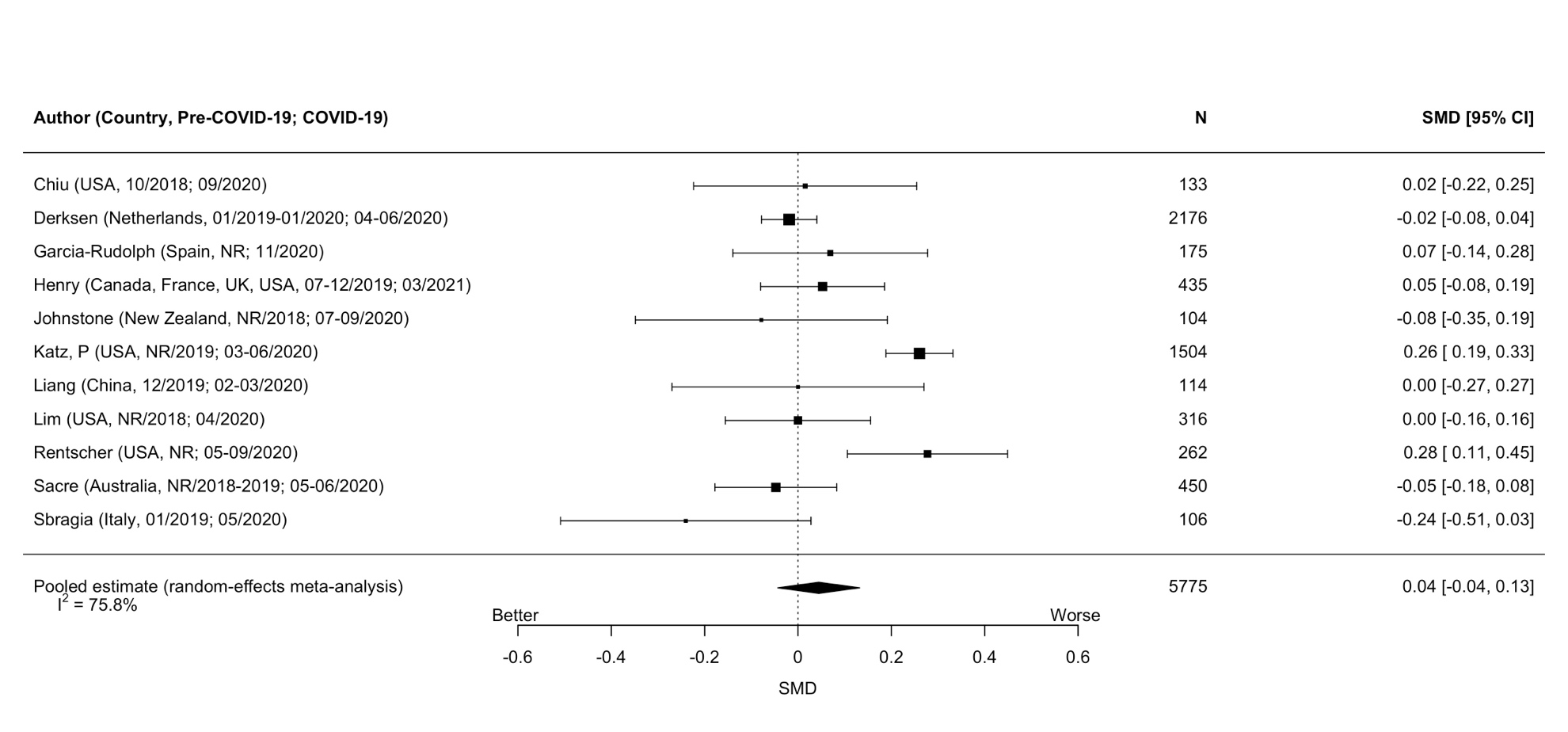

**Supplementary Figure 4.** Sensitivity Analysis of Depression Symptoms conducted with results from Henry et al. from September to October 2020 instead of April 2020
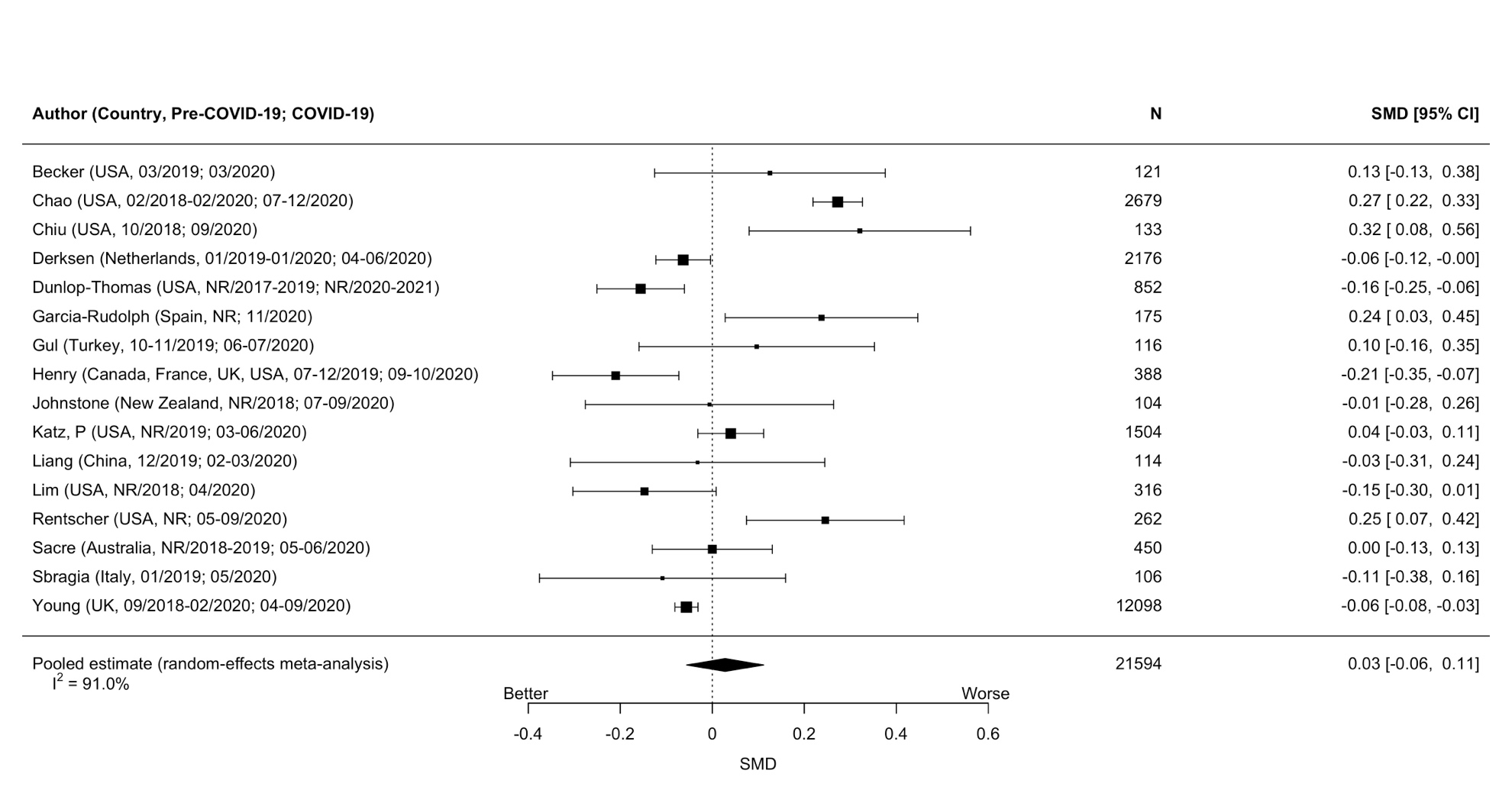

**Supplementary Figure 5.** Sensitivity Analysis of Depression Symptoms conducted with results from Henry et al. from March 2021 instead of April 2020
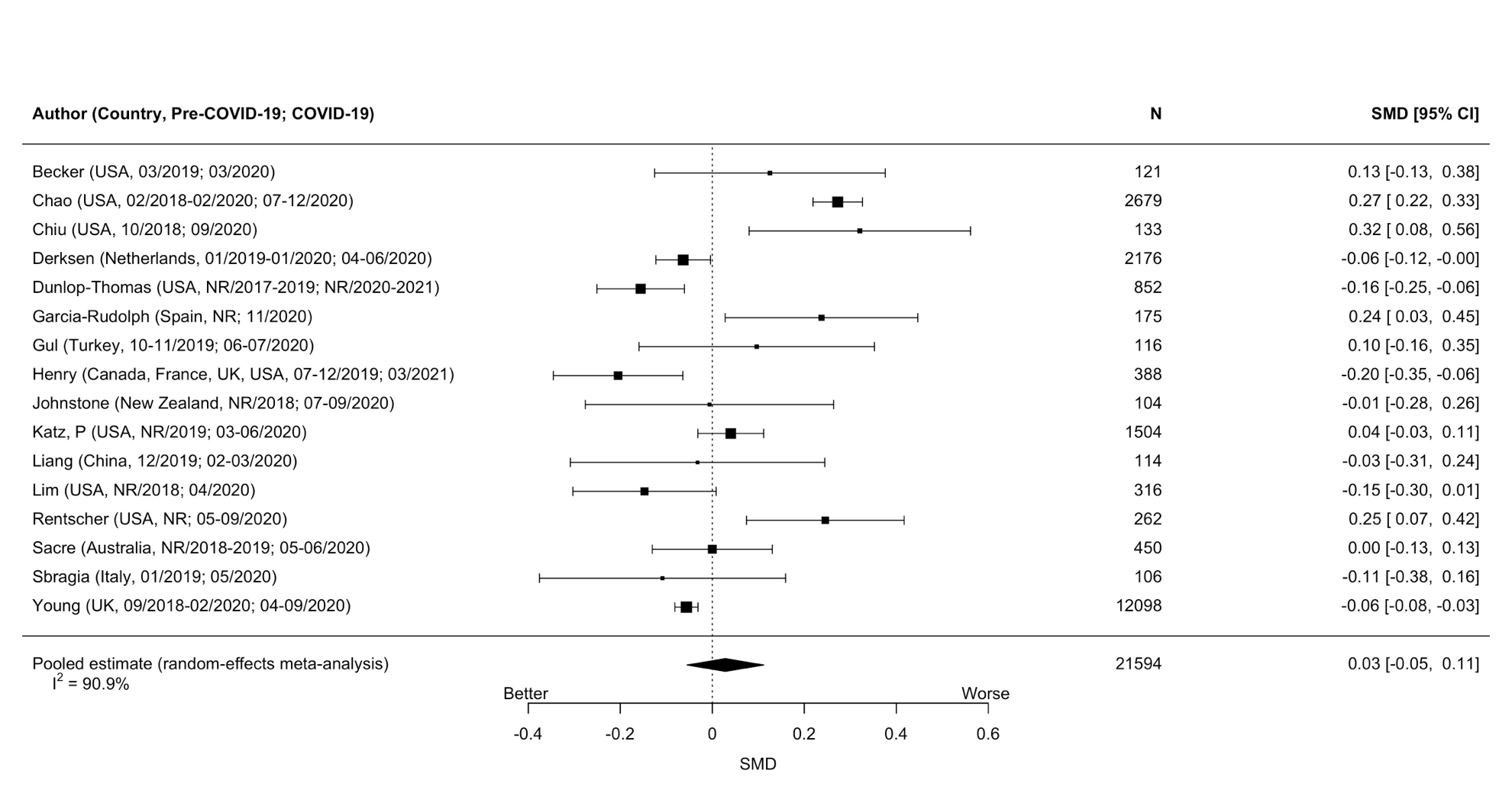
